# The costs and cost-effectiveness of different service models of palliative care, focusing on end of life care: A rapid review

**DOI:** 10.1101/2024.03.06.24303850

**Authors:** Llinos Haf Spencer, Bethany Fern Anthony, Jacob Davies, Kalpa Pisavadia, Liz Gillen, Jayne Noyes, Deborah Fitzsimmons, Ruth Lewis, Alison Cooper, Dyfrig Hughes, Rhiannon Tudor Edwards, Adrian Edwards

**Affiliations:** Centre for Health Economics and Medicines Evaluation (CHEME), Bangor University, United Kingdom; Bangor Institute for Medical and Health Research (BIMHR), Bangor University, United Kingdom; Wales Centre for Evidence Based Care (WCEBC), Cardiff University, United Kingdom; Swansea Centre for Health Economics (SCHE), Swansea University, United Kingdom; Health and Care Research Wales Evidence Centre, Bangor University, United Kingdom; Health and Care Research Wales Evidence Centre, Cardiff University, United Kingdom

## Abstract

Some people receive palliative or end of life care at home, others in hospitals or hospices, or a combination of home and hospice/home and hospital models. This rapid review aims to determine the costs and cost-effectiveness of different service models of palliative care or end of life care. These studies are mostly conducted from the perspective of the healthcare system, disregarding costs related to patients/caregivers economic burden (Perea-Bello et al., 2023).

**Research Implications and Evidence Gaps:** More UK research is needed on cost impacts of new services such as Enhanced Supported Care (ESC). Future research should consider which methods are most appropriate to evaluate palliative care models. Standard methodology, such as the calculation of quality-adjusted life years (QALYs), may not be most appropriate for this end of life population.

Improving QALYs may not be the intended aim of palliative care or end of life interventions, and prolonging death may be inconsistent with patient preferences and wishes. The quality and applicability of the evidence we found in our rapid review were variable, and therefore, uncertainty remains, especially when the perspective of analysis was not stated clearly.

Therefore, it was difficult to ascertain whether all relevant costs were considered. Assumptions on costs were not varied in many studies, and most studies had different time horizons.

**Policy and Practice Implications:** This rapid review has shown that **hospital-based palliative care costs are higher than hospice or home-based** palliative care. This suggests that **home-based palliative care should be available to all** patients in a recognisable end of life phase who desire to remain and die at home. Healthcare planners should **aim to reduce hospitalisation at the end of life** but **only if access to quality home care** at the end of life is guaranteed. **Patients should have a choice** about where they prefer to die without moving the costs from the healthcare system to the home caregivers, rendering the costs invisible.

**Funding Statement:** The Bangor Institute for Medical and Health Research, was funded for this work by the Health and Care Research Wales Evidence Centre, itself funded by Health and Care Research Wales on behalf of Welsh Government.

## 2. BACKGROUND

### 2.1 Who is this review for?

This Rapid Review was conducted as part of the Health and Care Research Wales Evidence Centre Work Programme. The research question was suggested by the Palliative and End of Life Care Delivery Plan team. The intended audience are palliative care service commissioners and policy makers in Wales.

### 2.2 Background and purpose of this review

Palliative care is an interdisciplinary medical caregiving approach to optimise quality of life and mitigate suffering among people with serious, complex, and often terminal illnesses (World Health Organisation (WHO), 2023). End of life care is defined by NICE as care that is provided in the ‘last year of life’ (National Institute for Health and Care Excellence (NICE), 2019). This time frame can be difficult to predict, so some people might only receive end of life care in their last weeks or days, and others may have end of life care for longer (NHS England, 2023a).

There are many different models of palliative care, with some people receiving palliative care or end of life care at home and others in hospitals or hospices(National Institute for Health and Care Excellence (NICE), 2019). In the United Kingdom (UK), much of the palliative care provided in the community is delivered by General Practitioners (GPs), practice nurses and district nurses. Patients also move from home to hospital, sometimes to hospices, and back and forth between a variety of services several times (Papworth et al., 2023). In the UK, adult, children and young people’s palliative care are commissioned separately and managed within clinical palliative care networks (Noyes, Edwards, et al., 2013). Children’s palliative care is considered a relatively new subspeciality (established in the 1980s), and neonatal palliative care is the latest advancement of the overall model (Kain & Chin, 2020).

Hospice care aims to improve the quality of lives and wellbeing of adults, young people and children who have a terminal illness (Dreamscape and Hospice UK, 2023; Noyes, Edwards, et al., 2013; Noyes, Hastings, et al., 2013; The Kings Fund, 2018; Ziwary et al., 2017). In Wales, most hospice care is provided at home by third sector services such as Marie Curie. However, it can also be provided in a care home, as an in-patient at the hospice itself, or as a day patient visiting the hospice (Baker, 2020; McBride et al., 2011; National Health Service (NHS), 2022; The National Gold Standards Framework (GSF) Centre in End of Life Care, 2022; Wheatley & Baker, 2007). There are also nurse led service models of palliative care (Dumont et al., 2022; Salamanca-Balen et al., 2018).

For this review, the term ‘model of palliative care’ was defined as any structured care model involving multiple components, including ‘who delivers (e.g., professionals, paid carers) the intervention (specialist or generalist palliative care), where (setting, e.g., hospital), to whom (care recipients), when (i.e. timing and duration), how (e.g., face to face) and for what purpose (i.e. expected outcomes)? (Brereton et al., 2017; Davidson et al., 2006).

Generalist palliative care is provided by professionals from primary care who have good basic palliative care skills and knowledge (National Institute for Health and Care Excellence (NICE), 2023). Specialised palliative care is provided by expert clinicians who provide assessment, advice, and responsive care to people with progressive life-limiting illnesses by managing physical symptoms such as pain as well as psychological and spiritual distress (NHS England, 2023b).

The aim of this rapid review is to investigate the costs and outcomes of different service models of palliative care with a focus on end of life care. The rapid review question was: What evidence is available on the costs and cost-effectiveness of different service models of palliative care, with a focus on end of life care?

## 3. RESULTS

The rapid review search strategy is presented in Appendix 1 (see Section 8.1). The searches yielded a possible 101 studies, which were sought for full text screening (see Figure 1 in Section 6 for the PRISMA diagram). Fifty-six studies were deemed relevant, of which eight were systematic reviews, and 48 were primary studies.

**Figure 1.**
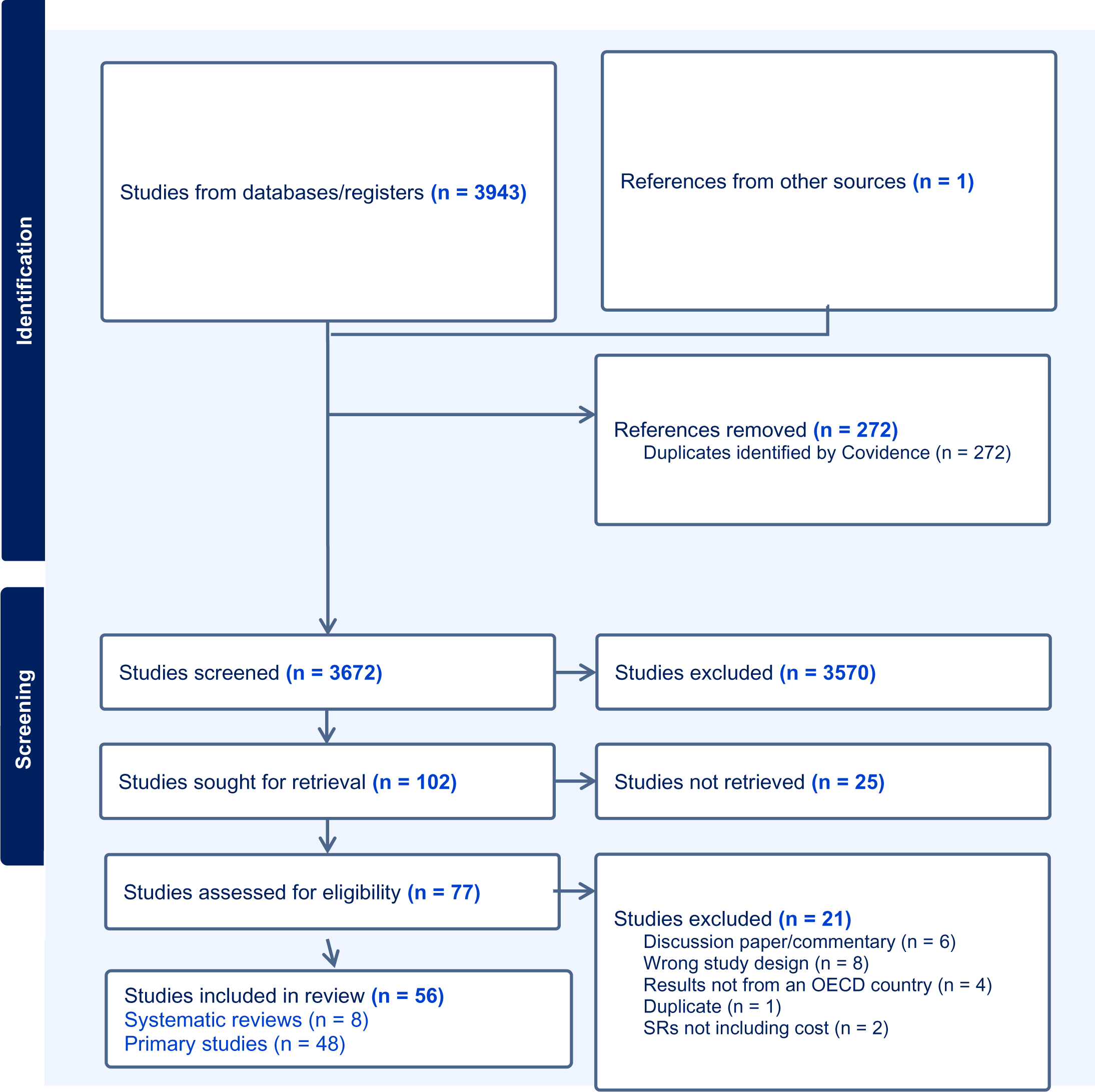
PRISMA 2020 flow diagram of included studies (Page et al., 2021).

Eight systematic reviews (SRs) were included as part of this rapid review (RR). These SRs were conducted by authors in Belgium (Simoens et al., 2010), Canada (Mathew et al., 2020), Ireland (Smith et al., 2014), Switzerland (Gonzalez-Jaramillo et al., 2021), United Kingdom (UK) (Bajwah et al., 2020; Gomes et al., 2013; Salamanca-Balen et al., 2018), and the United States of America (USA) (Yadav et al., 2020). Seven of the SRs were deemed of high quality, and one was of moderate quality (Smith et al., 2014) using the Joanna Briggs Institute (JBI) checklist for Systematic Reviews and Research Synthesis (Joanna Briggs Institute, 2017).

In terms of the UK SRs, the Bajwah et al., (2020) SR reported 13 studies which included costs of hospital palliative care (HSPC), n= 9 studies found no difference between HSPC, and usual care and two studies favoured HSPC over usual care. The difference in cost was unclear in one study. Another study reported mixed findings with lower cost of hospitalisation in favour of HSPC but no difference in the cost of emergency room visits. Four studies with full economic evaluations were inconclusive on the cost-effectiveness of HSPC (Bajwah et al., 2020). In the Gomes et al (2013) SR only two UK cost studies were reported from within the time period of this RR, with results suggesting that home based palliative care was less costly than hospital based palliative care (Gomes et al., 2013).

In a 2018 SR, two economic studies were identified. A short-term cost-minimisation study examining a telephone follow-up Clinical Nurse Specialist (CNS) led intervention for patients with breast cancer in the UK, and a cost analysis of effects of practice nurse-led care for chronic diseases. In both cases, the interventions were associated with higher costs compared to usual care although some patients preferred these care models (Salamanca-Balen et al., 2018).The SRs are presented in further detail in the evidence section (Section 7) and are not considered further in this results section as the emphasis for this RR was on the primary studies.

Of the 48 primary studies, most were cost analyses (n=39), n=1 was an SROI study (Hughes, 2021), n=5 were full economic evaluations (Isenberg et al., 2017; Pham & Krahn, 2014; Rosato et al., 2021; Saygili & Çelik, 2019; Sellars et al., 2019) and n=3 were Markov modelling studies (Kim et al., 2022; McBride et al., 2011; Nguyen et al., 2017). Of the full economic evaluations, n=5 were cost-effectiveness analyses (Isenberg et al., 2017; Pham & Krahn, 2014; Rosato et al., 2021; Saygili & Çelik, 2019; Sellars et al., 2019). A detailed summary of each included study is provided in Appendix 9.2.

Nine primary studies focused on hospital palliative care costs, three studies focused on hospice palliative care costs, 15 studies focused on community palliative care costs, and 22 studies either described or compared different models of palliative care. Of the 49 primary studies, two studies described paediatric models of palliative care, and 47 studies described adult models of palliative care. One paediatric study is described within the hospice section, and the other paediatric study is described within the community-based models of care section below.

### 3.1 Hospital palliative care costs

Nine studies were included which reported the costs of palliative care in hospital settings (Hanson et al., 2008; Isenberg et al., 2017; Kerr et al., 2017; McCarthy et al., 2015; Nathaniel et al., 2015; Pollock et al., 2022; Schneider et al., 2020; Sellars et al., 2019; Tan & Jatoi, 2011). With regards to quality as assessed by the JBI critical appraisal checklist for economic evaluations, n=2 were deemed to be of high quality (McCarthy et al., 2015; Sellars et al., 2019), n=6 were of moderate quality (Isenberg et al., 2017; Kerr et al., 2017; Nathaniel et al., 2015; Pollock et al., 2022; Schneider et al., 2020; Tan & Jatoi, 2011), and n=1 of low quality (Hanson et al., 2008). Most of these studies (n=6) were from the USA, and there were also studies from Australia (Sellars et al., 2019), the Netherlands (Schneider et al., 2020) and the UK (Kerr et al., 2017). N =7 studies were from a healthcare system perspective, and n=2 studies did not define the perspective of analysis. The results of these nine studies will be described under the following themes: palliative cancer care, palliative kidney disease care, general palliative care, and economic evaluations focusing on palliative care.

#### 3.1.1 Cost studies of hospital-based specialist palliative care (cancer)

Two of the included studies focused on specialist cancer palliative care within hospital settings. One was from the USA, and the other was from The Netherlands. A retrospective cost analysis from the USA examined n=120 patients with solid tumour diagnoses admitted to hospital for end of life care. The median total cost required to provide a medical service per patient hospitalisation episode was $12,962. However, the cost per oncology patient was higher at $25,320. When adjusting for patient age at death (median age of 61 years) and days in hospital (median length of stay of four days), advance directives and route of hospitalisation were not associated with a statistically significant difference in hospital costs, with p > 0.24 and p > 0.51 respectively (Tan & Jatoi, 2011).

A cost analysis study from the Netherlands examining hospital costs of patients who died of advanced breast cancer in hospital (n=558) found overall monthly hospital costs (including medication, treatment, diagnostics, and consultation costs) of €2,255 (SD = €492) per patient. The mean cost per patient across the final twelve months of life was €21,641 (SD = €20,147). In the first seven months of admission, monthly costs remained stable, with a mean of €1,984. From the eighth month before death until the final month before death, mean costs per month steadily increased with an average increase of €343 per month, reaching a maximum of €3,614 during the last month before death. Medication costs fell after the third month of admission, while hospitalisation costs contributed to the increased cost at the end of life (Schneider et al., 2020).

#### 3.1.2 Cost studies of hospital-based specialist palliative care (chronic kidney disease)

Two studies were included, which focused on hospital-based specialist palliative care for patients with chronic kidney disease (CKD). The studies were from the UK (Kerr et al., 2017) and the USA (Pollock et al., 2022).

A prospective cost analysis also investigating place of death, from the UK of n=211,215 patients receiving hospital care for CKD found that the mean cost of hospital admissions and outpatient care in the last twelve months before death was £11,916 for people with CKD and £7,832 for patients with other conditions. Costs increased by more than 50% in the final three months before death. The study also assessed the impact place of death had on the cost of hospital care. For people with CKD, the mean cost of hospital care in the twelve months prior to death was £9,877 for those who died at home and £12,160 for those who died elsewhere. The mean cost in the final thirty days of life was £1,077 for those who died at home and £3,206 for those who died elsewhere. People with CKD were less likely to die at home than those without CKD in every age category. Death at home was more common in men than in women in all age groups aged over 30 years old, for those with and without CKD. Overall, 12.5% of men and 8.5% of women with CKD died at home (Kerr et al., 2017).

A recent retrospective cost analysis study from the USA investigated the end of life hospital costs for patients admitted with CKD, cardiovascular (CV) and infection related admissions failure related encounters incurred longer hospital stays and higher costs than either CV or infection related encounters. The median total cost of a single patient encounter was $17,057. When disaggregated into reason for admission, the median costs were $18,469, $17,503 and $16,403 for CKD, CV and infection related admissions, respectively (Pollock et al., 2022).

#### 3.1.3 Cost studies of hospital-based generalist palliative care

Three studies described the costs of hospital-based generalist palliative care. These studies were all from the USA (Hanson et al., 2008; McCarthy et al., 2015; Nathaniel et al., 2015).

A cost analysis study published in 2008 described the impact of interdisciplinary palliative care consultations on hospital costs at the end of life. When compared to a matched control group (matched on diagnosis), palliative care had no significant difference in variable costs across the entire hospitalisation period, with mean costs of $16,748 and $15,926, respectively (P > 0.78). The length of stay between the palliative care and control group was also not significantly different (16.6 and 13.8 days, respectively) (Hanson et al., 2008).

A retrospective modelling study investigating the cost of palliative care from the USA matched PC patients to non-PC patients (separately by discharge status) using propensity score methods). The per patient stay of Individuals who died in hospital after having received palliative care, including palliative care from a palliative care physician and palliative care registered nurse, cost $3,426 less per inpatient stay than the control group (discharge status matched controls). For the cohort of patients dying in hospital, costs without a palliative care consultation were estimated to be $33,075 compared to costs of $29,649 for patients with a palliative care physician and research nurse consultation. Consultations initiated within the first ten days of inpatient stay showed significant savings in both cohorts, with mean savings of $2,696 among patients discharged alive and $9,689 among patients who died in hospital (McCarthy et al., 2015).

A retrospective cost analysis of a hospital-based palliative care unit in the USA found that the mean cost per patient per day was $1,522 in the days before transfer to the palliative care unit (PCU). Following transfer to the PCU, the mean cost fell to $835, a saving of $687 in daily patient costs (this did not account for confounding factors). Among patients who died in the hospital, average daily direct cost per patient in the days after transfer to PCU were statistically significantly lower by $240 as compared with patients being followed by Palliative Care Consultation Service (PCCS) on the general hospital wards (SE = $45, P < 0.001) (Nathaniel et al., 2015).

#### 3.1.4 Economic evaluations of hospital-based palliative care

Two cost-effectiveness studies investigated hospital-based palliative care services. One study was from Australia (Sellars et al., 2019), and the other was from the USA (Isenberg et al., 2017).

A cost-effectiveness study conducted in Australia with a hypothetical cohort of patients with Chronic Kidney Disease (CKD) focussed on Advance Care Planning (ACP). Advanced care planning is a voluntary process of person-centred discussion between an individual and their care providers about their preferences and priorities for their future care. The comparison group was treated with home haemodialysis (usual end of life care). The ACP intervention was, on average, AUS$519 per patient. In the decision analytic model, the average cost per patient for the ACP group was $100,579 (SD = 17,356), and the proportion of patients receiving end of life care according to preferences was 68% (SD = 48). In the no ACP group, the average cost per patient was $87,282 (SD = 19,078), and the proportion of patients having preferences met was 24% (SD = 43). The last twelve months of ACP was more expensive yet more effective in facilitating adherence to patient preferences than usual care. The incremental cost per additional case of end of life preferences being met (incremental cost-effectiveness ratio [ICER]) was $28,421. Using the Australian cost-effectiveness threshold, ACP would be highly cost-effective (Sellars et al., 2019).

Another cost-effectiveness study from the USA aimed to establish the costs of an inpatient palliative care unit (PCU) and conduct a threshold analysis to estimate the maximum possible costs for the PCU to be considered cost-effective. The authors found that PCU was cost saving, but not cost-effective compared to usual care without the inpatient PCU. The PCU can be cost-effective if the variable costs were under $559,800 (an additional $716 per patient encounter per day) (Isenberg et al., 2017). Quality appraisal of the hospital focussed studies can be viewed Appendix 9.2. See Table 1 for cost-effectiveness study tables showing the main results from See Table 2 for the table of inflated and converted hospital costs. (Caution should be taken when reading this table as the time horizons are different in nearly every row). The list of inflation calculators used for inflating and converting costs from the original currency into GBP is presented in Appendix 9.3.

**Table 1.**
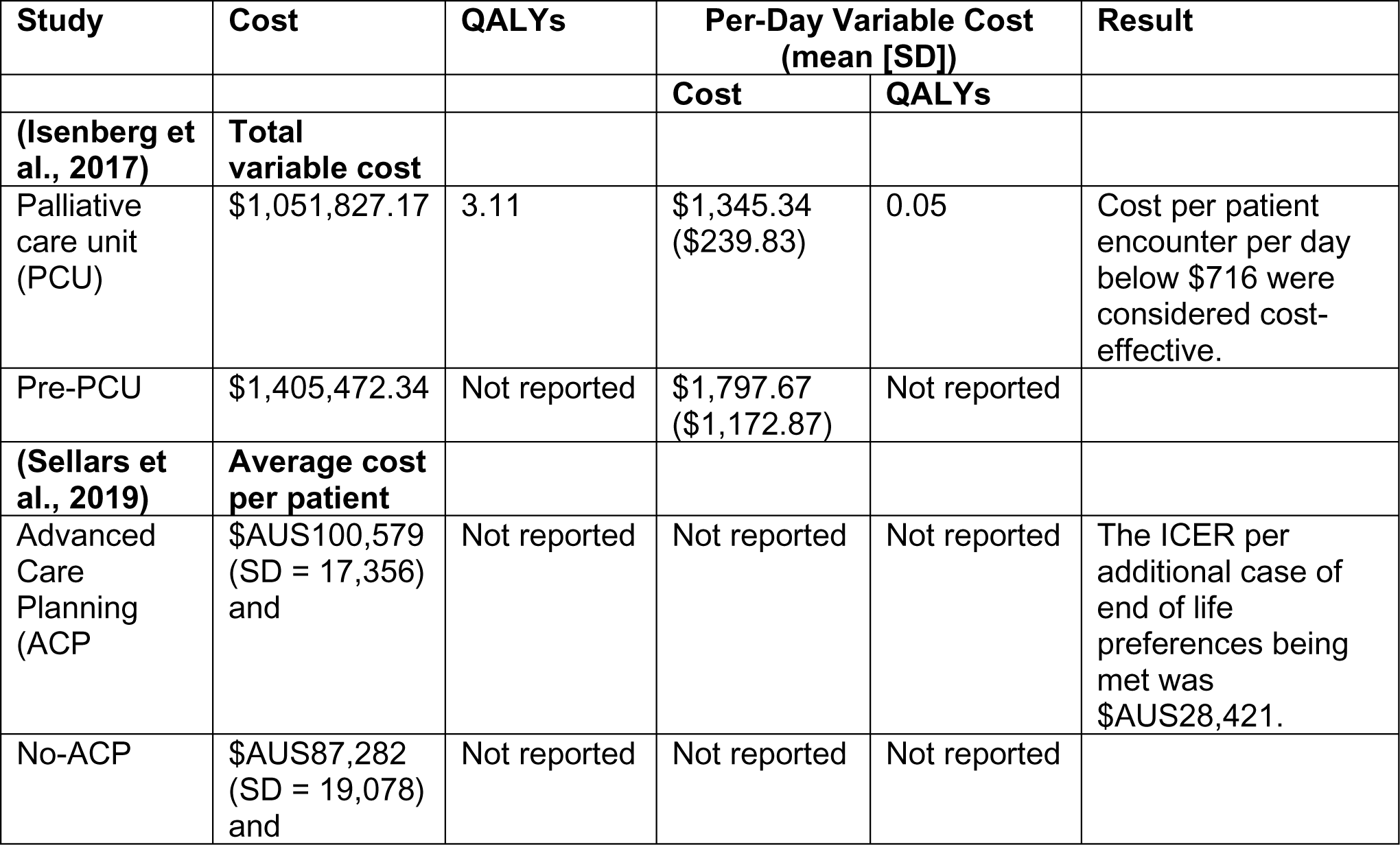
Summary of cost-effectiveness studies relating to hospital palliative care.

**Table 2.** Hospital palliative care costs table.

| Citation | Cost year | Description of cost | Original currency | Currency in 2023 | GBP in 2023 |
| --- | --- | --- | --- | --- | --- |
| Tan and Jatoi, 2011 | 2011 | Median hospital stay | US\$12,692 | US\$17,360 | £13,737 |
| Schneider et al., 2020 | 2020 | Mean end of life care cost | Euro 21,641 | Euro 26,870 | £23,023 |
| Kerr et al., 2017 | 2017 | Cost of hospital admission 12 months before death – Chronic Kidney Disease | £11,916 | £15,218 | £15,218 |
| Kerr et al., 2017 | 2017 | Cost of hospital admission 12 months before death – Non-CKD | £7,832 | £10,002 | £10,002 |
| Pollock et al | 2019 | Cost of any inpatient encounter between January 2016 – March 2020 (All patients) | US\$17,057 | US\$20,527 | £16,266 |
| Pollock et al | 2019 | Cost of any inpatient encounter between January 2016 – March 2020 (Kidney failure) | US\$18,469 | US\$22,227 | £17,613 |
| Pollock et al | 2019 | Cost of any inpatient encounter between January 2016 – March 2020 (Cardiovascular disease) | US\$17,503 | US\$21,064 | £16,692 |
| Hanson et al., 2008 | 2004 | Cost of usual care at end of life | US\$16,748 | US\$27,278 | £21,663 |
| Hanson et al., 2008 | 2004 | Cost of palliative end of life care after PC consultation | US\$15,926 | US\$25,939 | £20,599 |
| McCarthy et al., 2015 | 2015 | Cost of usual care at end of life | US\$33,075 | US\$42,935 | £34,103 |
| McCarthy et al., 2015 | 2015 | Cost of palliative end of life care after PC consultation | US\$29,649 | US\$38,487 | £30,570 |
| Nathaniel et al., 2015 | 2013 | Cost saving following implementation of a palliative care unit (versus a prior palliative care counselling service, PCCS) | US\$687 | US\$907 | £720 |
| Isenberg et al., 2017<br>Cost-effectiveness | 2017 | Difference between pre-PCU (higher cost) and PCU (lower cost) in the model. | US\$452.33 | US\$568 | £451 |
| Sellars et al., 2019<br>Cost-effectiveness | 2019 | Advanced care planning group | AUS\$110,579 | AUS\$122,273 | £63,626 |
| Sellars et al., 2019<br>Cost-effectiveness | 2019 | Non-Advanced care planning group | AUS\$87,282 | AUS\$96,512 | £50,280 |

#### 3.1.5 Bottom line results for costs of hospital-based palliative care

**Most of the hospital-based palliative care studies found that hospital-based palliative care costs increased in the last thirty days of life. This is due to increased hospitalisation costs as the condition of the patients deteriorates (Pollock et al., 2022; Tan & Jatoi, 2011). However, there was some evidence to indicate that if a palliative consultation was done prior to death, the hospitalisation costs were less as palliative care consultation is followed by decisions to forego costly treatment, resulting in greater cost-savings (Hanson et al., 2008; Isenberg et al., 2017; McCarthy et al., 2015). Advanced care planning is more costly but more effective in facilitating adherence to patient preferences for end of life care (Sellars et al., 2019).**

### 3.2 Hospice palliative care costs

Four hospice focused cost papers were included in the rapid review. Two were from the USA (Gans et al., 2016; Huskamp et al., 2008), and two were from the UK (Hughes, 2021; Mitchell et al., 2020). Three publications were focused on costs, and there was also one Social Return on Investment (SROI) study comparing the social returns from inpatient and day therapy (Hughes, 2021). In terms of quality appraisal, n=1 was of high quality (Mitchell et al., 2020), and n=3 were of moderate quality (Gans et al., 2016; Hughes, 2021; Huskamp et al., 2008). Two of the studies were from a provider perspective (Gans et al., 2016; Huskamp et al., 2008), one was from an NHS perspective in the UK (Mitchell et al., 2020), and one was from the perspective of hospice providers in Wales, UK (Hughes, 2021). See Table 2 below for converted currency inflated to 2023. The list of inflation calculators used for inflating and converting costs from the original currency into GBP is presented in Appendix 9.3.

A 2020 cost analysis study from the UK aimed to examine and estimate the effects of Palliative Care Day Services (PCDS) in three centres across the UK: England, Scotland, and Northern Ireland. As well as costs, quality of life questionnaires were included (including EQ-5D-5L and ICECAP-SCM). The mean cost per attendee per day ranged from £121 to £190 across the three centres (Mitchell et al., 2020). The other UK study conducted an SROI, which yielded a greater social return on investment for those receiving palliative care at home, in comparison with those who received inpatient palliative care at a hospice in North Wales (SROI ratios of £8.97 and £2.81 respectively) (Hughes, 2021).

A cost analysis study from the USA examined patient level cost data. The mean daily cost of hospice stay was $329 in 2008. This mean cost was higher on the first and last days of hospice care. The authors found that the costs would differ for younger and older adults, with younger adults being more costly as some elements of elderly care might have already been covered by residential nursing care costs (Huskamp et al., 2008). Another study from the USA focused on the Medicaid cost of child palliative care in a hospice compared with hospice-like services for children and investigated programme enrolment data. Hospice-like paediatric palliative care programme model was $3331 less costly than the traditional hospice model ($15,643 Vs $12,312) (Gans et al., 2016). Quality appraisal of the hospice focussed studies can be viewed in Appendix 9.2. See Table 3 for the table of inflated and converted hospice costs. (Caution should be taken when reading this table as the time horizons are different in nearly every row). The list of inflation calculators used for inflating and converting costs from the original currency into GBP is presented in Appendix 9.3.

**Table 3.** Hospice palliative care costs table.

| Citation | Cost year | Description of cost | Original currency | Currency in 2023 | GBP in 2023 |
| --- | --- | --- | --- | --- | --- |
| Gans et al, 2016 | 2016 | Usual care cost per month. | US\$15,643 | US\$20,05.15 | £15,898 |
| Gans et al, 2016 | 2016 | Cost for children after enrolment of Partners for Children (PFC) intervention per month. | US\$12,312 | US\$15,783 | £12,510 |
| Gans et al, 2016 | 2016 | Difference in cost between usual care and PFC per month. | US\$4,897 | US\$6,278 | £4976 |
| Hughes, 2021 | Assumed 2021 | SROI ratio for inpatient hospice care | GBP 2.81 (rounded up to £3) | £3.54 | £3.54 |
| Hughes, 2021 | Assumed 2021 | SROI ratio for day therapy | GBP 8.97 (rounded up to £9) | £10.62 | £10.62 |
| Huskamp et al, 2008 | 2008 | Mean total cost per hospice stay (various lengths of stays) | US\$2,192 | US\$3,132 | £2,483 |
| Mitchell et al, 2020 | 2018 | Mean cost per attendee/day range | £121 to £190 | £151-£237 | £151-£237 |
| Mitchell et al, 2020 | 2018 | The cost of providing PCDS when value of volunteering is accounted for. | £172 to £264 | £214-£329 | £214-£329 |

#### 3.2.1 Bottom line results for costs of hospice-based palliative care

**A paediatric palliative care programme for children with palliative care needs (who received the Partners for Children (PFC) intervention) cost £4,976 less than usual care without the extra support (Gans et al., 2016). The way that palliative care and end of life care costs are calculated vary. For example, one study calculated the cost per day of hospice care of between £151–£237 (Mitchell et al., 2020). However, other studies have calculated the mean total cost per hospice stay (of varying lengths) and found the mean cost per stay to be £2,483 in cost year 2023 (Huskamp et al., 2008). Due to the various lengths of stay, these palliative care costs are not comparable. This rapid review presents evidence that palliative care is more costly on the first and last days of end of life care in hospices and less costly for elderly people in the USA who live in residential care (due to USA medical insurance systems such as Medicare accounting for health and social care separately) (Comans et al., 2021). The SROI study conducted in Wales clearly showed a better social return on investment for those receiving palliative care at home, in comparison with those who received inpatient palliative care at a hospice in North Wales (SROI ratios of £8.97 and £2.81, respectively) (Hughes, 2021).**

### 3.3 Home-based or community-based palliative care and end of life care

Fifteen papers that describe models of palliative or end of life care delivered in the community setting were included. The perspective of analysis from seven of these studies was not defined. In terms of quality appraisal using the JBI critical appraisal checklist for economic evaluation, n=9 were deemed to be of high quality (Amador et al., 2014; Bentur et al., 2014; Butler et al., 2022; Chai et al., 2014; Gage et al., 2015; Maetens et al., 2019; Pham & Krahn, 2014; Rosato et al., 2021; Spiro et al., 2020), n=3 were of moderate quality (Chen et al., 2018; Johnson et al., 2009; Klinger et al., 2013) and n=3 were of low quality (Enguidanos et al., 2005; Goldhagen et al., 2016; Gordon et al., 2022). One study was from a payer perspective (Chai et al., 2014), four studies were from a healthcare system perspective (Gage et al., 2015; Johnson et al., 2009; Klinger et al., 2013; Rosato et al., 2021), one study was from a payer and a healthcare system perspective (Pham & Krahn, 2014) and one study was from a third party and patient co-payment perspective (Maetens et al., 2019). Thirteen were cost analyses, and two were full economic evaluations (Pham & Krahn, 2014; Rosato et al., 2021). Of the fifteen community-based palliative and end of life care studies, four were conducted in England (Amador et al., 2014; Butler et al., 2022; Gage et al., 2015; Spiro et al., 2020), four in Canada (Chai et al., 2014; Johnson et al., 2009; Klinger et al., 2013; Pham & Krahn, 2014) four in the USA (Chen et al., 2018; Enguidanos et al., 2005; Goldhagen et al., 2016; Gordon et al., 2022), one in Israel (Bentur et al., 2014), one in Belgium (Maetens et al., 2019), and one in Italy (Rosato et al., 2021). Most of the identified studies reported on home-based models of palliative and end of life care (n=13). The remaining two studies assessed the costs of end of life care in residential care homes (Amador et al., 2014) and community-based paediatric palliative care (Goldhagen et al., 2016). Quality appraisal of the fifteen community-based studies can be viewed in Appendix 9.2.

#### 3.3.1 Cost analyses of home-based models

Eleven cost analyses of home-based models of palliative and end of life care were identified. Of these, three were conducted in England (Butler et al., 2022; Gage et al., 2015; Spiro et al., 2020), three in the USA (Chen et al., 2018; Enguidanos et al., 2005; Gordon et al., 2022), three in Canada (Chai et al., 2014; Johnson et al., 2009; Klinger et al., 2013), one in Israel (Bentur et al., 2014), and one in Belgium (Maetens et al., 2019). The study characteristics and main findings from the cost analyses of home-based palliative and end of life care models can be found in Section 6 of this report. The studies are described via country below.

##### 3.3.1.1 England, UK

A realist-informed mixed methods evaluation to determine optimum levels of hospice at home services with an included cost analysis of hospice at home models was conducted in England (Butler et al., 2022). Findings indicated that costs increased with proximity to death (median daily costs for days 0–14 were £104.57, compared with £56.07 for days 29–92).

Median daily formal care costs were significantly lower (£40.43, £27.93 and £12.22 for 0–14, 15–28 and 29–92 days before death, respectively) than median daily informal care costs (£580.00, £449.50 and £348.00 for 0–14, 15–28 and 29–92 days before death, respectively) (Butler et al., 2022).

From an NHS perspective, resource utilisation cost analysis study was conducted in England in 2020 to (i) compare the characteristics of rapid response service (RSS) users and non-users, and (ii) explore differences in the proportions of users and non-users dying in the place of their choice. Preferences for place of death were obtained from hospice records. (Gage et al., 2015). Findings indicated no significant differences in total service costs between users and non-users of the rapid response service. Nevertheless, overall costs were higher for rapid response users who were referred to the service two days prior to death due to the costs associated with rapid response service input and other community costs (Gage et al., 2015). A pilot cost analysis of a visit-based home care service collected day-to-day costs of hospice-at-home services for up to two weeks in the last three months of life (Spiro et al., 2020). Health and social care resource use diaries collected data for n = 333 days of hospice-at-home care completed by n = 30 families, which equated to an average of n = 11 days per family and n = 708 staff visits (equating to n = 604 hours) at a total cost of £20,192 (Spiro et al., 2020).

##### 3.3.1.2 USA

Searches identified three cost analyses of home-based palliative and end of life care models in the USA. Firstly, a retrospective matched cohort study assessed Medicare reimbursement savings for before and after the enrolment into a home-based palliative care programme for older frail adults with advanced medical illness (Chen et al., 2018). There was a statistically significant difference between the intervention and propensity matched control group (p < 0.001) in favour of the home-based palliative care intervention, with annual Medicare saving of $18,251 per patient compared with matched control patients (Chen et al., 2018). Another study from the USA assessed healthcare resource use costs for congestive heart failure (CHF), chronic obstructive pulmonary disease (COPD) and cancer patients who received home-based palliative care compared with usual care at the end of life (Enguidanos et al., 2005). Home-based palliative care resulted in significant cost savings for patients for all patient groups compared to usual care. For example, patients with COPD enrolled in the palliative care group spent $11,325 less on average compared to those in usual care, amounting to a 67% decrease in the cost of care (Enguidanos et al., 2005). A further costing analysis study from the USA aimed to evaluate an adult home palliative care (HPC) programme for multiple insurance product lines conducted from a payer perspective (Gordon et al., 2022) 2021). Enrolment in the programme was associated with medical cost savings of $24,643 per member for the calendar year 2019 (Gordon et al., 2022).

##### 3.3.1.3 Canada

Three studies were cost analyses of home-based palliative care models in Canada. From a societal perspective, in 2014, a cost analysis study (of the last 12 months of life) was conducted to explore the extent of unpaid care costs in total healthcare costs for home-based palliative care patients with malignant neoplasm. Findings demonstrated that unpaid care accounts for a considerable proportion (77%) of monthly home-based palliative care costs in the last twelve years of life, at an average monthly cost per patient of $11 334. For all cost categories, monthly costs increased significantly with proximity to death (Chai et al., 2014). A cost analysis conducted in 2008 evaluated the costs of a pilot interdisciplinary healthcare model of home-based palliative care from a health system perspective (Johnson et al., 2009). Total costs for 434 patients enrolled in the pilot programme were $2.4 million, equating to a cost per patient of $5,586.33 (Johnson et al., 2009).

An evaluation of resource utilisation and costs of an enhanced end of life shared-care project delivered to 95 patients (87% cancer patients; the remaining 13% mainly COPD and heart disease patients) reported total costs of $1,625,658.07, equating to $17,112.19 per patient or $117.95 per patient day (Klinger et al., 2013). Although these costs were higher than previously reported expenditures for cancer patients, the authors concluded that costs remained proportionate to funding allocated to long-term care homes and less than alternate level of care and hospital costs in Canada (Klinger et al., 2013).

##### 3.3.1.4 Israel

A cost analysis conducted in Israel compared resource utilisation and costs of health services in the last six months with and without home-hospice care among a sample of n=429 patients with metastatic cancer (Bentur et al., 2014). Over the last six months of life, the average cost of care for patients who had received home-hospice care was US$13,648, whereas the average cost of care over the last six months for patients who did not receive home hospice care was significantly higher at a cost of US$18,503 (Bentur et al., 2014).

##### 3.3.1.5 Belgium

In Belgium, a cost analysis assessed the quality of care and costs of palliative home care support in the last 720 to 15 days of life (Maetens et al., 2019). The analysis was conducted from a third party and patient co-payment perspective and included inpatient and outpatient cost with a matched comparison group. People (n = 8837) who received palliative home care support in the last 720 to 15 days of life were matched 1:1 by propensity score to 8837 people who received usual care. The provision of home-based palliative care was associated with more physician contacts, an increased chance of a home death and a lower chance of hospital admissions, compared with no palliative home care support. Moreover, cost findings revealed lower mean total costs of care for those who had received home-based palliative care support (€3,081 [95% CI €3,025 to €3,136] vs €4,698 [95% CI €4,610 to €4,787]; incremental cost: −€1,617 [p<0.001]) (Maetens et al., 2019).

#### 3.3.2 Cost analysis of a residential care home model of end of life care

One cost analysis study assessed the costs of community end of life care of older people with dementia in residential care homes with no on-site nursing in England (Phase 1) and then evaluated the costs of an intervention designed to improve end of life care in this setting (Phase 2) (Amador et al., 2014). Findings from Phase 1 revealed that monthly costs per resident were £2,800. The intervention implemented in Phase 2 of the study resulted in a 43% reduction in total service costs and an 88% reduction in hospital care costs (Amador et al., 2014). Further information on study characteristics and main findings of this study can be viewed in Section 6 of this report.

#### 3.3.3 Cost analysis of a community-based paediatric palliative care model

A cost analysis study conducted in the USA used administrative data to calculate hospital utilisation and costs associated with a community-based paediatric palliative care model that involved engaging children and families in their homes, schools, and communities to provide a holistic range of services across primary, secondary and community settings (Goldhagen et al., 2016). The community-based paediatric care model was associated with positive findings in relation to health-related quality of life (HRQoL), was generally high, and hospital charges per child declined by $1,203 for total hospital services (P = .34) and $1,047 for diagnostic charges per quarter (p=0.13). Moreover, there was a decrease in length of stay at hospital from 2.92 days per quarter to 1.22 days per quarter (P < .05) (Goldhagen et al., 2016). Further information on study characteristics and main findings of this study can be viewed in Section 6 of this report.

#### 3.3.4 Bottom line results for costs of community-based models of palliative care

Findings from the eleven cost analyses of home-based models of palliative and end of life care reported either positive (n=6) or neutral findings (n=5) in relation to costs. Informal care makes up a significant financial contribution to models of home-based palliative care (Butler et al., 2022; Chai et al., 2014). Moreover, a residential care home model of end of life care (Amador et al., 2014) and a community-based model of paediatric palliative care (Goldhagen et al., 2016) resulted in fewer healthcare costs.

#### 3.3.5 Economic evaluations of community-based models of palliative care

Two economic evaluations assessed the cost-effectiveness of home-based models of palliative and end of life care (Pham & Krahn, 2014; Rosato et al., 2021). In Italy, a cost-effectiveness and cost-utility analysis of a home-based palliative care approach for people living with severe multiple sclerosis (MS) compared with usual care was conducted (Rosato et al., 2021). The mean baseline-adjusted cost difference was € −394 (95% confidence interval, CI −3,532 to 2,743). The cost-effectiveness analysis considering Palliative Outcome Scale-Symptoms-MS scored revealed no change in costs. In summary, home-based palliative care did not impact QALYs and only produced slightly improved symptom scores with no increase in associated costs (Rosato et al., 2021).

A cost-effectiveness of end of life intervention study identified in the ‘end of life mega analysis’ was conducted from the Canadian health payer’s perspective (Pham & Krahn, 2014). The analysis used multiple data sources, including systematic reviews, linked health administration data, and survey data. Results from the primary cost-effectiveness analysis indicated that in-home palliative care decreased per patient costs by $4,400 and increased the likelihood of dying at home rather than in hospital by 10%, increased the average number of patient days at home (6 days) and quality-adjusted life-days (0.5 days) (Pham & Krahn, 2014). Tables 9-20 in Section 6 of this report provide further information on the study characteristics and main findings of the two economic evaluations of community-based models of palliative and end-of life care. Quality appraisal of the community-based studies can be viewed in Appendix 9.2. See Table 4 for a summary of the cost-effectiveness study relating to home/community palliative care.

**Table 4.** Summary of cost-effectiveness studies relating to home/community palliative care.

| Study | Cost | QALYs | Per-Day Variable Cost (mean [SD]) |  | Results |
| --- | --- | --- | --- | --- | --- |
|  |  |  | Cost | QALYs |  |
| **(Pham & Krahn, 2014)**<br>**Canada** | **Mean total cost (SD)** |  |  |  |  |
| Comprehensive palliative team care | \$CAN2,109 (\$CAN1,160) | Not reported | 72,717 | | 32% likely to be cost-effective at a threshold of \$CAN50,000 per QALY.<br>Not dominant |
| Usual care | \$CAN50,129 | Not reported | Not reported | Not reported | |
| **(Rosato et al., 2021)** | **Average cost per patient** |  |  |  |  |
| Home-based palliative approach (HPA) for people with severe multiple sclerosis (MS). | €2,217.3 (€1,874.2) | Not reported | Not reported | 0.07 | No cost difference between home-based palliative approach and usual care.<br>Not dominant |
| Usual care | €1,909.3 (€1,852.7) |  | Not reported | 0.08 | Not dominant |

See Table 5 for the table of inflated and converted community-based palliative care costs. (Caution should be taken when reading this table as the time horizons are different in nearly every row). The list of inflation calculators used for inflating and converting costs from the original currency into GBP is presented in Appendix 9.3.

**Table 5.** Home/Community-based palliative care costs table.

| Citation | Cost year | Description of cost | Original currency | Currency in 2023 | GBP in 2023 |
| --- | --- | --- | --- | --- | --- |
| Amador et al., 2014 | 2010 | Cost of palliative care per month | £2,800 | £4,133 | £4,133 |
| Bentur et al., 2014 | 2010 | Palliative care costs over the last 6 months of life – with Home Hospice Unit (HHU) | US\$13,648 | US\$19,257 | £15,291 |
| Bentur et al., 2014 | 2010 | Palliative care costs over the last 6 months of life – without Home Hospice Unit (HHU) | US\$18,503 | US\$26,107 | £20,731 |
| Butler et al., 2022 | 2019 | Median cost of informal palliative care costs 0-14 days before death | £580 | £710 | £710 |
| Chai et al., 2014 | 2011 | Mean cost of palliative care over the last 12 months of life. | CAN\$14,924 | CAN\$19,594 | £11,446 |
| Chen et al., 2018 | 2014 | The mean saving to Medicare expenditures of an established palliative care homebound programme. | US\$18,251 | US\$23,720 | £18,835 |
| Enguidanos et al., 2002 | 2005 | Cost savings for patients with COPD enrolled in a palliative care group. | US\$11,325 | US\$17,841 | £14,165 |
| Enguidanos et al., 2002 | 2005 | Cost savings for patients with CHF enrolled in a palliative care group. | US\$8,445 | US\$13,304 | £10,563 |
| Gage et al., 2015 | 2010 | 11 Rapid Response Services (RRS) visits | £425 | £627 | £627 |
| Goldhagen et al., 2016 | 2016 | Cost saving of hospital services per child in the Community Based Paediatric Palliative Care (CBPPC) Service | US\$1,203 | US\$1,542 | £1,224 |
| Gordon et al., 2021 | 2019 | Gross savings when enrolled in the home palliative care (HPC) programme for 6-12 months. | US\$26,409 | US\$31,782 | £25,217 |
| Johnson et al., 2009 | 2007 | Cost of palliative home care for two months in Canada. | CAN\$5586.33 | CAN\$793,900 | £463,830 |
| Klinger et al., 2012 | 2007 | Cost per patient for all patient related palliative care services in rural Ontario. | CAN\$17,112 | CAN\$24,319 | £14,206 |
| Maetens et al., 2019 | 2017 | Palliative care - home support cost | €3,081 | €3,984 | £3,412 |
| Maetens et al., 2019 | 2017 | Cost without home palliative care | €4,698 | €6,076 | £5,204 |
| Pham and Krahn, 2014 Cost-effectiveness | 2013 | The per-patient cost of providing in-home palliative team care. | CAN\$1,700 - CAN\$2,400. | CAN\$2,192 - CAN\$3,095. | £1,281 - £1,808 |
| Rosato et al., 2021 Cost-effectiveness | 2017 | The per-patient cost of a home-based palliative approach (HPA) for people with severe multiple sclerosis. | €23,195 | €29,997 | £17,525 |
| Spiro et al., 2020 | 2020 | Cost of palliative care by the Hospice at Home (HathH) Service per person over an average of 11 days. | £673 | £883 | £883 |

#### 3.3.6 Bottom line results for the cost-effectiveness of community-based models of palliative and end of life care

There is a dearth of evidence on the cost-effectiveness of community-based models of palliative and end of life care. However, results from one cost-effectiveness analysis indicated that in-home palliative care in Canada was cost-effective compared to hospital care (Pham & Krahn, 2014). A home-based palliative care approach in Italy for people with severe MS was similar to usual care in terms of cost-effectiveness (Rosato et al., 2021).

### 3.4 Models of palliative care which are not hospital, hospice or home-based, but a combination of pathways at various times

Of the n=22 combination models of palliative care papers, n=4 included outcomes relating to hospital, hospice, and home or community-based palliative and end of life care (Bjørnelv et al., 2020; Brick et al., 2017; Rolden et al., 2014; Yi et al., 2020); n=2 included cost outcomes for hospital and hospice (Hoverman et al., 2020; Saygili & Çelik, 2019); n=13 included cost outcomes related to hospital and home (Comans et al., 2021; Duncan et al., 2019; Emmert et al., 2013; Kalluri et al., 2020; Kato & Fukuda, 2017; Lustbader et al., 2017; McBride et al., 2011; Seow et al., 2022; Spilsbury & Rosenwax, 2017; Tanuseputro et al., 2015; Terada et al., 2018; Urban et al., 2018; Yu et al., 2015), n=1 related to hospice and home costs (Kim et al., 2022), and n=2 related to enhanced supported care services such as ESC and ACP (Monnery et al., 2023; Nguyen et al., 2017). In terms of quality, all (n=22) were of high quality.

#### 3.4.1 Combination models of care: Hospital, Hospice and Home

Four studies focused on combination models of palliative care encompassing hospital, hospice, and home/community costs. These studies were published between 2014 and 2020 and were from Ireland, Norway, the Netherlands (Bjørnelv et al., 2020; Brick et al., 2017; Rolden et al., 2014) and England, Ireland and the USA (Yi et al., 2020). Three of the studies were from a healthcare system perspective, and one was from a health insurer perspective (Rolden et al., 2014). See Section 6 of this report for more details.

In Ireland, the mean total formal palliative care costs (calculated over the total sample of decedents, n=215) in the last year of life did not vary significantly across three areas of Ireland (P > 0.136). The healthcare end of life costs ranged from €50,071 in the Midlands, to €50,036 in the Mid-West, to €40,137 (Table 3) in the South East (2011 prices). Informal care was valued as the replacement cost of care (Brick et al., 2017). In 2014, in the Netherlands, the average cost of dying was €25,919 in 2014. The authors of this study included all deceased subjects for whom healthcare expenses were known for 26 months prior to death. Costs of dying were defined as healthcare expenses made in the last six months before death (Rolden et al., 2014).

End of life costs in the last three months of life was calculated in a 2020 study comparing costs in England, Ireland, and the USA. Mean care costs per person with cancer/non-cancer were US$37,250/US$37,376 (the United States), US$29,065/US$29,411 (Ireland), US$15,347/ US$16,631 (England) and differed significantly (F = 25.79/14.27, p < 0.000). In all countries, hospital care accounted for > 80% of total care costs; community care 6%– 16%, palliative care 1%–15%; 10% of decedents used ∼30% of total care costs. Being a high-cost user was associated with older age (>80 years), facing financial difficulties and poor experiences of home care, but not with having cancer or multimorbidity. Results were similar in the sensitivity analyses using the same unit costs for all countries. Hospital costs were 79%–88% of total care costs. (Yi et al., 2020). Also, in 2020, a study from Norway investigated the healthcare costs of persons now deceased. They estimated that the costs for the last six months of life were NOK 400,000 (approximately £31,734 in 2023) (Bjørnelv et al., 2020).

#### 3.4.2 Combination models of care: Hospital and Hospice

Two papers were included that described combination models of care, including hospital and hospice care between 2019 and 2020. One study was from the USA (Hoverman et al., 2020), and the other was from Turkey (Saygili & Çelik, 2019). One study was from a healthcare system perspective (Hoverman et al., 2020), and the other was from a societal perspective (Saygili & Çelik, 2019).

A cost-effectiveness study from Turkey looked at the cost of palliative care services from a societal perspective. From a patient perspective, home healthcare services (HHC) were found to be more cost-effective compared to the other two models. The average indirect cost ($164.10) for the patients receiving care from Hospital Inpatient Services (HISs) was found to be the lowest compared with the indirect costs of HHCs ($344.62) and comprehensive palliative care centre (CPCCs) ($778.43). Estimated incremental cost-effectiveness ratios (ICERs) indicated that HHC was more likely to produce a better quality of life at the cost of an additional $33.43 per additional one quality of life (QoL) score when it was compared with HIS. However, HHC has the capability of producing a better QoL score and even reduces indirect costs ($18.30) for patients with an additional QoL score compared with CPCC. The hospital inpatient service model was found to be more cost-effective than the CPCC model (Saygili & Çelik, 2019).

The aim of the Medicare cost analysis study from the USA was to measure and characterise the total cost of care for those who received less than three days of hospice care (HC) at the end of life compared with those who received three days or more. It was found that dying in hospital was twice the cost of dying at home ($20,113 vs. $10,803). The average final 30-day spend was $22,410 if the death was at hospital, but more if a person died in the intensive care unit (ICU) at $28,301. Dying at a Skilled Nursing Facility (SNF) cost $19,400, dying in a medical hospice cost $17,418, and dying with hospice at home support cost $10,098 in 2020 (Hoverman et al., 2020).

#### 3.4.3 Combination models of care: Hospital and Home

Thirteen of the included papers estimated the cost of hospital and home palliative care. All of the studies were cost analyses, and they were from Australia (Comans et al., 2021; Spilsbury & Rosenwax, 2017), Canada (Kalluri et al., 2020; Seow et al., 2022; Tanuseputro et al., 2015; Yu et al., 2015), England (McBride et al., 2011), Germany (Emmert et al., 2013), Japan (Kato & Fukuda, 2017; Terada et al., 2018), and the USA (Duncan et al., 2019; Lustbader et al., 2017; Urban et al., 2018). Nine of the studies were from a healthcare system perspective, one was from a payer perspective (Kalluri et al., 2020), one was from an insurer perspective (Lustbader et al., 2017), and one was from a societal perspective (McBride et al., 2011). These studies will be described below.

##### 3.4.3.1 Australian setting

In 2017, an Australian cost analysis study found that community-based specialist palliative care was associated with a reduction of inpatient averaged hospital costs. The cohort included 12,764 decedents who, combined, spent 451,236 (9.7%) days of the last year of life in hospital. Overall, periods of time receiving community-based specialist palliative care were associated with a 27% decrease from A$112 (A$110-A$114) per decedent per day to $A82 (A$78-A$85) per decedent per day of CA hospital costs. Community-based specialist palliative care was also associated with a reduction of inpatient averaged hospital costs of 9% (7%-10%) to A$1030 per hospitalised decedent per day. Hospital cost reductions were observed for decedents with organ failures, chronic obstructive pulmonary disease, Alzheimer’s disease, Parkinson’s disease, and cancer, but not for motor neurone disease.

Cost reductions associated with community-based specialist palliative care were evident four months before death for decedents with cancer and by one to two months before death for decedents dying from other conditions.(Spilsbury & Rosenwax, 2017).

In contrast, a more recent Australian cost analysis study found that a palliative care bed in hospital cost less than a bed in an end of life palliative care facility in a modified unit in a Residential Aged Care Facility (RACF). An additional $120 per day is required to provide the higher level of care required by people with complex palliative care needs in a modified RACF unit. QoL and utility of the participants were measured at baseline, end of programme, and three and six months post baseline using the EQ-5D and ICECAP-O (Comans et al., 2021).

##### 3.4.3.2 USA setting

Three studies which focused on hospital and home palliative care were from the USA (Duncan et al., 2019; Lustbader et al., 2017; Urban et al., 2018). In 2017, a retrospective study found that the cost per patient of end of life care in the last three months of life was $12,000 lower with home based palliative care (HBPC) than with usual care ($20,420 vs. $32,420; p = 0.0002); largely driven by a 35% reduction in Medicare Part A ($16,892 vs. $26,171; p = 0.0037). HBPC also resulted in a 37% reduction in Medicare Part B in the final three months of life compared to usual care ($3,114 vs. $4,913; p = 0.0008). Hospital admissions were reduced by 34% in the final month of life for patients enrolled in HBPC. The number of admissions per 1,000 beneficiaries per year was 3,073 with HBPC and 4,640 with usual care (p = 0.0221). HBPC resulted in a 35% increased hospice enrolment rate (p = 0.0005) and a 240% increased median hospice length of stay compared to usual care (34 days vs. 10 days; p < 0.0001) (Lustbader et al., 2017). Another Medicare cost analysis study from 2019 also presented evidence that more effective use of palliative care and hospices offers a lower cost, higher quality alternative for patients at end of life (Duncan et al., 2019). A further study from the USA utilised the linked Surveillance, Epidemiology and End Results (SEER)-Medicare database and identified a cohort of women with stage III/IV epithelial ovarian cancer diagnosed between 1995 and 2007. The authors defined the end of life as the last 90 days prior to death. They concluded that reducing the prescription of chemotherapy and increasing the use of hospice services for ovarian cancer patients is a way of reducing costs at the end of life (Urban et al., 2018).

##### 3.4.3.3 German setting

In Germany, a cost analysis study was conducted to estimate the costs of palliative care for colorectal cancer (CRC) from the perspective of German statutory health insurance and to measure the patients’ quality of life (QoL) for a 2-year period. The mean costs per patient during the first and second years were calculated to be €42,361 and €32,023, respectively. Highest mean costs were calculated for the second quarter, which reached an amount of €12,900 (95 % CI: €11,127–€14,673)(Emmert et al., 2013).

##### 3.4.3.4 Canadian setting

Four of the included papers were from Canada (Kalluri et al., 2020; Seow et al., 2022; Tanuseputro et al., 2015; Yu et al., 2015). The aim of one of the 2015 Canadian cost analysis studies was to examine healthcare use and cost in the last year of life. Among 264,755 decedents, the average healthcare cost in the last year of life was $53,661 (Quartile 1–Quartile 3: $19,568–$66,875). The total captured annual cost of $4.7 billion represents approximately 10% of all government-funded healthcare. Inpatient care, incurred by 75% of decedents, contributed 42.9% of total costs ($30,872 per user). Physician services, medications/devices, laboratories, and emergency departments combined to less than 20% of total cost. About one quarter used long-term care, and 60% used home care ($34,381 and $7,347 per user, respectively). Costs rose in the last 120 days prior to death, predominantly for inpatient care (Tanuseputro et al., 2015).

Another Canadian cost analysis study published in 2015 found that the estimated total societal cost of end of life care was $34,197.73 per patient over the entire palliative trajectory (four months on average). Results showed no significant difference (P > 0.05) in total societal costs between home and hospital deaths. Higher hospitalisation costs for hospital deaths were replaced by higher unpaid caregiver time and outpatient service costs for home death patients. Thus, from a societal cost perspective, alternative sites of death, while not associated with a significant change in the total societal cost of end of life care, resulted in changes in the distribution of costs borne by different stakeholders such as carers (Yu et al., 2015).

The aim of a Canadian administrative health data study published in 2020 was to evaluate the differences in resource use and associated costs of end of life care between patients with idiopathic pulmonary fibrosis (IPF) who received early integrated palliative care and patients with IPF who received conventional treatment. Multidisciplinary collaborative (MDC) patients were less likely to die in the hospital (44.9% MDC vs. 64.9% Specialised Care (SC) vs. 66.8% NSC; P,0.001) and had the highest rates of no hospitalisation in the last year of life. The median total healthcare costs in the last three months of life were approximately $CAN7,700 lower for MDC patients than for those receiving SC, driven primarily by fewer hospitalisations and emergency department visits. MDC patients were also less likely to die in the hospital (44.9% MDC vs. 64.9% SC vs. 66.8% Non-Specialist Care (NSC); P,0.001) and had the highest rates of no hospitalisation in the last year of life. (Kalluri et al., 2020).

A 2022 Canadian publication aimed to investigate the impact of early versus not-early palliative care among cancer decedents on end of life healthcare costs. In the early-palliative care group, 56.3% used inpatient care in the last month compared with 66.7% of control group (P < .001), which resulted in a statistically different average inpatient hospital costs: $7,105 (CAN$10,710) in the early group versus $9,370 (CAN$13,685) in the hard matched (on age, sex, cancer type, and stage at diagnosis) control group (P < .001). The average overall health system costs per patient in the early-palliative care group versus control group was $12,753 (CAN$10,868) versus $14,147 (CAN$14,288; P, .001) in the last month of life. The sensitivity analyses looked at early versus late paired groups and showed the same statistically significant trends as the main and sub-analysis. However, in the early versus never paired groups, the never users had lower overall costs, although this was not statistically significant (Seow et al., 2022).

##### 3.4.3.5 Japanese setting

Two cost analysis papers were from the Japanese setting (Kato & Fukuda, 2017; Terada et al., 2018). The aim of the 2017 Japanese study was to quantify the difference between adjusted costs for home-based palliative care and hospital-based palliative care in terminally ill cancer patients. Home care was significantly associated with a reduction of $7,523 (95% CI $7,093–$7,991, P = 0.015) in treatment costs. The cost data was collected through insurance claims and medical records (Kato & Fukuda, 2017). The aim of the 2018 Japanese study was to evaluate the costs associated with healthcare and long-term care costs according to public insurance schemes during the last 24 months before death among all decedents older than 75 years, according to major disease groups. For the 2,149 decedents studied, the average healthcare costs per capita in the last 24 months of life for moderately old (75 to 84 years) and extremely old (85 years and older) decedents was 4,135,467 Japanese Yen (JPY) and 2,493,001 JPY, respectively, while the average long-term care costs per capita for 24 months was 1,300,710 JPY and 2,723,239 JPY, respectively. The total costs (healthcare and long-term care combined) ranged from 9,169,547 JPY for chronic kidney disease to 5,023,762 JPY for ischemic heart disease. In all the diseases studied, the moderately old decedents incurred higher healthcare costs, while the extremely old decedents incurred higher long-term care costs (Terada et al., 2018).

##### 3.4.3.6 UK setting

One Markov modelling study from England, UK found that the cost to the taxpayer of providing care in the last year of life, based on 127,000 patients who died of cancer in 2006, was approximately £1.8 billion or £14,236 per patient in 2006 currency. Costs were based on daily costs of care in hospital, costs of community care, and ambulance journey costs (McBride et al., 2011).

#### 3.4.4 Combination models of care: Hospice and Home

One Markov modelling study from Korea focussed on hospice and home palliative care from a healthcare system perspective. The incremental cost-effectiveness ratio (ICER) of the home-start group was 796,476 Korean won/quality-adjusted life week (KRW/QALW). Based on one-way sensitivity analyses, the ICER was predicted to increase to 1,626,988 KRW/QALW if the weekly cost of home-based hospice doubled, but it was estimated to decrease to −2,898,361 KRW/QALW if death rates at home doubled (Kim et al., 2022).

#### 3.4.5 Combination models of care: Enhanced supported care services (ESC services)

Two studies were regarding enhanced supported palliative care services (Monnery et al., 2023; Nguyen et al., 2017). One was study was from a healthcare system perspective in Australia (Nguyen et al., 2017) and the other was from a societal perspective in England (Monnery et al., 2023).

An Australian Markov modelling study indicated that if the cost per individual of advanced care planning (ACP) reached $850 (equivalent to seven visits), then the programme is no longer cost-effective. Nonetheless, this scenario is unlikely because individual ACP has been provided successfully in Australia through group information sessions followed by 1–2 visits by trained ACP facilitators, which costs less than $250. At this cost, the programme is more likely to be cost-effective than the base case scenario. Extensive sensitivity analyses, including threshold analyses, were conducted on the key parameters to assess the likelihood of ACP remaining cost-effective. The result was highly sensitive to several key parameters: ACP completion and compliance rates and dying choice (hospital versus non-hospital settings). The probability sensitivity analysis of 5000 Monte Carlo replications highlighted that there was a 50–50 chance that a nationwide ACP programme would be cost-effective (see Fig. 2) due to high uncertainty around the key parameters (Nguyen et al., 2017).

A cost analysis study from England published in 2023 investigated enhanced supported care (ESC) across eight cancer centres in England. ESC service design and costs were recorded. Data relating to patients’ symptom burden were collected using the Integrated Palliative Care Outcome Scale (IPOS). For patients in the last year of life, secondary care use was compared against an NHS England published benchmark. In total, £1,676,044 was spent delivering ESC across the eight centres. Reductions in secondary care usage for the 1,061 patients who died saved a total of £8,490,581 for the UK health system. ESC services appear to be effective at supporting people dying of cancer and significantly reduce the costs of their care (Monnery et al., 2023). Quality appraisal combined models of care studies can be viewed in Appendix 9.2.

See Table 6 for the summary of the cost-effectiveness study relating to combined models of palliative care.

**Table 6.** Summary of cost-effectiveness studies relating to combined models of palliative care.

| Study | Cost | QALYs | Per-Day Variable Cost (mean [SD]) |  | ICER |
| --- | --- | --- | --- | --- | --- |
|  |  |  | Cost | QALYs |  |
| **(Saygili & Çelik, 2019)** | **Mean direct cost (SD)** |  |  |  |  |
| Comprehensive Palliative Care Centre (CPCC) | US\$4,776 | Not reported | Not reported | Not reported | -33.57 (dominant) |
| Hospital inpatient services (usual care) | US\$2,385 | Not reported | Not reported | Not reported | |

See Table 7 for the table of inflated and converted community-based palliative care costs.

**Table 7.** Combination models of palliative care costs table.

| Citation | Cost year | Description of cost | Original currency | Currency in 2023 | GBP in 2023 |
| --- | --- | --- | --- | --- | --- |
| (Bjørnelv et al., 2020) | 2020 | The average patient cost (four months at home, 27 days in long-term institutions, 16 days in short-term institutions, and 21 days in the hospital). | NOK 400,000 | NOK 437,795 | £31,734 |
| (Brick et al., 2017) | 2011 | Last year of life costs range | From €40,137-€50,071 | €55,234-€68,905 | £47,306-£59,014 |
| (Comans et al., 2021) | 2021 | The palliative care hospital bed cost per day | AUS\$1,664 | AUS\$1,774 | £926 |
| (Duncan et al., 2019) | 2017 | 1-3 days prior to death palliative care cost: Hospital | US\$5,983 | US\$7,510 | £5,963 |
| (Duncan et al., 2019) | 2017 | 1-3 days prior to death palliative care cost: Hospice | US\$231 | US\$290 | £230 |
| (Hoverman et al., 2020) | 2020 | Average final 30-day spend for end of life care: Hospital | US\$22,410 | US\$26,640 | £21,159 |
| (Hoverman et al., 2020) | 2020 | Average final 30-day spend for end of life care: Hospice | US\$17,418 | US\$20,706 | £16,446 |
| (Hoverman et al., 2020) | 2020 | Average final 30-day spend for end of life care: Hospice at Home | US\$10,098 | US\$12,004 | £9,534 |
| (Kalluri et al., 2020) | 2017 | The median total healthcare costs in the last 3 months of life. | CAN\$7,700 | CAN\$9,329 | £5,457 |
| (Kato & Fukuda, 2017) | 2014 | Cost saving due to home based palliative care versus hospital care. | US\$7,523 | US\$7,777 | £7,768 |
| (Kim et al., 2022) | 2022 | Weekly cost per patient in a hospice | KW 2,481,479 | KW 2,584,227 | £1,560 |
| (Kim et al., 2022) | 2022 | Weekly cost per patient with home-based palliative care | KW 225,688 | KW 235,033 | £142 |
| (Nguyen et al., 2017) | 2015 | Usual care (for the model) | AUS\$7,250 | AUS\$8,571 | £4,483 |
| (Nguyen et al., 2017) | 2015 | Advanced Care Planning (ACP) (for the model) | AUS\$7,160 | AUS\$8,465 | £4,428 |
| (Lustbader et al., 2017) | 2017 | Cost per patient during the final three months of life – Home based palliative care (HBPC) | US\$20,420 | US\$255,631 | £20,359 |
| (Lustbader et al., 2017) | 2017 | Cost per patient during the final three months of life – Usual care | US\$32,420 | \$US40,693 | £32,323 |
| (McBride et al., 2011) | 2011 | Cost of the last year of life. | £14,236 | £23,510 | £23,510 |
| (Monnery et al., 2023) | 2022 | Cost saving after delivery of Enhanced Supported Care (ESC) across the eight centres. | £8,490,581 | £9,209,748 | £9,209,748 |
| (Rolden et al., 2014) | 2014 | Care costs in the last six months before death. | €25,919 | €34,189 | £29,276 |
| (Saygili & Çelik, 2019)<br>Cost-effectiveness | 2016 | Mean direct cost of hospital inpatient services | US\$2,385 | US\$3,057 | £2,427 |
| (Saygili & Çelik, 2019)<br>Cost-effectiveness | 2016 | Mean direct cost of Comprehensive Palliative Care Centre (CPCC) | US\$4,776 | US\$6122 | £4,860 |
| (Seow et al., 2022) | 2022 | Early palliative care in the last month of life | CAN\$12,753 | CAN\$13,151 | £7,695 |
| (Seow et al., 2022) | 2022 | Usual care in the last month of life | CAN\$14,288 | CAN\$14,734 | £8,621 |
| (Spilsbury & Rosenwax, 2017) | 2017 | Reduction of inpatient averaged hospital costs per day due to Community-based specialist palliative care, | AUS\$1030 | AUS\$1179 | £616 |
| (Tanuseputro et al., 2015) | 2013 | Average healthcare cost in the last year of life. | CAN\$53,661 | CAN\$69,192 | £40,479 |
| (Terada et al., 2018) | 2018 | Average healthcare costs per capita in the last 24 months of life for moderately old (75 to 84 years) people. | 4,135,467 JPY | 4,400,642 JPY | £23,737 |
| (Urban et al., 2018) | 2009 | The mean total payment per patient in the last 90 days of life | US\$24,073 | US\$28,617 | £22,708 |
| (Yi et al., 2020) | 2020 | Mean care costs for patients with cancer in the USA. | US\$37,250 | US\$44,282 | £35,138 |
| (Yi et al, 2020) | 2020 | Mean care costs for patients without cancer in the USA | US\$37,376 | US\$44,432 | £35,257 |
| (Yi et al, 2020) | 2020 | Mean care costs for patients with cancer in Ireland. | US\$29,065 | US\$34,552 | £27,417 |
| (Yi et al, 2020) | 2020 | Mean care costs for patients without cancer in the Ireland | US\$29,411 | US\$34,963 | £27,743 |
| (Yi et al, 2020) | 2020 | Mean care costs for patients with cancer in England. | US\$15,347 | US\$18,244 | £18,244 |
| (Yi et al, 2020) | 2020 | Mean care costs for patients without cancer in England. | US\$16,631 | US\$19,771 | £15,688 |
| (Yu et al., 2015) | 2012 | The estimated total societal cost of end of life care was per patient over the entire palliative trajectory (4 months on average). | CAN\$34,197 | CAN\$44,383 | £25,995 |

#### 3.4.6 Bottom line results for models of care with combined costs

There are many models of palliative care defined as any structured care model involving multiple components, including setting. Hospital care costs tend to be higher than the costs of hospice end of life care, and home-based palliative care is the least costly choice in many studies (Duncan et al., 2019; Saygili & Çelik, 2019; Yi et al., 2020).

## 4. DISCUSSION

This rapid review aimed to determine the costs and outcomes of different service models of palliative care or end of life care in OECD countries. In this review, the term ‘model of palliative care’ was defined as any structured care model involving multiple components, including ‘who delivers (e.g., professionals, paid carers) the intervention (specialist or generalist palliative care), where (setting e.g., hospital), to whom (care recipients), when (i.e. timing and duration), how (e.g., face to face) and for what purpose (i.e. expected outcomes) (Brereton et al., 2017; Davidson et al., 2006). Much of the focus of this RR was on the setting (hospital, hospice, home/community setting or a combination of settings).

Hospital care costs account for most of the costs of dying, and this has been reported to be 80% or more of the costs of death for some people (Duncan et al., 2019; Saygili & Çelik, 2019; Yi et al., 2020). Other authors have also reported on the high cost of dying in hospital, especially for cancer patients (Kerr et al., 2017; Tan & Jatoi, 2011). Most of the hospital-based palliative care studies found that hospital based palliative care costs increased significantly in the last thirty days of life. This is due to increased hospitalisation costs as the condition of the patients deteriorates (Pollock et al., 2022; Tan & Jatoi, 2011). However, there was some evidence to indicate that if a palliative consultation was done prior to death, the hospitalisation costs were less as palliative care consultation is followed by decisions to forego costly treatment, resulting in greater cost-savings (Hanson et al., 2008; Isenberg et al., 2017; McCarthy et al., 2015). A cost-effectiveness study conducted in Australia found that Advanced Care Planning (ACP) is more costly but more effective in facilitating adherence to patient preferences for end of life care (Sellars et al., 2019). A cost-effectiveness study from the USA found that a hospital based palliative care unit (PCU) produced cost savings and was profitable for the hospital compared to usual hospital end of life care at the hospital (Isenberg et al., 2017).

There is limited evidence for hospice care costs, and the data is not comparable across different countries and health systems currently as the times of measurements and what is measured differs a great deal (Duncan et al., 2019; Huskamp et al., 2008). This rapid review presents evidence that palliative care is more costly on the first and last days of end of life care in hospices (Comans et al., 2021).

Findings from eleven cost analyses of home-based models of palliative and end of life care reported either positive (n=6) or neutral findings (n=5) in relation to costs. Informal care makes up significant financial contribution to models of home-based palliative care (Butler et al., 2022; Chai et al., 2014). Additionally, a residential care home model of end of life care reduced healthcare costs (Amador et al., 2014).

There is a dearth of evidence on the cost-effectiveness of community-based models of palliative and end of life care. However, one cost-effectiveness analysis indicated that in-home palliative care in Canada was cost-effective and reduced costs by approximately $4,400 per patient. It also increased the chance of dying at home by 10%(Pham & Krahn, 2014). A home-based palliative care approach in Italy for people with severe MS was similar to usual care in terms of costs. The slight reduction of symptom burden produced by home-based palliative care was not associated with increased costs. The National Healthcare System and people with severe MS almost equally sustained these costs (Rosato et al., 2021).

Most of the included papers were cost analyses based on observational evidence as there are additional challenges when conducting traditional economic evaluation methods, such as cost-effectiveness studies that use quality-adjusted life years (QALY’s) as an outcome measure. Improving QALYs may not be the intended aim of palliative care or end-of life care interventions (Pham & Krahn, 2014).

There were only two studies relating to the costs of paediatric palliative care models (Gans et al., 2016; Mitchell et al., 2020). None of the studies included in our review provided evidence on the costs or cost-effectiveness of neonatal palliative care. This is a notable gap in the evidence. However, the authors are aware that a large NIHR funded study to establish children’s palliative care models is underway but has yet to be completed. This work covers Wales (Bedendo et al., 2023; Papworth et al., 2023).

The quality and applicability of the evidence we found in our rapid review were variable, and therefore, uncertainty remains, especially when the perspective of analysis was not stated clearly (Perea-Bello et al., 2023). Therefore, it was difficult to ascertain whether all relevant costs were considered. Assumptions on costs were not varied in many studies, and most studies had different time horizons.

Models of palliative care for patients with cancer, CKD, and COPD were well represented in the evidence base identified in our rapid review. However, other conditions were not well represented in the RR, indicating a gap in the evidence base.

### 4.1 Summary of the findings

Cost analyses were found for hospital, hospice, home-based and community-based palliative care models as well as primary care focussed models of palliative care and mixed models of care.

Generally, hospital palliative care is the most costly with a range of costs between £10,000 and £64,000 per hospital death in 2023 prices (Kerr et al., 2017; Sellars et al., 2019).

Hospice end of life care costs ranged between £2,000 and £16,000 in 2023 prices, according to the evidence presented in our review (Huskamp et al., 2008).

Only one study looked at the cost of paediatric palliative care, which ranged from £13,000 to £16,000 per death in 2023 prices (Gans et al., 2016).

In terms of home and community based palliative care costs for end of life care ranged from £900 - £21,000 in 2023 prices (Bentur et al., 2014; Spiro et al., 2020).

In terms of the combined models of palliative care studies, the costs of end of life care ranged from £5,000 to £41,000 in 2023 prices (Kalluri et al., 2020; Tanuseputro et al., 2015).

### 4.2 Strengths and limitations of the available evidence

The available economic evidence of palliative care is focused on costs of hospital care and less so on hospice care. Combination models of care are commonplace, and there is emerging evidence of Enhanced Supported Care (ESC), which seems to be cost saving to the NHS in the UK (Monnery et al., 2023). More studies on this type of palliative care service are needed. Assessing the cost-effectiveness of palliative and end of life care models is challenging as they often do not meet the criteria required for conducting economic evaluations. There was a lack of information about the economic perspective adopted in many of the included studies. This lack of clarity makes it difficult to determine whether all relevant costs (and outcomes in the included economic evaluations) were included in the analysis. Many of the studies were retrospective studies of decedents and not interventional studies or RCTs.

### 4.3 Strengths and limitations of this Rapid Review

The main strength of this rapid review is that it identified 40 relevant cost studies, and five cost-effectiveness studies, three Markov modelling studies as well as eight relevant systematic reviews (See Table 22). The main limitation was that the evidence focused on different time periods as well as various aspects of costs of services, making interpretation difficult. In this review, we focused mainly on cost analyses, as there were few cost-effectiveness papers comparing one type of palliative care model with another. We did not consider the quality of death or the burden imposed on caregivers as outcomes.

This RR used the JBI critical appraisal checklist for economic evaluations to appraise the evidence in this review. However, most of the included studies were not full economic evaluations, and therefore, some of the checklist questions were not applicable to the cost analysis study design. The lack of a standardised cost analysis quality appraisal checklist/tool limits the ability to quality appraise such studies (Xu et al., 2021). Due to the nature of such studies, they fail to meet some components of the JBI Critical Appraisal Checklist for Economic Evaluations. Notably, questions surrounding discounting, incremental analyses, and the comprehensive description of alternatives. The authors chose to extend the application of the JBI Critical Appraisal Checklist for Economic Evaluations checklist to cost analyses by awarding an equal point score to any element marked with an ‘NA’ to not penalise such studies. The scoring algorithm employed by the authors awarded a single point to any element marked Y or NA, while awarding no point to any element marked U or N. These points were totalled out of 11, and quality cut offs created to categorise the evidence into quality levels. For the costing studies, only the ‘cost’ aspect of questions 3, 5 and 6 was considered, and the outcome aspect was disregarded due to irrelevancy. Cut off scores are defined in this review as; 11 to 9 out of 11 – high quality, 6 to 8 out of 11 – moderate quality, 0 to 5 out of 11 – low quality.

Another limitation is that a specialist database of children’s palliative care studies was not searched. Future searches should include the Together for Short Lives website for children and young people’s palliative care (www.togetherforshortlives.org.uk).

### 4.4 Implications for policy and practice

This rapid review has shown that hospital-based palliative care costs are higher than hospice or home-based palliative care. However, hospice care can be less costly because of the number of volunteers helping to deliver the service and fundraise for the hospices, which are, in most cases, charities (at least in the UK) funded by the NHS and private donations.

Reducing hospital utilisation at the end of life should be a goal for healthcare planners only if access to quality home care at the end of life is guaranteed. Patients should be given a choice with regard to where they would prefer to die without moving the costs from the healthcare system to the home caregivers, rendering the costs invisible.

### 4.5 Implications for future research

More research is needed from the UK to determine the impact of new services such as Enhanced Supported Care (ESC) to examine which palliative care costs can be reduced with the implementation of such a programme.

Future research should consider which methods would be most appropriate to evaluate palliative care models. Standard economic evaluation methodology, such as the calculation of QALYs, may not be the most appropriate methodology in this end of life population (Wichmann et al., 2020). Prolonging death may be inconsistent with patient preferences and wishes. It is also important to consider appropriate patient reported outcome measures (PROMs) in this population. Some PROMs will be more suited to a particular condition e.g., dementia. PROMs are used in palliative care to evaluate the quality of care by quantifying various aspects of potential suffering, such as sleeplessness, loss of appetite, and pain. If PROM data is routinely collected, this data could be made available to researchers to evaluate end-of life care services.

## Data Availability

All data produced in the present study are available upon reasonable request to the authors

## 1.3 List of Abbreviations

Acronym: Full Description
ACP: Advance Care Planning - Advanced care planning is a voluntary process of person-centred discussion between an individual and their care providers about their preferences and priorities for their future care.
CAN$: The currency of Canada (dollars)
CBA: Cost-Benefit analysis
CEA: Cost-Effectiveness Analysis
CCT: Controlled Clinical Trial
CHF: Congestive Heart Failure
CKD: Chronic Kidney Disease
CNS: Clinical Nurse Specialist-led
COPD: Chronic Obstructive Pulmonary Disease
CPCC: Comprehensive Palliative Care Centre
CRC: Colorectal Cancer
CV: Cardiovascular
Decedents: Also understood as ‘deceased.’
End-of-life care (EoL): End-of-life care is for people who are thought to be in the last year of life. This time frame can be difficult to predict, so some people might only receive end of life care in their last weeks or days. Others may have end of life care for longer.
Enhanced Supported Care (ESC): Enhanced Supportive Care (ESC) is the prevention and management of the adverse effects of cancer and its treatment.
Euro €: The currency of the Eurozone
GBP £: The currency of the United Kingdom (Great British Pounds)
GP: General Practitioner
HBPC: Home Based Palliative Care
HSPC: Hospital Based Palliative Care
HatH: Hospice at home Service
HHC: Home healthcare Service
HIS: Hospital Inpatient Service
HC: Hospice Care
HRQoL: Health Related Quality of Life
ICER: Incremental Cost-Effectiveness Ratio
ICU: Intensive Care Unit
IPF: Idiopathic Pulmonary Fibrosis
JPY: Japanese Yen
KRW: Korean Won
MDC: Multidisciplinary collaborative
NHS: National Health Service
OECD: The Organisation for Economic Cooperation and Development (OECD)
Hospice: A home providing care for the sick or terminally ill.
NOK: The currency of Norway
NSC: Non-Specialist Care
ONS: Office for National Statistics
Palliative care (PC): Palliative care is an interdisciplinary medical caregiving approach aimed at optimising quality of life and mitigating suffering among people with serious, complex, and often terminal illnesses
PCDS: Palliative Care Day Services
PCS: Palliative Care Services
PCCS: Palliative Care Consultation Services
PCU: Palliative Care Units
PFC: Partners for Children (PFC), a paediatric palliative care pilot programme offering hospice-like services for children eligible for full- scope Medicaid delivered concurrently with curative care, regardless of the child’s life-expectancy.
QALD: Quality-Adjusted Life Day
QALW: Quality-Adjusted Life Week
QALY: Quality-Adjusted Life Year
QoL: Quality of Life
RR: Rapid Review
RCT: Randomised Controlled Trial
SC: Specialist Care
SNF: Skilled Nursing Facility
SR: Systematic Review
SROI: Social Return on Investment
UK: United Kingdom
US$: The currency of the USA (dollars)
USA: United States of America
WEC: Wales Evidence Centre (Health and Care Research Wales Evidence Centre)
WHO: World Health Organization
ZBI: Zarit Carer Burden Inventory

## 6. RAPID REVIEW METHODS

### 6.1 Eligibility criteria

The eligibility criteria are described in Table 8.

**Table 8:** Eligibility Criteria (PICo: Population, phenomenon of Interest, Context)

|  | **Inclusion criteria** | **Exclusion criteria** |
| --- | --- | --- |
| **Population** | Children and adults receiving palliative or end of life care | People not receiving palliative or end of life care |
| **Phenomenon of interest** | Models of palliative and end of life care | Models of care not related to palliative or end of life care |
| **Context** | Service delivery of palliative care or end of life care in healthcare systems like the UK. | Services not relevant to palliative or end of life care in countries dissimilar to the UK. |
| **Outcome measures** | Primary outcomes(s): cost-effectiveness, cost of palliative care or end of life care. | Costs unrelated to palliative care or end of life care. |
| **Study design** | Economic evidence of cost and cost-effectiveness of models of service delivery for palliative or end of life care. | Studies not including economic evidence of palliative care or end of life care. |
| **Countries** | We will include studies from the UK and countries where palliative care and end of life services are similarly comparable to Wales, including OECD countries. | Countries other than OECD countries where palliative and end of life care delivery is vastly different to the UK. |
| **Language of publication** | English or Welsh | Languages other than English or Welsh |
| **Publication date** | January 2003 to present |  |
| **Publication type** | Published and preprint | Protocols, editorials, letters, commentaries. |
| **Other factors** | None. |  |

### 6.2 Literature search

The search strategy conducted in Medline through OVID is presented in Appendix 1. Dates of the searches were from January 2003 to October 2023. We limited the dates of the searches to only include recent evidence from the past twenty years due to the anticipated substantial number of database hits. Previous high calibre Cochrane Reviews have previously identified literature from earlier dates. The following databases were used for the searches:

Medline

EMBASE

Cochrane Library

CINAHL

### 6.3 Study selection process

Two reviewers screened 100% of titles and abstracts independently using the Covidence review management software. After this, the level of agreement was assessed with disagreements settled by discussion and consensus. During independent screening, the review lead (LHS) consulted with the two reviewers to come to an agreement on the final inclusions if there was ongoing disagreement.

### 6.4 Data extraction

Data extraction was based on the outlined eligibility criteria. We extracted details/characteristics on study country, study design, type of intervention/model, type of economic evaluation, perspective of analysis, number of participants, relevant costs, and outcomes (see eligibility criteria) and study settings. All four members of the core BIHMR team were involved with the data extraction with the review lead (LHS) checking 25% of the data extraction tables and the other reviewers checking the remaining 25%.

### 6.5 Quality appraisal

Full economic evaluations and cost studies were assessed with the JBI economic evaluations checklist (Joanna Briggs Institute, 2022).

### 6.6 Synthesis

Due to the heterogeneity of the costs and outcomes in the included studies, a narrative synthesis of the results was reported.

## 7. EVIDENCE

### 7.1 Search results and study selection

The title and abstract searches yielded 101 inclusions. Full texts (n=77) were reviewed, and n=8 Systematic Reviews (SRs) and n=48 primary studies were included in this Rapid Review (RR). The PRISMA flow diagram is shown in Figure 1.

### 7.2 Data extraction

The data extraction are shown below in Tables 9-–20.

**Table 9:** Evidence for costs of hospital based palliative care: Cancer.

| Citation (Country)<br>Aim | Model of care | Study characteristics, health economics methods and quality appraisal | Outcome and costs measured | Main health economics findings |
| --- | --- | --- | --- | --- |
| Tan and Jatoi., 2011<br>(Tan & Jatoi, 2011)<br>(USA)<br>**Aim:** To explore whether advance directives or route of hospital admission reduced cancer patients' terminal hospitalisation costs. | **Intervention (specialist or generalist palliative care):** Specialist cancer care<br>**Intervention recipients and sample size:** n = 120 solid tumour oncology patients.<br>**Setting:** Hospital setting in the USA.<br>**Delivery mode (e.g., remotely online, in person):** In person.<br>**Intervention deliverers (e.g., professionals or paid carers):** Healthcare professionals within hospital.<br>**Timing and duration:** Median time spent at hospital before death was 4 days.<br>**Intervention description:** Last hospitalisation before death for solid tumour oncology patients. | **Dates of data collection:** Between 2008 and 2009<br>**Length of follow-up:** No follow-up.<br>**Type of economic evaluation/cost analysis:** Cost analysis.<br>**Perspective of analysis:** Healthcare system perspective<br>**Currency and cost year:** USD\$, cost year not stated.<br>**Discounting:** Not conducted.<br>**Sensitivity analysis:** No. | **Outcome(s): Costs only.**<br>(Patients' total costs were compared based on advance directives and route of hospitalisation).<br>**Types of costs measured**<br>Total cost (defined as the entire sum of money required to provide a medical service after adjustment for the average cost of such services for a given year) | **Base case results:**<br>The median total cost for hospitalisation per patient was \$12,962 (range: 1,244–138,877). The IQR of 25–75% was \$6,760–\$23,375. Patients who had an advance directive had a median total terminal hospitalisation cost per patient of \$12,840, and those without cost \$13,084 ($p < 0.30$). Patients who had been admitted to the hospital after an oncology clinic visit or hospice visit had a median total terminal hospitalisation cost per patient of USD\$25,320, and those who had entered the hospital via another pathway had a cost of USD£24,335 ($p < 0.43$). After adjusting for patient age at death and days in the hospital, the existence of an advance directive was not associated with a statistically significant difference in total hospital cost ($p < 0.24$). The same was found for route of hospitalisation ($p < 0.51$). |
| Schneider et al., 2020 | **Intervention (specialist or generalist palliative care):** Specialist care (breast cancer). | **Dates of data collection**<br>Between January 2010 and June 1<sup>st</sup> 2017. | **Outcome(s)**<br>Costs only. (Costs of hospital care over the last | **Base case results:** Mean end of life costs per patient were €21,641 (SD = €20,147). Over |
| (Schneider et al., 2020)<br>(The Netherlands)<br>**Aim:** To explore the variability of longitudinal patterns of costs in advanced breast cancer patients in the Netherlands during end of life care (last 12 months of life). | **Intervention recipients and sample size:** n = 558 patients who died of advanced breast cancer.<br>**Setting:** Hospital setting in The Netherlands.<br>**Delivery mode (e.g., remotely online, in person):** In person.<br>**Intervention deliverers (e.g., professionals or paid carers):** Healthcare professionals.<br>**Timing and duration:** Last 12 months of life assessed.<br>**Intervention description:** Hospital admission and care for advanced breast cancer | **Length of follow-up:** No follow-up.<br>**Type of economic evaluation/cost analysis:** Prospective costing study.<br>**Perspective of analysis:** Hospital perspective in the Netherlands.<br>**Currency and cost year:** Euro, cost year not defined.<br>**Discounting:** Not conducted.<br>**Sensitivity analysis:** Yes. | twelve months before death were analysed, and the variability of longitudinal patterns between patients were explored using group-based trajectory modelling).<br>**Types of costs measured**<br>• Medication and transfusions<br>• Local treatments and procedures<br>• Consultations and hospitalisations<br>• Diagnostic procedures | the last twelve months before death, the average costs per patient month were €2,255 with a standard deviation of €492. From month 12 until month 5 before death, overall monthly costs remained stable with a mean of €1,984. From month 5, mean costs per month steadily increased with an average increase of €343 per month, reaching a maximum of €3,614 during the last month before death.<br>Medication costs reduced after month 3, while hospitalisation costs increased from month 5 before death. Diagnostic and local procedures were marginal.<br>**Sensitivity analysis:** A sensitivity analysis in the subgroup of patients with at least twelve months survival time was conducted. Six latent cost trajectory groups were extracted, which were very similar to the groups found in the full cohort model. Minor differences in latent trajectory patterns were observed. |

**Table 10:** Evidence for costs of hospital based palliative care: Chronic Kidney Disease (CKD)

| Citation (Country)<br>Aim | Model of care | Study characteristics, health economics methods and quality appraisal | Outcome and costs measured | Main health economics findings |
| --- | --- | --- | --- | --- |
| Kerr et al., 2017<br>(Kerr et al., 2017)<br>(UK)<br>**Aim:** To estimate causes and place of death as well as cost of associated hospital care for people with Chronic Kidney Disease (CKD) in England in the final 3 years of their life. | **Intervention (specialist or generalist palliative care):** Generalist hospital care<br>**Intervention recipients and sample size** 211,215 individual records used from data collection period.<br>**Setting:** Hospital setting in the UK.<br>**Delivery mode (e.g., remotely online, in person):** In person delivery.<br>**Intervention deliverers (e.g., professionals or paid carers):** Hospital medical staff.<br>**Timing and duration:** Between April 1 2006 and 31 March 2010.<br>**Intervention description:** Hospital admission for end stage chronic kidney disease | **Dates of data collection:** Between April 1 2006 and 31 March 2010<br>**Length of follow-up:** No follow-up.<br>**Type of economic evaluation/cost analysis:** Cost analysis using historic Hospital Episode Statistics and Office for National Statistics Morbidity data.<br>**Perspective of analysis:** Not defined.<br>**Currency and cost year:** GBP, cost year not stated.<br>**Discounting:** Not applied.<br>**Sensitivity analysis:** Yes. | **Outcome(s):**<br>Costs of death and place of death.<br>**Types of costs measured:** Where costs were obtained not defined. Total hospital costs presented. | **Base case results:** The mean cost of hospital admissions and outpatient care in the 12 months before death was £11,916 for people with CKD and £7,832 for standardized non-CKD. For both groups, >50% of final year costs arose in the final 3 months, and around a quarter in the final month. For both CKD and non-CKD patients, hospital costs were lower for those who died at home over 12-, 3- and 1-month perspectives. However, the greatest impact of home death on costs was observed in the final 30 days of life.<br>For people with CKD, the mean cost of hospital care in the 12 months before death was £9,877 for those who died at home and £12,160 for those who died elsewhere. The mean cost in the final 30 days of life was £1,077 for those who died at home and £3,206 for those who died elsewhere.<br>**Sensitivity analysis:**<br>Conducted by removing 5.6% of patients with highest hospital costs over final year of life to reduce bias from this group. |
| Pollock et al., 2022 (Pollock et al., 2022)<br>(USA)<br>**Aim:** To examine the length of hospitalisation and costs associated with end of life inpatient encounters using retrospective data on kidney failure, cardiovascular (CV) and infection-related admissions | **Intervention (specialist or generalist palliative care):** Specialist care (Kidney failure; Cardiovascular (CV) and infection-related admissions)<br>**Intervention recipients and sample size** n=25,118 (CV), n= 4,210 (kidney failure), n=76,3017 (infection).<br>**Setting:** Hospital setting in the USA.<br>**Delivery mode (e.g., remotely online, in person):** In person.<br>**Intervention deliverers (e.g., professionals or paid carers):** Medical professionals (physicians and nurses)<br>**Timing and duration:** Mean length of stay.<br>**Intervention description:** Hospital based acute care for the listed conditions. | **Dates of data collection:** Hospital records from January 2016 to March 2020.<br>**Length of follow-up:** No follow-up.<br>**Type of economic evaluation/cost analysis:** Retrospective costing analysis.<br>**Perspective of analysis:** Hospital perspective.<br>**Currency and cost year:** USD\$, 2019 cost year.<br>**Discounting:** Not conducted.<br>**Sensitivity analysis:** No. | **Outcome(s):** Costs of palliative care and costs of death.<br>**Types of costs measured:** Medical care costs. Medication costs. Procedural costs. | **Base case results:** Kidney failure-related encounters incurred longer hospital stays and higher costs than either CV or infection-related encounters.<br>The median [interquartile range (IQR)] total cost of any inpatient encounter was \$17,057 (\$8,040–\$35,873). The median (IQR) costs, stratified by the reason for the encounter, were \$18,469 (\$8637–\$38,315) for kidney failure, \$17,503 (\$7,766–\$39,693) for CV and \$16,403 (\$,7762–\$34,910) for infection. |

**Table 11:**
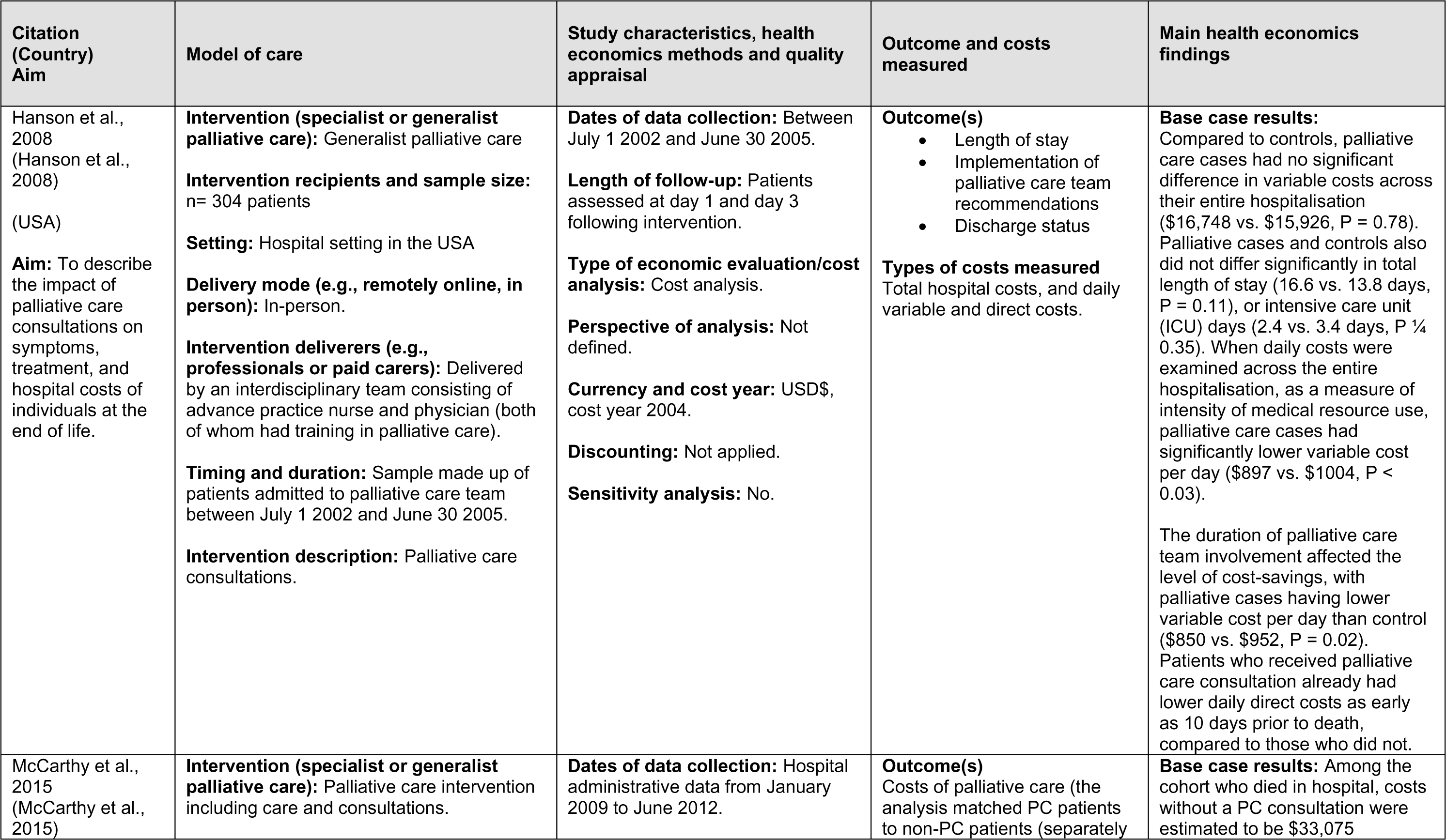
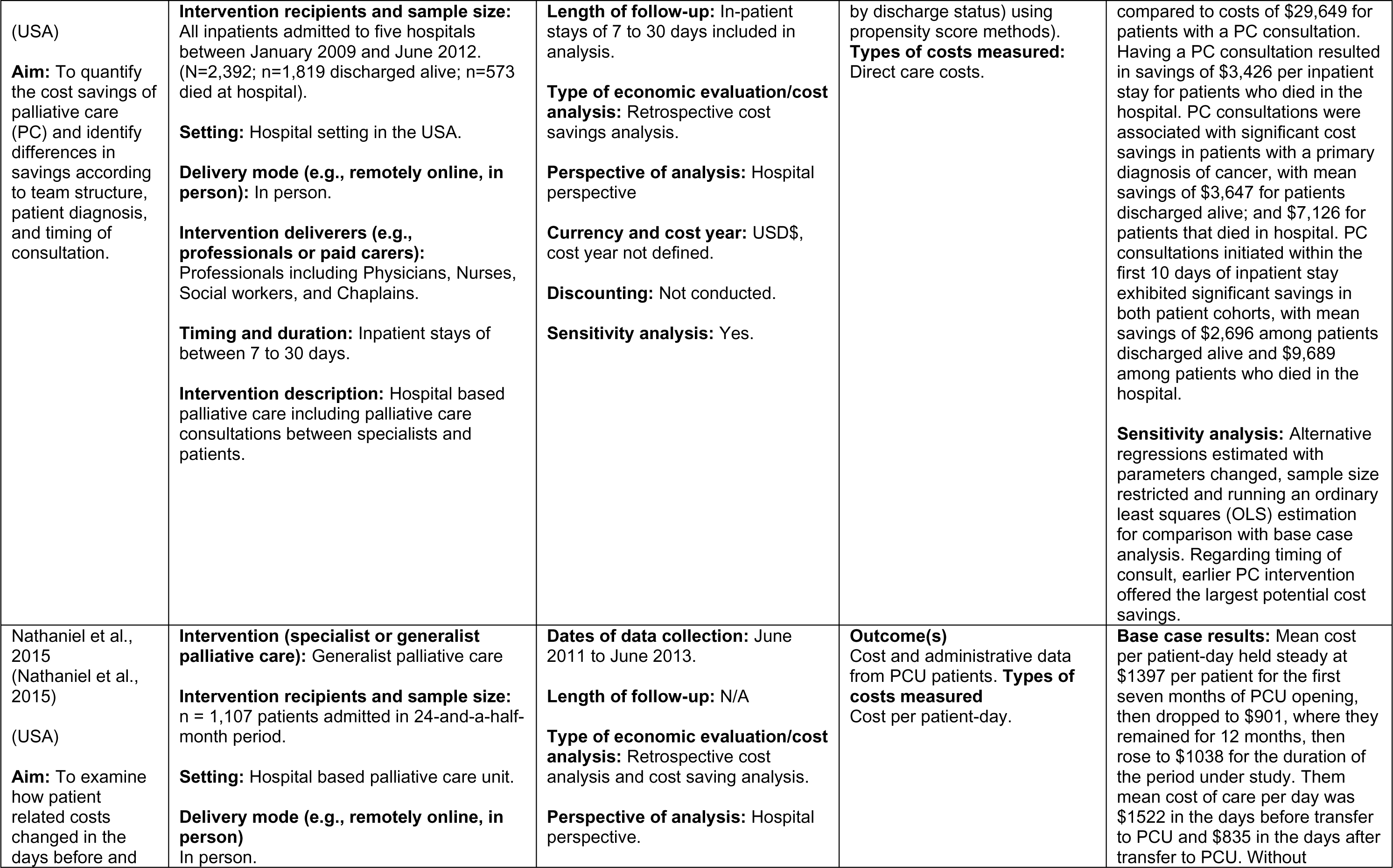
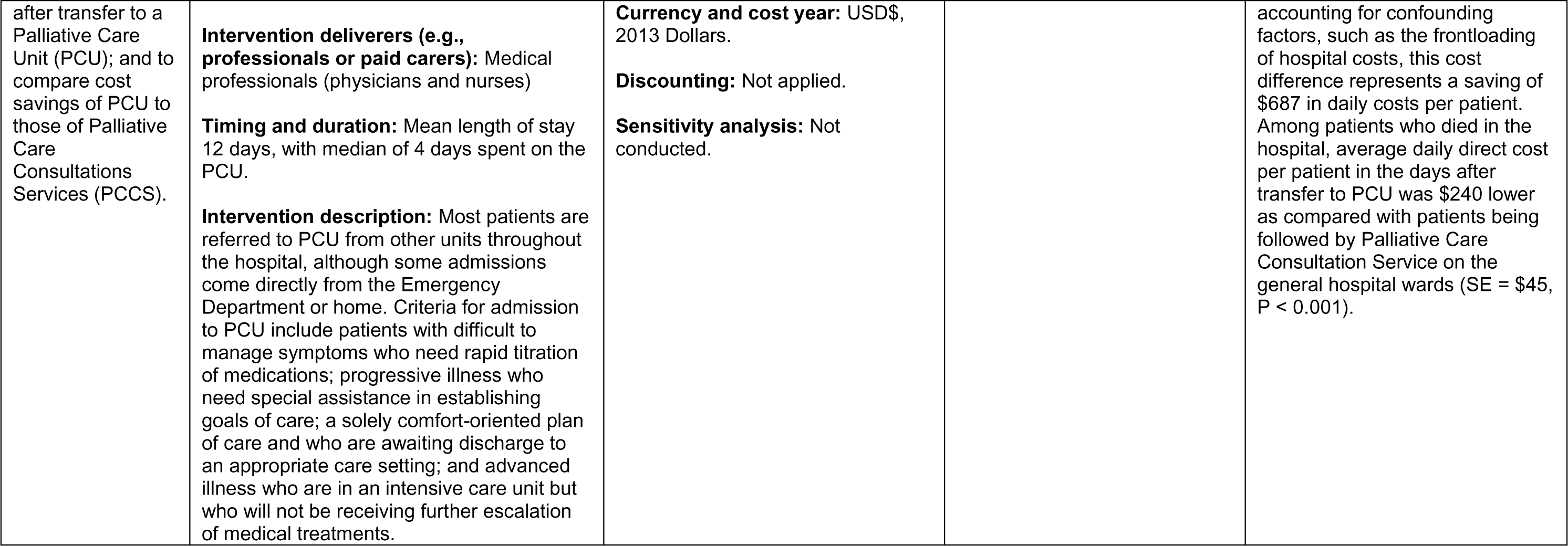
Evidence for costs of hospital based palliative care: General.

**Table 12:** Evidence for costs of hospital based palliative care: Economic evaluations.

| Citation<br>(Country)<br>Aim | Model of care | Study characteristics, health economics methods and quality appraisal | Outcome and costs measured | Main health economics findings |
| --- | --- | --- | --- | --- |
| Isenberg et al., 2017<br>(Isenberg et al., 2017)<br>(USA)<br>**Aim:** To establish costs of an inpatient palliative care unit (PCU) and conduct a threshold analysis to estimate the maximum possible costs for the Palliative Care Units (PCU) to be considered cost-effective. | **Intervention (specialist or generalist palliative care):** Specialist care<br>**Intervention recipients and sample size:** n =153 patient encounters during data collection period.<br>**Setting:** Hospital setting in the USA.<br>**Delivery mode (e.g., remotely online, in person):** In person – inpatient admission.<br>**Intervention deliverers (e.g., professionals or paid carers):** Physician and or advance practice nurse.<br>**Timing and duration:** March 2013 to March 2014.<br>**Intervention description:** Combination of specialist care support (Chaplaincy and social work) and active management (transfusions, pain therapy, physical therapy, and radiation therapy). | **Dates of data collection:** March 2013 to March 2014.<br>**Length of follow-up:** No follow-up.<br>**Type of economic evaluation/cost analysis:** Cost-effectiveness threshold analysis.<br>**Perspective of analysis:** Health services cost perspective.<br>**Currency and cost year:** USD\$, cost year not defined.<br>**Discounting:** Not conducted as analysis only one year.<br>**Sensitivity analysis:** No. | **Outcome(s):** Quality-adjusted life years from the literature.<br>**Types of costs measured:** Palliative direct costs (patients transferred from emergency department or clinics).<br>Palliative transfers (patients admitted from other departments)<br>Professional fees for physician services. | **Base case results:** Based on the contribution margin, the PCU produces cost savings and is profitable for the hospital. Based on the variable costs, the programme needed to cost a maximum of \$559,800 (\$716 per encounter per day) to be rendered cost-effective; however, the programme generated \$1,050,031 in costs (\$1,343 per encounter per day). When factoring in the cost-minimization analysis, the programme is cost-saving from both hospital standpoints: variable costs and contribution margin. |
| Sellars et al., 2019<br>(Sellars et al., 2019)<br>(Australia)<br>**Aim:** To examine hospital costs and outcomes of a nurse-led advanced care | **Intervention (specialist or generalist palliative care):** Specialist care (advanced care planning in kidney disease)<br>**Intervention recipients and sample size:** Hypothetical cohort of patients<br>**Setting:** Hospital setting in Australia.<br>**Delivery mode (e.g., remotely online, in person)** | **Dates of data collection:** Unclear.<br>**Length of follow-up:** No-follow up.<br>**Type of economic evaluation/cost analysis:** Prospective cost-effectiveness analysis<br>**Perspective of analysis:** Hospital perspective | **Outcome(s):** Costs of end-of life treatment and preferences being met or not.<br>**Types of costs measured:**<br>Hospital costs including direct care costs, consultation costs and recruitment/scheduling | **Base case results:** The cost of implementing the ACP intervention was on average \$519 per patient. The average cost per patient for the ACP group was \$100,579 (SD = 17,356) and the proportion of patients receiving end of life care according to preferences was 68% (SD = 48). In the no |
| planning (ACP) intervention compared with usual care in the last 12 months of life for older people with end-stage kidney disease managed with haemodialysis. | In person.<br>**Intervention deliverers (e.g., professionals or paid carers):** Health professionals (nurse-led).<br>**Timing and duration:** Last 12 months of life modelled.<br>**Intervention description:** Modelled nurse-led advanced care planning. | **Currency and cost year:** AUD\$, cost year not defined.<br>**Discounting:** Not conducted<br>**Sensitivity analysis:** Yes | Programme set up costs | ACP group, the average cost per patient was \$87,282 (SD = 19,078) and the proportion of patients having preferences met was 24% (SD = 43). The average hospital costs incurred by patients in the last 12 months of life was higher for patients who withdrew from dialysis versus those who died from other causes (\$110,696 vs. \$71,737,<br>The last 12 months of life for those undergoing ACP was more expensive yet more effective in facilitating adherence to patient preferences than usual care. The incremental cost per additional case of end of life preferences being met (incremental cost-effectiveness ratio [ICER]) was **\$28,421**. Australian cost-effectiveness threshold is between \$30,000AUD to \$70,000AUD per QALY gained.<br>**Sensitivity analysis:**<br>A series of one-way sensitivity analyses were performed to evaluate the robustness of the model. |

**Table 13:**
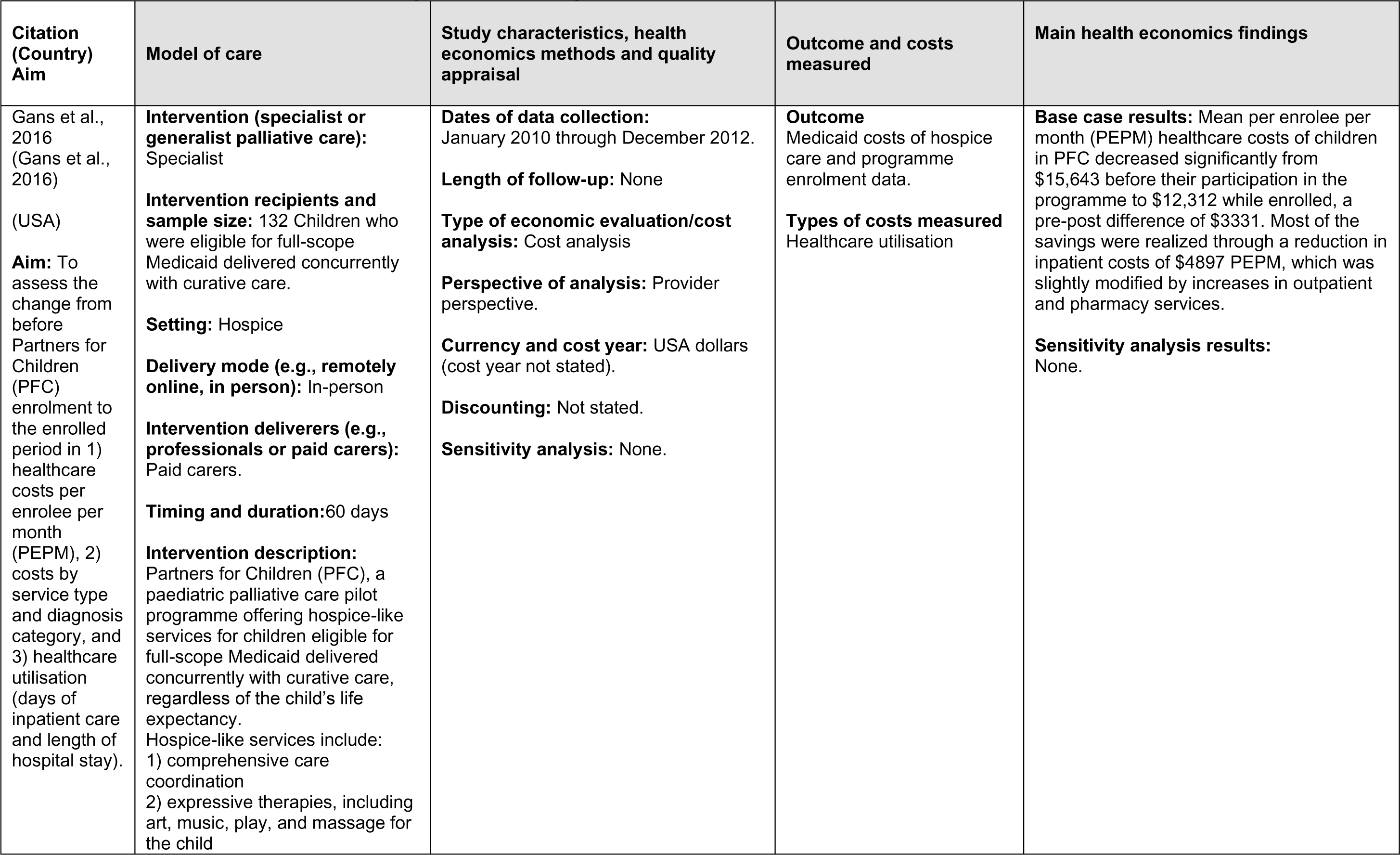
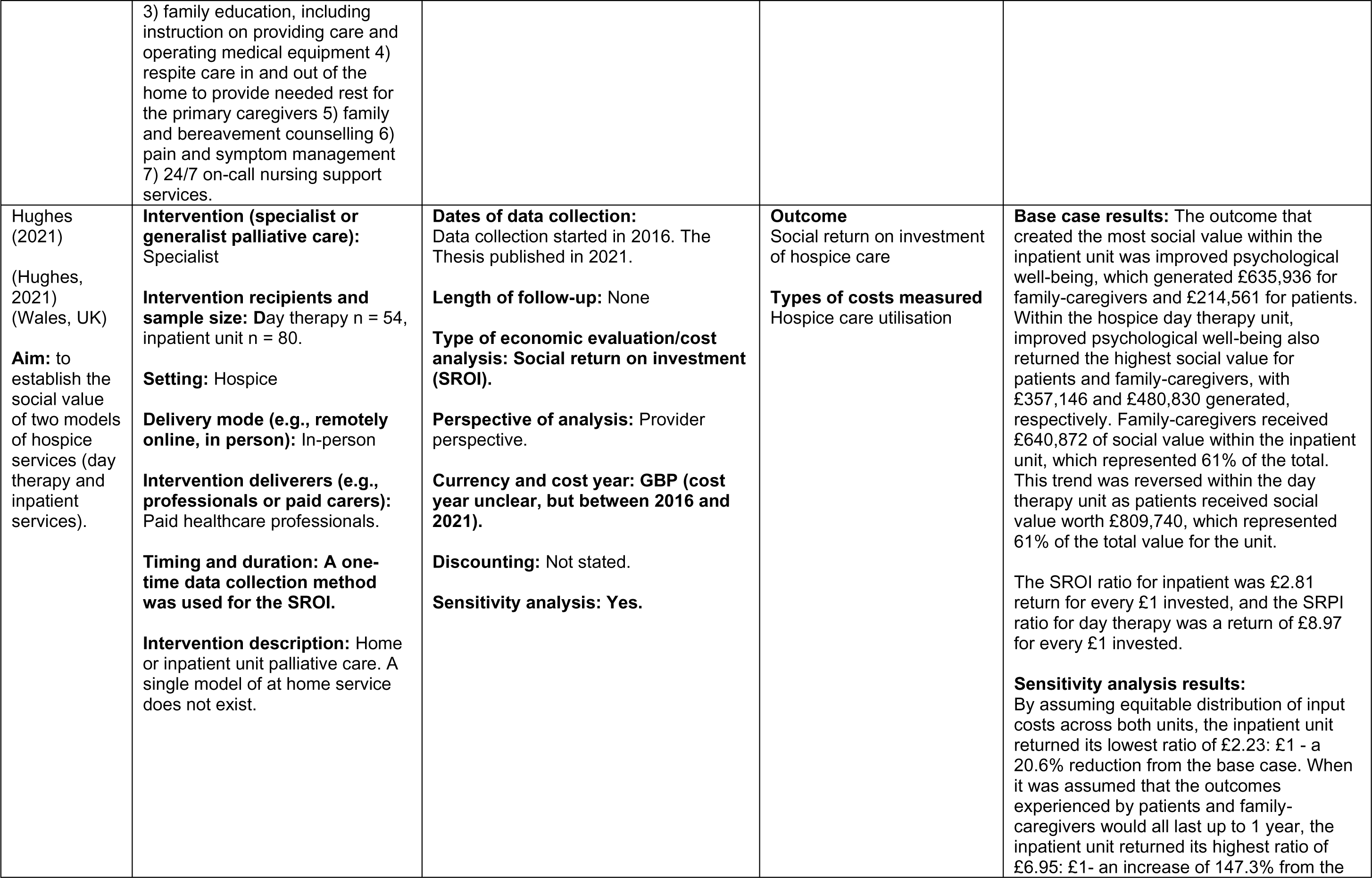
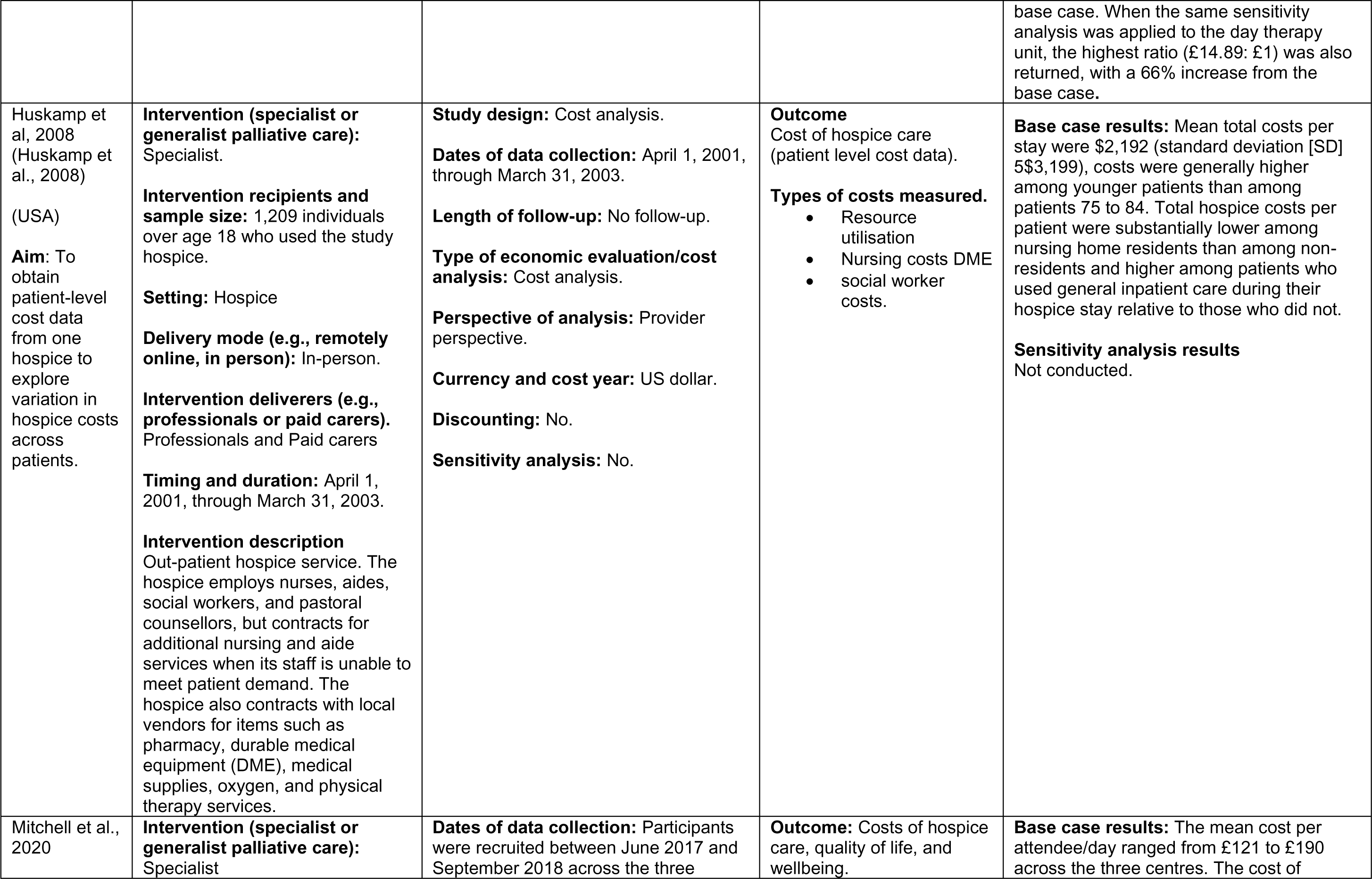
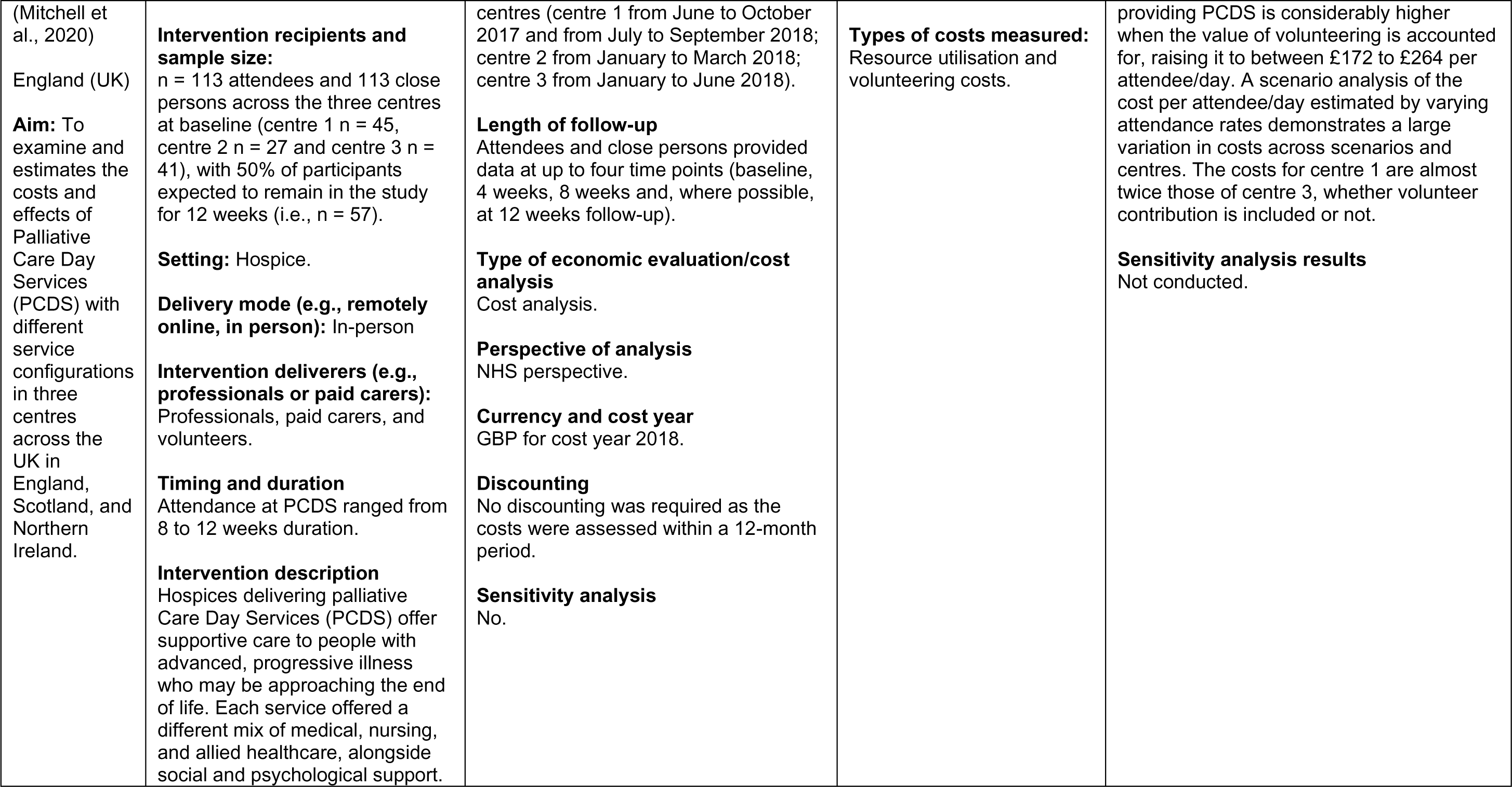
Evidence for costs of hospice models of palliative care.

**Table 14:** Evidence for costs of community-based models of palliative care.

| Citation (Country)<br>Aim | Model of care | Study characteristics, health economics methods and quality appraisal | Outcome and costs measured | Main health economics findings |
| --- | --- | --- | --- | --- |
| Amador et al. 2014 (Amador et al., 2014)<br>(England, UK)<br>**Aim:** To describe end of life care costs of older people with dementia (OPWD) residents in care homes (CHs) with no on-site nursing and evaluate the economic case for an intervention designed to improve end of life care for OPWD in CHs. | **Intervention/care (specialist or generalist palliative care):** Generalist.<br>**Intervention/care recipients and sample size:** Older people with dementia<br>Sample size during phase 1: 133 OPWD in six residential CHs. Sample size during phase 2: Seventy-four residents were subsequently recruited across the three intervention homes in Phase 2, 28 of whom had participated in Phase 1.<br>**Setting:** Community – residential care homes<br>**Intervention/care deliverers (e.g., professionals or paid carers):** Care home (CH) staff and visiting healthcare professionals.<br>**Timing and duration:** EOL stage of participants not explicitly described in paper. Phase 1: tracked for a year, from March 2009, health services received by 133 OPWD in six residential CHs in the East of England. Phase 2: Intervention delivered over 6 months from January 2011.<br>**Intervention/model of care description:**<br>Phase 1: Costs of services received by OPWD over a year time horizon.<br>Phase 2: The 'Appreciative Inquiry' (AI) intervention designed to foster collaborative working among CH staff and visiting healthcare professionals to facilitate co-development of context-specific innovations<br>to improve care for OPWD at end of life (EOL). | **Dates of data collection:** Phase 1: March 2009 – Feb 2010<br>Phase 2: January 2011<br>**Length of follow-up:** NA. Cost for Phase 2 collected over 6-month period.<br>**Type of economic evaluation/cost analysis:** Cost analysis<br>**Perspective of analysis:** Not defined.<br>**Currency and cost year:** British pounds sterling, for cost year 2010.<br>**Discounting:** No discounting as study time horizon less than 12 months.<br>**Sensitivity analysis:** No. | **Outcome:** Costs.<br>**Types of costs measured:** Resource utilisation - hospital, community, and primary care health costs, medication, and accommodation costs. | **Base case results:** Phase 1: Costs for each resident in Phase 1 were about £2800 per month, including service, accommodation, and medication. Resource use was associated with resident characteristics.<br>Phase 2: The intervention was perceived as having a positive impact on working relationships between CHs and visiting healthcare practitioners. Following the intervention total service costs fell by 43%. Hospital care costs fell by 88%.<br>**Sensitivity analysis results:** None. |
| Bentur et al. 2014 (Bentur et al., 2014)<br>(Israel)<br>**Aim:** To examine the utilisation and cost of all health services consumed during the last six months of life by cancer patients, and compared those with and without home-hospice care. | **Intervention/care (specialist or generalist palliative care):** Specialist<br>**Intervention/care recipients and sample size:** Cancer patients, 429 individuals who had lived in the community and died of metastatic cancer between January and September 2009.<br>**Setting:** Community – home hospice care<br>**Intervention/care deliverers (e.g., professionals or paid carers)** Multidisciplinary palliative care teams including physicians, nurses, and social workers.<br>**Timing and duration:** Data collection of services consumed during last 6-months of life.<br>**Intervention/care description:** The health plan supplies community care that includes family physicians, nurses, home care, hospice care, and oncology day care. There are 4 small hospitals in the district. Referrals to home hospice unit (HHU) care are initiated by all those services. The HHU is a 24-hour service provided by a multidisciplinary palliative care team that includes physicians, nurses, and social workers who visit the patient's home once a week or more, as needed. | **Dates of data collection:** 6 months prior to death for individuals who died between January and September 2009.<br>**Length of follow-up:** NA - Costs collected over 6-month time horizon.<br>**Type of economic evaluation/cost analysis:** Cost analysis<br>**Perspective of analysis:** Not defined.<br>**Currency and cost year:** US Dollars (US\$) for cost year 2010<br>**Discounting:** No discounting as study time horizon less than 12 months.<br>**Sensitivity analysis:** Not conducted. | **Outcome:** Costs<br>**Types of costs measured.**<br>• Healthcare resource use<br>• Hospital admission<br>• Emergency room (ER) visits<br>• Medication<br>• Enrolment in home care unit<br>• Enrolment in home hospice unit<br>• Oncology day care. | **Base case results:** The average cost of care for the last 6 months of life, for patients with HHU care, was US\$13 648 compared to US\$18 503 for patients without HHU care. Hospitalisation contributed 32% to the cost of patients with HHU care, and 64% of the total expenditure for patients without HHU care. The findings support the justification for significant expansion of home-hospice care.<br>**Sensitivity analysis results** None. |
| Butler et al. 2022 (Butler et al., 2022) (England)<br>**Aim:** To find out what models of hospice at home services work best, | **Intervention/care (specialist or generalist palliative care):** Generalist<br>**Intervention/care recipients and sample size:** A total of 339 patient and family/informal carer dyads were recruited from 12 HAH services. Of these, 221 (65.2%) provided service use data by completing one or more | **Dates of data collection:** Prior to March 2021.<br>**Length of follow-up:** NA<br>**Type of economic evaluation/cost analysis:** Cost analysis | **Outcome:** NA<br>**Types of costs measured:** Resources and costs of running each case study HAH service, covering staff; service facilities, inpatient beds, | **Base case results:** Costs per day were higher closer to death: the median daily costs for 0–14, 15–28 and 29–92 days before death for all formal care were £104.57, £80.08, and £56.07, respectively. Among these |
| for whom and in what circumstances. Other objectives supported this aim, including an analysis of the health economic costs of hospice at home models. | Ambulatory and Home Records. Of these 221, it was possible to include 178 (80.5%) in the analysis of costs. Information on the resources involved in running each hospice and costs was sought through interviews with case study site managers during phase 1.<br>**Setting:** Community – hospice at home services<br>**Intervention/care deliverers (e.g., professionals or paid carers):** Hospice at home services led by registered nurses and healthcare assistants.<br>**Timing and duration:** Costs calculated retrospectively for different time points before death.<br>**Intervention/model of care description:** No intervention but a realist evaluation of hospice at home services in England. | **Perspective of analysis:** Not defined.<br>**Currency and cost year:** British pounds sterling for cost year 2019.<br>**Discounting:** Not required<br>**Sensitivity analysis:** Not conducted. | equipment, overheads; transport for home care; and other sundry items associated with care delivery. Formal and informal care costs were also considered. | overall costs, the median daily costs for in-home nursing and personal/social care were £40.43, £27.93, and £12.22 for 0–14, 15–28 and 29–92 days before death, respectively. Informal care costs, valued at replacement costs (support worker), exceed formal care costs, with daily medians of £580.00, £449.50, and £348.00 for 0–14, 15–28 and 29–92 days before death, respectively.<br>**Sensitivity analysis results**<br>None |
| Chai et al. 2014 (Chai et al., 2014)<br>(Canada)<br>**Aim:** To (i) assess the magnitude and share of unpaid care costs in total healthcare costs for home-based palliative care patients, from a societal perspective and (ii) examine the sociodemographic and clinical factors that account for variations in this share. | **Intervention/care (specialist or generalist palliative care):** Specialist<br>**Intervention/care recipients and sample size:** 169 caregivers of patients with a malignant neoplasm were interviewed, results for 129 participants reported in the analysis.<br>**Setting:** Community – home-based palliative care<br>**Intervention/care deliverers (e.g., professionals or paid carers):** Home-based palliative care delivered by a multidisciplinary team and provided by unpaid family and friend caregivers.<br>**Timing and duration:** Data collection of services consumed during last 12-months of life. | **Dates of data collection:** July 2005 and September 2007<br>**Length of follow-up:** NA - Costs collected over 12-month time horizon.<br>**Type of economic evaluation/cost analysis:** Cost analysis.<br>**Perspective of analysis:** Societal perspective.<br>**Currency and cost year:** Canadian dollars valued in 2008 and inflated to 2011 prices.<br>**Discounting:** No discounting as time horizon was 12 months. | **Outcome: Costs over the last 12 months of life.**<br>**Types of costs measured:** Healthcare resource use and unpaid care. Costs were grouped into three categories: Publicly financed costs, privately financed costs, and Unpaid care-giving costs. | **Base case results:** Over the last 12 months of life, the average monthly cost was \$14 924 (2011 CDN\$) per patient. Unpaid care-giving costs were the largest component – \$11 334, accounting for 77% of total palliative care expenses, followed by public costs (\$3211; 21%) and out-of-pocket expenditures (\$379; 2%). In all cost categories, monthly costs increased exponentially with proximity to death. Unrelated regression estimation suggested that the share of unpaid care costs of total costs was driven by patients' and caregivers' sociodemographic |
|  | **Intervention/model of care description**<br>The Temmy Latner Centre for Palliative Care (TLCPC) provides multidisciplinary, home-based care (24 hours per day, 7 days per week) for terminally ill patients in partnership with the Toronto Community Care Access Centre. | **Sensitivity analysis:** Not conducted. |  | characteristics. Results suggest that overwhelming the proportion of palliative care costs is unpaid caregiving.<br>**Sensitivity analysis results**<br>None |
| Chen et al. 2018<br>(Chen et al., 2018)<br>(USA)<br>**Aim:** To assess the Medicare reimbursement savings of an established palliative care homebound programme. | **Intervention/care (specialist or generalist palliative care):** Specialist.<br>**Intervention/care recipients and sample size:** Older Adults with Advanced Medical Illness.<br>50 participants enrolled in a palliative care homebound programme and 95 propensity-matched control patients receiving usual care.<br>**Setting:** Community – homebound programme<br>**Intervention/care deliverers (e.g., professionals or paid carers):** Delivered by an interdisciplinary team (IDT) led by five certified palliative care physicians and two certified palliative care APRNs, in addition to several certified geriatricians and geriatric APRNs, social workers and community health workers, RN care coordinators, and pharmacists.<br>**Timing and duration:** Programme described as selective life extending therapy available for patients who either desire them or are not predictably dying in the next six months. Eligible patients had elder risk assessment (ERA) scores of greater than 15, and either four-year mortality score greater than 13 or a three-year mortality score greater than 5 as noted below.<br>Participants enrolled between September 1, 2012, and March 31, 2013. | **Dates of data collection:** Enrolment was between September 1, 2012, and March 31, 2013. Total Medicare reimbursement was compared in the year before enrolment with the year after enrolment for participants and control.<br>**Length of follow-up:** NA. Estimated Medicare reimbursement was calculated for the 12 months before programme enrolment through the 12 months after enrolment. Similar time frames were used for control patients.<br>**Type of economic evaluation/cost analysis:** Cost analysis<br>**Perspective of analysis:** Not defined.<br>**Currency and cost year:** US dollars for cost year 2014<br>**Discounting**<br>No discounting as time horizon was 12 months.<br>**Sensitivity analysis**<br>Not conducted | **Outcome:** Medicare costs.<br>**Types of costs measured:** Estimated Medicare reimbursement was calculated using 2014 fee schedules based on services billed by Mayo Clinic. | **Base case results:** The Mayo Clinic Palliative Care Homebound Programme reduced annual Medicare expenditures by \$18,251 per programme participant compared with matched control patients. Total Medicare reimbursement per programme participant the year before programme enrolment was \$16,429 compared with \$14,427 per control patient, resulting in \$2004 higher charges per programme patient. In 12 months after programme enrolment, mean annual payment was \$5783 per patient among participants and \$22,031 per patient among the matched controls. In the second year, the intervention group had a decrease of \$10,646 per patient; the control group had an increase of \$7604 per patient. The difference between the participant group and control group was statistically significant ($P < 0.001$) and favoured the palliative care homebound programme |
| | **Intervention/model of care description**<br>It is a specialised palliative care service delivered to eligible primary care patients with advanced medical illness and limited life expectancy. | | | enrollees by \$18,251 (95% CI, \$11,268e\$25,234).<br>**Sensitivity analysis results**<br>None |
| Enguidanos et al. 2005<br>(Enguidanos et al., 2005)<br>(USA)<br>**Aim:** To examine differences in site of death and costs of services by primary diagnosis for patients receiving home-based palliative care as compared to usual care at the end of life. | **Intervention/care (specialist or generalist palliative care):** Specialist<br>**Intervention/care recipients and sample size**<br>Patients with Congestive Heart Failure, Chronic Obstructive Pulmonary Disease, and Cancer. 210 participants enrolled in the Kaiser Permanente Palliative Care Programme (KPCC) and 348 were enrolled in the comparison group (usual care).<br>**Setting**<br>Community – home-based palliative care<br>**Intervention/care deliverers (e.g., professionals or paid carers)**<br>Physicians, nurses, social workers, and other healthcare professionals.<br>**Timing and duration**<br>Participants had an estimated prognosis of approximately one year of life expectancy.<br>**Intervention/model of care description**<br>The KPCC programme is an interdisciplinary home-based model of care designed to provide treatment with the primary goals of enhancing comfort and improving quality of care in a patient's last year of life. The KPCC programme used an interdisciplinary team approach, with the central care team composed of the patient and family, and physician, nurse, and social worker all with expertise in symptom management and biopsychosocial intervention. | **Dates of data collection:** Patients were enrolled from March 1999 through August 2000 and died during the two-year study.<br>**Length of follow-up:** NA<br>**Type of economic evaluation/cost analysis:** Cost analysis<br>**Perspective of analysis:** Not defined.<br>**Currency and cost year:** US dollars for cost year 1999.<br>**Discounting:** Discounting not performed<br>**Sensitivity analysis:** Not conducted. | **Outcome: Costs and severity of illness (the Palliative Performance Scale (PPS) was used to measure severity of illness).**<br>**Types of costs measured:** Healthcare resource use including emergency department, physician office, and home health and palliative visits, hospital and skilled nursing facility. | **Base case results:** Cancer patients enrolled in the palliative care group spent \$5,936 less on average as compared to those in usual care, amounting to a 35% reduction in average service costs. Patients diagnosed with COPD enrolled in the palliative care group, controlling for days on service and severity of illness, spent \$11,325 less on average as compared to those in usual care, amounting to a 67% decrease in the cost of care. Patients diagnosed with CHF enrolled in the palliative care group spent \$8,445 less on average as compared to those in usual care, amounting to a 52% decrease in the cost of care. Among all diseases, patients enrolled in palliative care were more likely to die at home. Enrolment in palliative care was significant associated with cost reductions for patients with cancer, COPD, and CHF. No significant difference was found between diagnostic groups in terms of magnitude of cost savings.<br>**Sensitivity analysis results:**<br>None. |
| Gage et al. 2015 (England) (Gage et al., 2015)<br>**Aim:** To (i) compare the characteristics of Rapid Response Service (RSS) users and non-users, (ii) explore differences in the proportions of users and non-users dying in the place of their choice, (iii) monitor the whole system service utilisation of users and non-users and compare costs. | **Intervention/care (specialist or generalist palliative care):** Generalist<br>**Intervention/care recipients and sample size:** All hospice patients who died with a preferred place of death. 668 participants were eligible for the Rapid Response service (RRS). 247 RRS users were compared with 441 non-users.<br>**Setting:** Community – rapid response service delivered at home.<br>**Intervention/care deliverers (e.g., professionals or paid carers):** Delivered by a team of experienced healthcare assistants (band 3), who were trained by the hospice and supported by the full hospice multidisciplinary team.<br>**Timing and duration:** RRS was rolled out sequentially to the three areas, starting January 2010, with six months between the start of provision in each area. The study continued for six months after the RRS was introduced in the third area (total of 18 months).<br>**Intervention/model of care description**<br>Palliative rapid response service (RRS) provided by a large hospice provider. The team responds rapidly 24/7 to crises in patients' homes (including care homes). Patients' needs and prognosis, and family circumstances are assessed, including patient/ family preferences. Hand-on care is provided in coordination with other community service. | **Dates of data collection:** Study began in January 2010 and continued for a total of 18 months.<br>**Length of follow-up:** NA.<br>**Type of economic evaluation/cost analysis:** Cost analysis<br>**Perspective of analysis:** NHS perspective<br>**Currency and cost year:** GBP for cost year 2010<br>**Discounting:** No discounting as time horizon less than 12 months.<br>**Sensitivity analysis:** No. | **Outcome:** Costs and preferred place of death. Demographic, preferences for place of death were obtained from hospice records.<br>**Types of costs measured:** Resource utilisation. General practitioners; community services; acute (hospital) services (A&E, inpatient nights, outpatient appointments, day hospital); Marie Curie home sitting; out-of-hours services (GP/nurse home visits, telephone advice, 'walk-in' attendances; social services received; hospice services, other than the RRS (home, outpatient, inpatient, day hospice). | **Base case results:** The median number of visits to people with different times in the study was similar (overall median: 11 visits; cost £425). There was no significant difference in the total service costs of users and non-users for any time period, except, amongst those referred to the hospice within 2 days of death, when RRS users had significantly higher overall cost of services than non-users due to the RRS input and other community care costs.<br>**Sensitivity analysis results:** None. |
| Goldhagen et al. 2016 (Goldhagen et al., 2016) | **Intervention/care (specialist or generalist palliative care):** Specialist | **Dates of data collection:** Study used unpublished evaluation study results from 2007. | **Outcome:** Costs and health related quality of life (HRQoL) was assessed but not | **Base case results:** Health related quality of life was generally high, and hospital charges per child declined by |
| (USA)<br>**Aim:** To determine the Programme's potential impact on HRQoL, and administrative data to assess the Programme's potential impact on hospital utilisation and costs. | **Intervention/care recipients and sample size:** End of life care to children with life threatening, complex chronic conditions and their families. Children with chronic complex medical conditions. enrolled in Community PedsCare through the years 2000 and 2006 were eligible for inclusion in the utilisation and cost study.<br>**Setting:** Community - Community-Based Paediatric Palliative Care (CBPPC)<br>**Intervention/care deliverers (e.g., professionals or paid carers):** Multidisciplinary professional delivered intervention.<br>**Timing and duration:** Patients enrolled in Community PedsCare (admissions range from 2002 to 2007) at the time of the study who had documented hospital admissions during the 2 years prior to and the first two quarters after enrolment in the programme between 2002–2006. Criteria for admission to PedsCare were broadly defined to include all chronic life-limiting conditions (with new diagnosis, change in status, complex situation) including children already enrolled in hospice.<br>**Intervention/model of care description**<br>CBPPC programme to provide comprehensive and compassionate palliative and end of life care to children with life threatening, complex chronic conditions and their families. The programme was designed to relieve suffering, provide comfort, and improve overall quality of life., providing community-based medical, nursing, social work, child life, spiritual and volunteer care. Services include pain and symptom management; medical consultation; mental health, psychosocial and spiritual support,<br>and counselling; family respite; assistance with financial issues and resource development; case management and care coordination; and bereavement and grief support. Special attention also paid to the needs of the siblings. | **Length of follow-up:** NA.<br>**Type of economic evaluation/cost analysis:** Cost analysis<br>**Perspective of analysis:** Not defined.<br>**Currency and cost year:** US dollars, costs were collected in quarterly sums for Quarters 3 and 4 of 2005 and Quarters 1 to 4 of 2006.<br>**Discounting:** Not required.<br>**Sensitivity analysis:** No. | incorporated within an economic evaluation.<br>**Types of costs measured:** Hospital utilisation costs - room and board, medical equipment and supplies, diagnostic costs, drug therapy, physical therapy, subspecialty institutional departments, pharmacy, dialysis, gastrointestinal services, and increment nursing. | \$1203 for total hospital services ($p = .34$) and \$1047 for diagnostic charges per quarter ($p = 0.13$) due to the CBPPC programme. Hospital length of stay decreased from 2.92 days per quarter to 1.22 days per quarter ($p < .05$).<br>**Sensitivity analysis:** None. |
| Gordon et al. 2022 (Gordon et al., 2022)<br>(USA)<br>**Aim:** The aim of this study was to evaluate an adult home palliative care (HPC) programme for multiple insurance product lines using multiple vendors to determine if the annual costs of healthcare decreased for those enrolled in HPC. | **Intervention/care (specialist or generalist palliative care):** Specialist<br>**Intervention/care recipients and sample size:** In the calendar year 2019, 506 members were referred to and qualified for health plan sponsored HPC. Of the 506 referrals, 396 were enrolled in the programme and 110 voluntarily refused, did not qualify, or were unable to be reached.<br>**Setting:** Community – home-based palliative care<br>**Intervention/care deliverers (e.g., professionals or paid carers):** 13 palliative care vendors to deliver care. Vendors utilise physicians, physician extenders, nurses, social workers, clergy, and coordinators.<br>**Timing and duration:** The key indications for eligibility for palliative care are severe progressive disease, using the hospital and emergency department to manage their illness, and the need for an extra layer of support.<br>**Intervention/model of care description**<br>The health maintenance organisation (HMO) health plan established the palliative care programme in 2014. This homebased palliative care programme has since expanded to serve Medicaid, Medicare, and commercial members in 38 California counties. The health plan administers this programme with the support of two medical directors and one manager who have other<br>areas of responsibility as well. Operationally, five, full-time, palliative care health plan nurses identify potential candidates through data mining, outreach, and engaging other health plan nurses who are involved in concurrent review, transition care management, and case management. | **Dates of data collection:** Resource use costs for calendar year 2019<br>**Length of follow-up:** NA. Cost savings calculated over 12-month period.<br>**Type of economic evaluation/cost analysis:** Cost analysis<br>**Perspective of analysis:** Payer perspective<br>**Currency and cost year:** US dollars for cost year 2019.<br>**Discounting:** No discounting as time horizon was 12 months.<br>**Sensitivity analysis:** Not conducted. | **Outcome: Costs and length of stay (1–5 versus 6–12 months).**<br>**Types of costs measured:** Inpatient, outpatient, and pharmacy costs. | **Base case results:** Overall medical costs for these 396 enrollees for the calendar year 2019 showed a gross savings of \$24,643 per member (16.7% decrease in cost). For members enrolled for 1–5 months, annual gross savings were \$23,314 per member (15.8% decrease from the comparison group), and for members enrolled for 6–12 months, annual gross savings were \$26,409 per member (17.9% decrease). The savings were most prominent for the commercial insurance product with a 51% decrease in annual costs.<br>**Sensitivity analysis results:** None. |
| Johnson et al. 2009 (Johnson et al., 2009)<br>(Canada)<br>**Aim:** To describe healthcare resource utilisation and costs of a pilot interdisciplinary healthcare model of palliative home care in Ontario, Canada. | **Intervention/care (specialist or generalist palliative care):** Specialist<br>**Intervention/care recipients and sample size:** There were 434 patients included in the pilot programme.<br>**Setting:** Community - interdisciplinary healthcare model of palliative home care<br>**Intervention/care deliverers (e.g., professionals or paid carers):** Palliative care team comprising of one palliative medicine physician, one palliative care coordinator, two palliative care nurses provided, and hospice and psychosocial spiritual support group or volunteers from any of five community hospices.<br>**Timing and duration:** Cancer patients who were expected to die within 12 months after the programme began, and/or were experiencing unmet symptom control and support needs because of an advanced life-threatening illness.<br>**Intervention/model of care description**<br>The pilot programme offered in-home care 24 h a day, 7 days a week, and incorporated both direct and indirect services, community outreach, flexible hours, and consultation with allied professionals and home care community providers. | **Dates of data collection:** 1 October 2000 to 31 October 2001.<br>**Length of follow-up:** NA.<br>**Type of economic evaluation/cost analysis:** Cost analysis.<br>**Perspective of analysis:** Healthcare system perspective.<br>**Currency and cost year:** Canadian dollars for cost year 2007.<br>**Discounting:** NA. Discounting not required as time horizon for cost analysis was less than 12 months.<br>**Sensitivity analysis:** Not conducted. | **Outcome:** Costs and length of stay broken down by disposition (discharged, alive, death).<br>**Types of costs measured:** Costs of formal hospice at home care. | **Base case results:** There were 434 patients included in the pilot programme. Total costs were approximately CAN\$2.4 million, and the cost per person amounted to approximately CAN\$5586.33 with average length of stay equal to over 2 months (64.22 days).<br>**Sensitivity analysis results**<br>None reported. |
| Klinger et al., 2013 (Klinger et al., 2013)<br>(Canada)<br>**Aim:** To assess resource utilisation and costs of a shared-care demonstration project in rural Ontario (Canada) from the public healthcare system's perspective. | **Intervention/care (specialist or generalist palliative care):** Generalist with support from specialist palliative care team.<br>**Intervention/care recipients and sample size:** Of the 95 study participants (average age 71 years), 83 had a cancer diagnosis (87%); the non-cancer diagnoses (12 patients, 13%) included mainly advanced heart diseases and Chronic Obstructive Pulmonary Disease (COPD).<br>**Setting:** Community – home-based palliative care<br>**Intervention/care deliverers (e.g., professionals or paid carers):** Primary care teams (Including visiting nurses and family physicians) and a palliative care consultant/team.<br>**Timing and duration:** All patients enrolled with a local primary care physician/Family Health Group with a survival prognosis of up to one year.<br>**Intervention/model of care description:** To strengthen primary care physicians' ability to deliver quality palliative home care and to meet the complex needs of end of life patients and their families, the Niagara West End of life Care Project was designed to provide enhanced interprofessional home-based shared-care in a rural community setting. The project sought to optimise coherent patient care through collaboration of primary care teams – including visiting nurses and family physicians – and a palliative care consultant/team, with an emphasis on the exchange of expertise and knowledge alongside the coordination of services in line with the understood goals of care and fully covered in this demonstration project. | **Dates of data collection:** January 2005 to March 2006.<br>**Length of follow-up:** NA. No follow-up, costs collected over 15-month study period.<br>**Type of economic evaluation/cost analysis:** Cost analysis<br>**Perspective of analysis:** Healthcare system perspective<br>**Currency and cost year:** Canadian dollars for cost year 2007<br>**Discounting:** Discounting was not conducted.<br>**Sensitivity analysis:** Not conducted. | **Outcome:** Costs.<br>**Types of costs measured:** Patient-based resource utilisation and costing information from the EPCT, the CCAC and family physicians/Family Health Groups as well as emergency room (ER) visits and hospitalisations at the local community hospital. | **Base case results:** Community Care Access Centre and Enhanced Palliative Care Team-based homemaking and specialised nursing services were the most frequented offerings, followed by equipment/transportation services and palliative care consults for pain and symptom management. Total costs for all patient-related services (in 2007 \$CAN) were \$1,625,658.07 – or \$17,112.19 per patient/\$117.95 per patient day.<br>**Sensitivity analysis results:** None. |
| Maetens et al. 2019 (Maetens et al., 2019)<br>(Belgium)<br>**Aim:** To evaluate the impact of palliative home care support on the quality of care and costs in the last 14 days of life. | **Intervention/care (specialist or generalist palliative care):** All forms of palliative home care support (specialist and generalist)<br>**Intervention/care recipients and sample size:** 8837 people who received palliative home care support in the last 720 to 15 days of life matched 1:1 by propensity score to 8837 people who received usual care.<br>**Setting:** Community – home care support<br>**Intervention/care deliverers (e.g., professionals or paid carers):** Multidisciplinary palliative home care teams, home care nurses and physiotherapists.<br>**Timing and duration:** People who received palliative home care support in the last 720 to 15 days of life. Costs collected for the last 14 days of life.<br>**Intervention/model of care description**<br>The forms of palliative home care support available in Belgium: (1) a multidisciplinary palliative home care team, (2) palliative home care nursing or physiotherapy and (3) the allowance for palliative home care patients, available twice and meant for non-reimbursed palliative care-related costs. | **Dates of data collection:** A matched cohort study was conducted on all deaths in Belgium in 2012, using linked data from eight administrative databases.<br>**Length of follow-up:** NA. Costs collected for the last 14 days of life.<br>**Type of economic evaluation/cost analysis:** Cost analysis.<br>**Perspective of analysis:** Third-party and patient co-payment perspective.<br>**Currency and cost year:** Euros for cost year 2017.<br>**Discounting:** No discounting as time horizon was less than 12 months.<br>**Sensitivity analysis:** Yes, sensitivity analyses were performed in which each separate support type was selected as the basis for the exposure group. | **Outcome: Costs and appropriateness/quality of palliative home care support**<br>**Types of costs measured:** Inpatient costs included all specific intervention and medication costs in the hospital. Outpatient costs included all specific intervention and medication costs outside the hospital. | **Base case results:** After matching, those using palliative home care support had, compared with those who did not, more family physician contacts (mean 3.1 [SD=6.5] vs 0.8 [SD=1.2]), more chance of home death (56.2%vs13.8%; relative risk [RR]=4.08, 95%CI 3.86 to 4.31), lower risk of hospital admission (27.4%vs60.8%; RR=0.45, 95%CI 0.43 to 0.46), ICU admission (18.3%vs40.4%; RR=0.45, 95%CI 0.43 to 0.48) or ED admission (15.2%vs28.1%; RR=0.54,95%CI 0.51 to 0.57). Mean total costs of care were lower for those using palliative home care support (€3081 [95% CI €3025 to €3136] vs €4698 [95% CI €4610 to €4787]; incremental cost: –€1617 [p<0.001]). Palliative home care support use positively impacts quality of care and reduces total costs of care at the end of life in Belgium.<br>**Sensitivity analysis results:** Sensitivity analyses conducted on each palliative home care support type separately revealed no substantial differences in the impact on the quality indicator outcomes. |
| Pham and Krahn, 2014 (Pham & Krahn, 2014) | **Intervention/care (specialist or generalist palliative care):** Specialist. | **Dates of data collection:** NA. The primary cost-effectiveness analysis used data from 8 interventions identified from the end of life mega-analysis. | **Outcome: Costs and effectiveness measures** included days at home, percentage dying at home, | **Base case results:** Based on the cost-effectiveness analysis, the per-patient cost of providing in-home palliative |
| (Canada)<br>**Aim:** To evaluate the cost-effectiveness of end of life (EoL) care interventions included in the EoL care mega-analysis. | **Intervention/care recipients and sample size:** Home-based end of life care to n = 431,762 decedents from Ontario, Canada.<br>**Setting:** Multiple settings including community (in-home palliative care)<br>**Intervention/care deliverers (e.g., professionals or paid carers):** Healthcare professionals and home carers.<br>**Timing and duration:** End of life care patients.<br>**Intervention/model of care description:** Cost of end of life care interventions in Ontario, Canada. | **Length of follow-up:** NA. Economic evaluation used multiple data sources with varying follow-up periods.<br>**Type of economic evaluation/cost analysis:** Cost-effectiveness analysis<br>**Perspective of analysis:** Healthcare payer's perspective.<br>**Currency and cost year:** Canadian dollars for cost year 2013.<br>**Discounting:** No discounting as time horizon was less than 12 months.<br>**Sensitivity analysis:** Yes, using quality-adjusted life days (QALDs) as an outcome measure. | and quality-adjusted life days.<br>**Types of costs measured:** The expected costs and health outcomes for a cohort of decedents in their last year of life. Assumed that those with a palliative prognosis could be identified (and therefore targeted for end of life interventions) according to a pattern of receiving end of life care services (e.g., physician billings). | team care was estimated to be between \$1,700 and \$2,400.<br>**Sensitivity analysis results:** Mean QALDs for in-home palliative team care were slightly higher than usual care because patients with in-home palliative team care spent less time in the ED or hospital. Mean QALDs for caregivers were also slightly higher than usual care because the analysis included a decrement in QALY weight for caregivers when patients were cared for in the ED or hospital (i.e., decrement in QALY weight of not having a break from caregiving). |
| Rosato et al., 2021 (Rosato et al., 2021)<br>(Italy)<br>**Aim:** To assess the cost-effectiveness of a home-based palliative approach (HPA) for people with severe multiple sclerosis (pwSMS). To assess direct healthcare costs in this population. | **Intervention/care (specialist or generalist palliative care):** Generalist<br>**Intervention/care recipients and sample size:** Patients with severe multiple sclerosis (pwSMS), 78 randomised pwSMS and 76 analysed (50 HPA, 26 usual care).<br>**Setting:** Community – home-based palliative care.<br>**Intervention/care deliverers (e.g., professionals or paid carers):** Nurse [team leader], neurologist, psychologist, social worker.<br>**Timing and duration:** Participant inclusion criteria included the following - primary or secondary progressive course; Expanded Disability Status Scale (EDSS) score ≥ 8.0. | **Dates of data collection** Home-based palliative care approach was delivered over 6 months.<br>**Length of follow-up** 6-months<br>**Type of economic evaluation/cost analysis:** Cost-effectiveness and cost-utility analysis.<br>**Perspective of analysis:** NHS and a personal perspective.<br>**Currency and cost year:** Euros for cost year 2017.<br>**Discounting:** No discounting as time horizon less than 12-months. | **Outcome: Costs and changes in symptom burden** (Palliative care Outcome Scale-Symptoms-MS, POS-S-MS), and QOL. PwSMSs quality-adjusted life years (QALYs) were assessed weighting surviving time with European QOL Five Dimensions (EQ-5D-3L).<br>**Types of costs measured:** Healthcare resource use – hospitalisations, home rehabilitation, healthcare professional costs, hospital attendances. | **Base case results** Mean QALYs were close to zero, and the mean group difference was -0.006 (95% CI -0.057 to 0.044). The mean baseline-adjusted cost difference was € -394 (95% confidence interval, CI -3,532 to 2,743). POS-S-MS cost-effectiveness showed a slight mean reduction of symptom burden (-1.9; 95% CI -1.1 to 5.0) with unchanged costs. Mean direct costs due to MS were €23,195 per year, almost equally distributed between NHS (€ 13,108) and the patients (€ 10,087). Personal care, medications and home rehabilitation accounted for |
|  | Other inclusion criteria were presence of a significant other; $\geq 2$ unmet care needs, or the pwSMS declared for comfort care only; one or more prognostic indicators (significant complex symptoms/medical complications, dysphagia/poor nutritional status, communication difficulties).<br>**Intervention/model of care description**<br>The home-based palliative care approach team made a comprehensive assessment of the needs of the pwSMS. HPA content was then agreed on, discussed with the pwSMSs caring physician, and delivered over six months. The intervention was not intended to replace existing services. | **Sensitivity analysis:** Yes. | PwSMS out of pocket costs. The HPA programme costs (not included in the cost of illness analysis) were also assessed, including the teams' home visits and meetings. Teams' travel costs and HPA programme development costs (training and cross-training of the teams) were not considered. | 80% of total expenditures. Most personal care costs were covered by patients, and these costs were 3/4 of patient out-of-pocket.<br>**Sensitivity analysis results:**<br>The sensitivity analysis produced consistent cost estimates. |
| Spiro et al., 2020 (Spiro et al., 2020)<br>(England)<br>**Aim:** To assess the cost of visit-based community care based around a 24/7 hospice-at-home (HatH) night service in the last 3 months of life. | **Intervention/care (specialist or generalist palliative care):** Generalist<br>**Intervention/care recipients and sample size:** Between June and August 2017, 27 cost diaries were distributed to accepting family carers and patients, and 13 returned (48%), making 30 diaries available for analysis.<br>**Setting:** Community – hospice-at-home service<br>**Intervention/care deliverers (e.g., professionals or paid carers):** Clinical staff<br>**Timing and duration:** Costs were collected for up to two weeks in the last three months of life.<br>**Intervention/model of care description**<br>The data from this study derive from an evaluation of the quality and effectiveness of a Hospice-at Home (HatH) night service. The service is a charitable organisation providing 24-hour care for parts of the Thames Valley and East of England, UK. In 2017, clinical<br>staff visited and cared for 1,760 patients and their families at home. Care provision included medical, technological, and nursing care, and provided for the emotional, practical, and informational needs of patients and family members, with 85% of clients who wished to die at home achieving their wish. | **Dates of data collection**<br>July–December 2016<br>**Length of follow-up**<br>NA, costs collected for up to 2-weeks in the last 3-months of life.<br>**Type of economic evaluation/cost analysis**<br>Pilot cost analysis<br>**Perspective of analysis:** Not defined.<br>**Currency and cost year:** GBP for cost year 2016.<br>**Discounting:** No discounting as time horizon less than 12-months.<br>**Sensitivity analysis:** Not conducted | **Outcome:** Service use captured by families completing a health and social care diary of at-home visits over two-weeks following contact with the HatH night service.<br>**Types of costs measured:** Day-to-day costs of hospice-at-home based services including all health-care visits made daily by General Practitioner; District Nurse; Specialist Nurse; Hospice Nurse; Marie Curie/MacMillan; Social Services/Formal Carers; and 'Others,' which included visits from chiropodists, occupational therapist, and physiotherapists, to capture all other professionals. | **Base case results:** Diaries captured 333 days of care provision, averaging 11 diary days per family, 708 healthcare professional and carer visits, lasting 604 hours at a cost of £20,192 (\$24,946). This can be calculated as average costs of £28.5 (\$35.2) per visit, £60.6 (\$74.8) per day, and £673.1 (\$831.6) per person over an average of 11 days. The lowest cost of care for an individual reported by the diaries was £33.2 (\$41.0) for one visit prior to a patient's death, and the highest care for an individual was £2,276.3 (\$2,812.3) over 20 days and 162 visits.<br>**Sensitivity analysis results:**<br>None. |

**Table 15:**
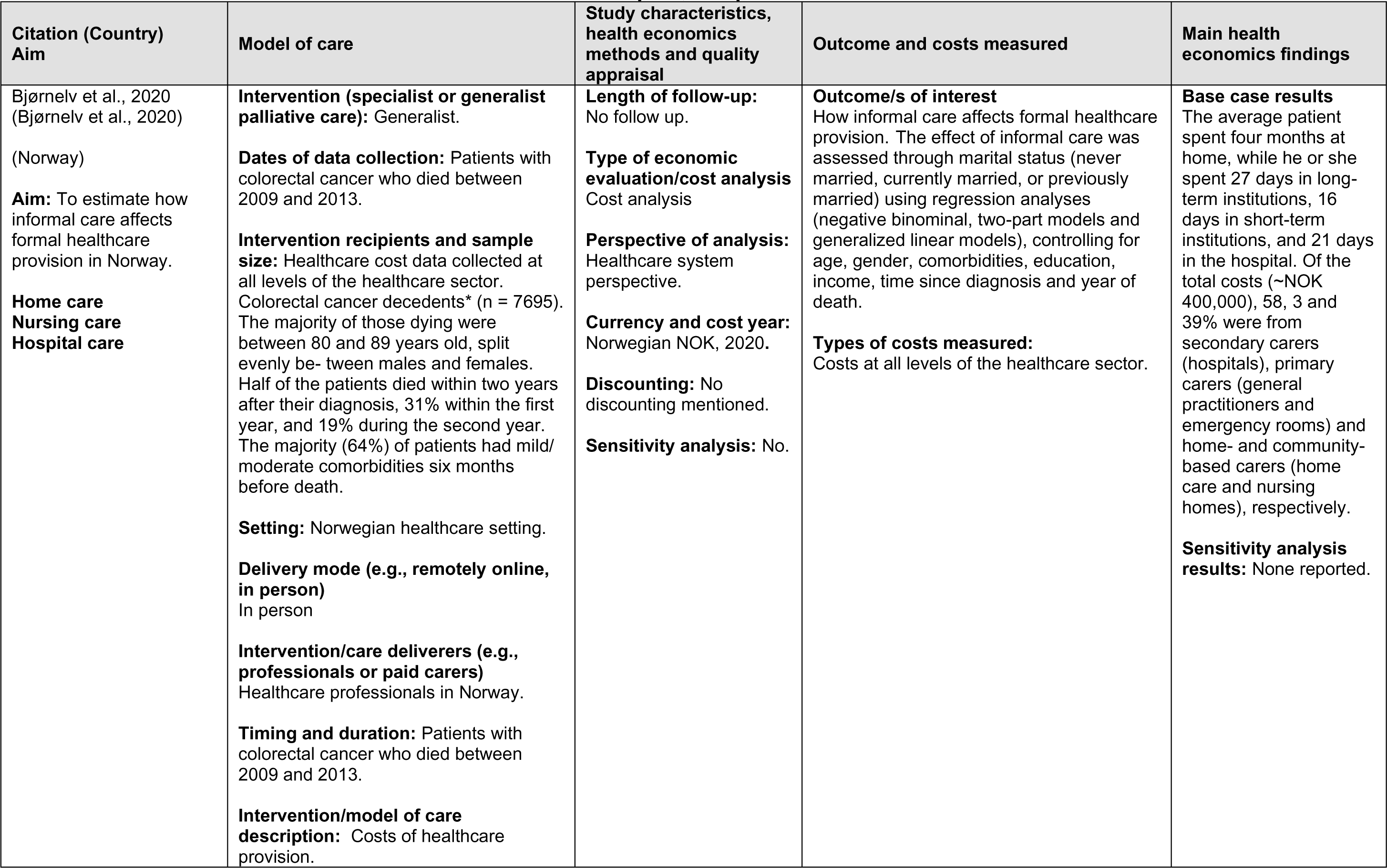
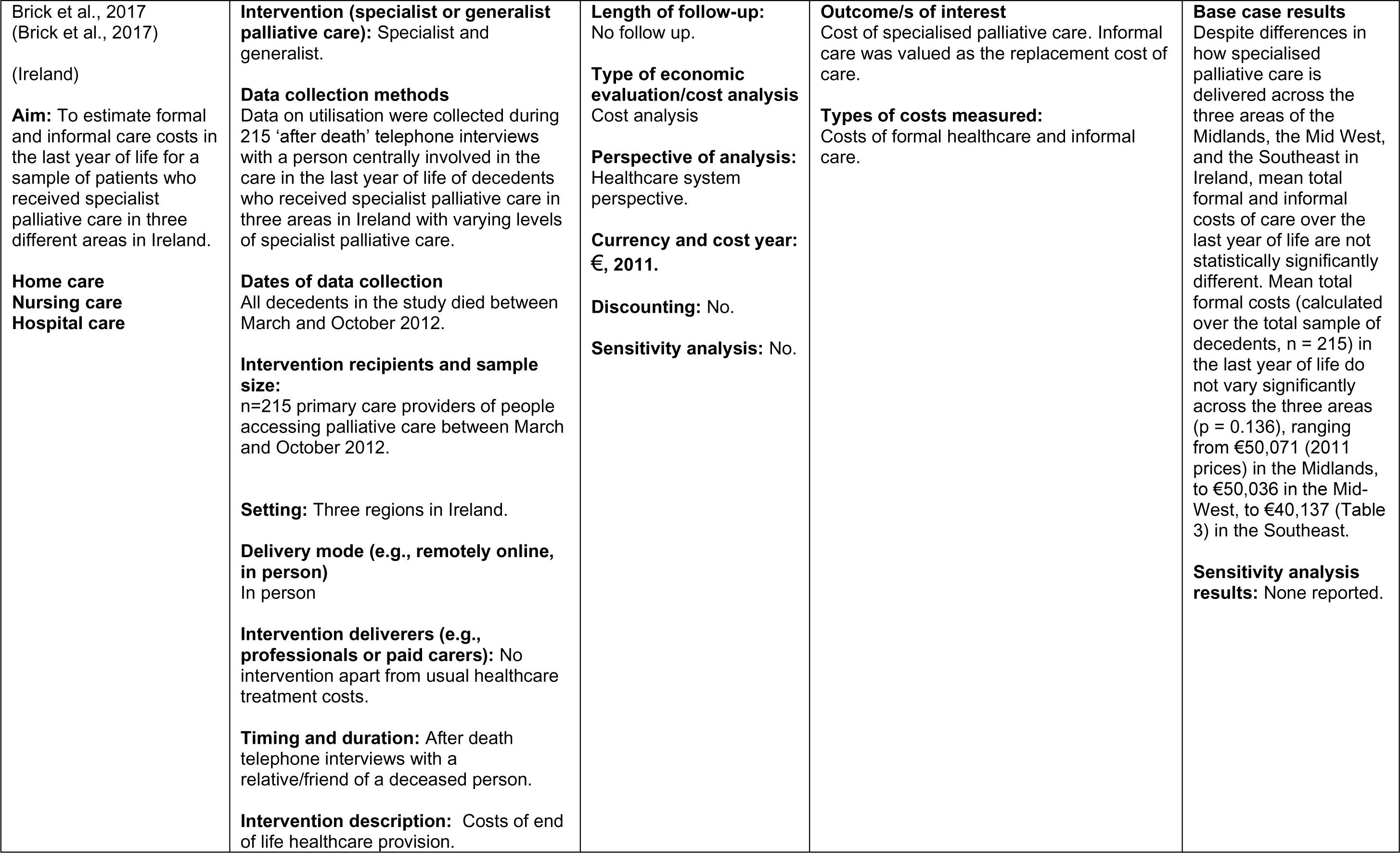
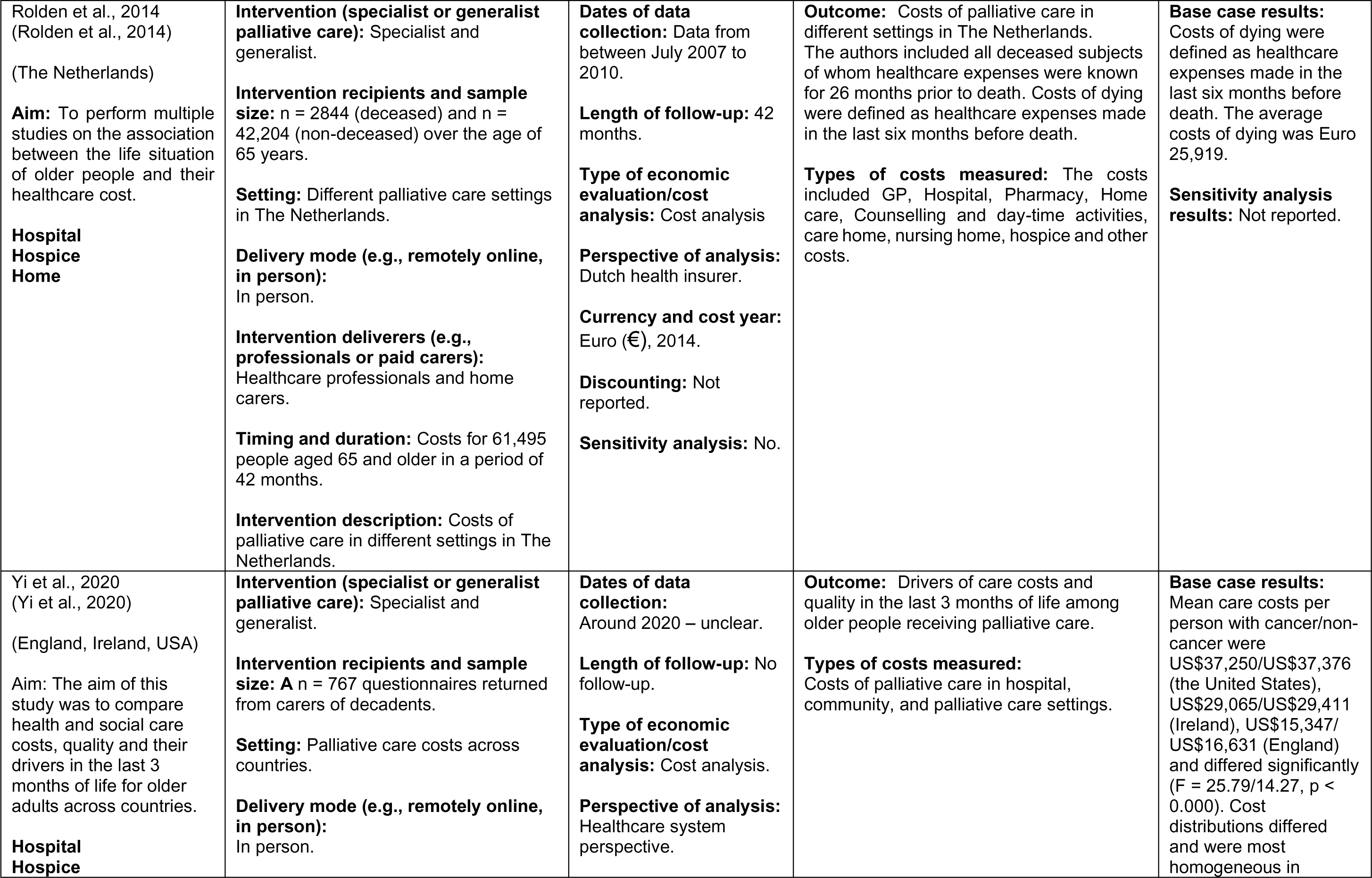
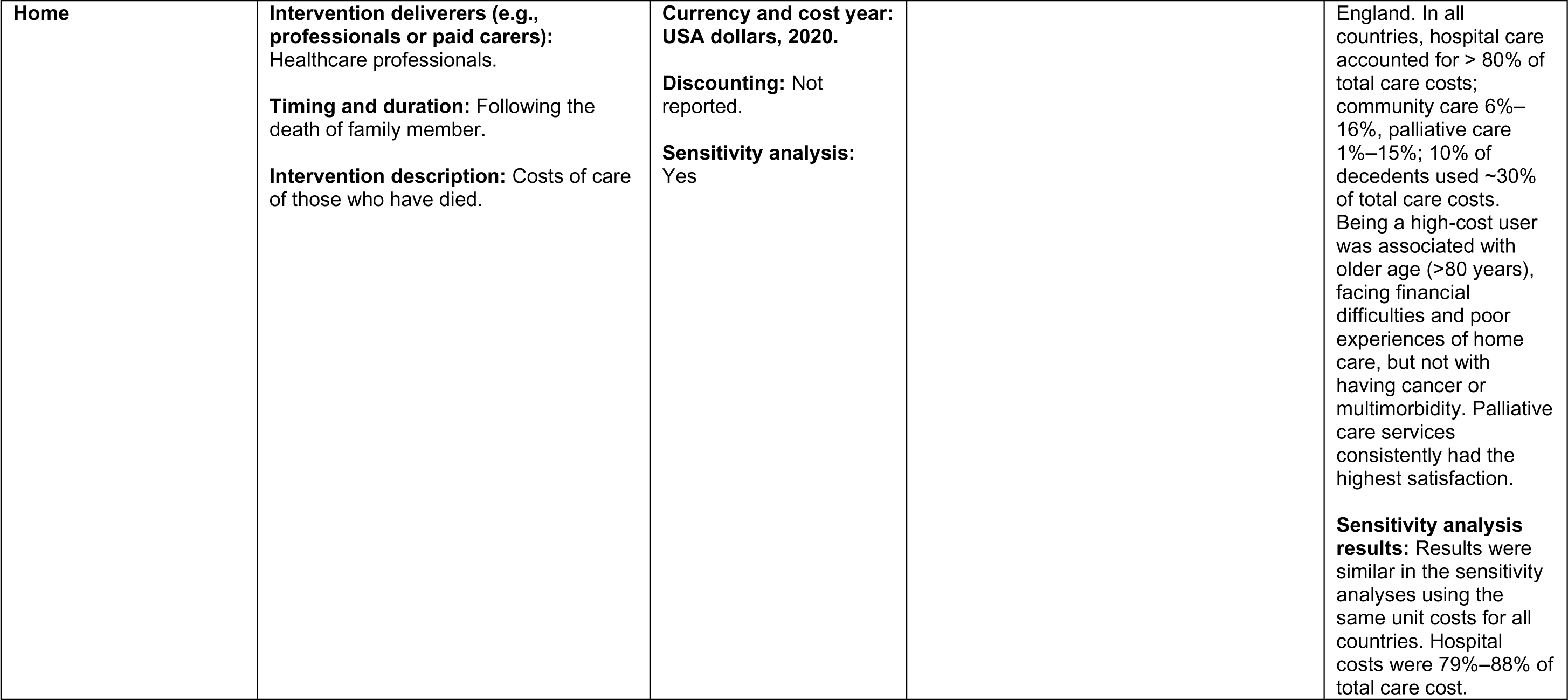
Evidence of costs from combined studies - Hospital, Hospice and Home.

**Table 16:** Evidence of costs from combined studies - Hospital and Hospice.

| Citation (Country)<br>Aim | Model of care | Study characteristics,<br>health economics<br>methods and quality<br>appraisal | Outcome and costs measured | Main health<br>economics findings |
| --- | --- | --- | --- | --- |
| Hoverman et al., 2020<br>(Hoverman et al., 2020)<br>(USA)<br>**Aim:** To measure and characterize the total cost of care for those who received less than three days of hospice care (HC) at the end of life compared with those who received three days or more.<br>**Hospital Hospice** | **Intervention (specialist or generalist palliative care):** Specialist<br>**Study design**<br>Economic cost study of hospital and hospice based palliative care for oncology patients.<br>**Dates of data collection**<br>The data from the first two years of the programme (2016–2018) on 7329 patients are presented here to display the costs of care in the last six months of life, and especially in the last 30 days.<br>**Intervention recipients and sample size:**<br>**Participants and sample size**<br>Oncology patients. (There were 7329 deaths).<br>**Setting**<br>USA Healthcare setting.<br>**Delivery mode (e.g., remotely online, in person)**<br>In person<br>**Intervention/care deliverers (e.g., professionals or paid carers):**<br>Healthcare professionals.<br>**Timing and duration:** Patients who had died between 2016 and 2018.<br>**Intervention/model of care description:** | **Study design:** Cost analysis.<br>**Length of follow-up:**<br>No follow up.<br>**Type of economic evaluation/cost analysis**<br>Cost analysis.<br>**Perspective of analysis:**<br>Healthcare providers.<br>**Currency and cost year:**<br>USA dollars, 2020.<br>**Discounting:** No.<br>**Sensitivity analysis:** No. | **Outcome/s of interest**<br>Medicare cost per site of care including hospice, hospital/intensive care unit [ICU], ICU/hospital, and emergency room visits.<br>**Types of costs measured:**<br>Costs of formal hospital care and hospice care. | **Base case results:** The Dying in the hospital was twice the cost of dying at home (\$20,113 vs. \$10,803.<br>The average final 30 day spend: dying at hospital was \$22,410, but more if a person died in ICU \$28,301. Dying at a Skilled Nursing Facility was \$19,400. Dying hospice (medical) was \$17,418. Dying hospice at home was \$10,098.<br>**Sensitivity analysis results:** None reported. |
|  | No interventions, just cost of different settings of care including (hospice, hospital/intensive care unit [ICU], ICU/hospital, and emergency room visits). |  |  |  |
| Saygili and Celik, 2019 (Saygili & Çelik, 2019)<br>(Turkey)<br>**Aim:** to evaluate the cost-effectiveness of three alternative palliative care models for cancer patients<br>**Hospital Hospice Home** | **Intervention (specialist or generalist palliative care):** Specialist and generalist.<br>**Intervention recipients and sample size:** n = 160 patients diagnosed with cancer (CPCC:60, HIS:59, HHC:41).<br>**Setting:** Different palliative care settings in Turkey.<br>**Delivery mode (e.g., remotely online, in person):** In person.<br>**Intervention deliverers (e.g., professionals or paid carers):** Healthcare professionals and home carers.<br>**Timing and duration:** The study sample consisted of cancer patients receiving PC in three alternative models. It was thought that comparing patients with similar care needs and levels of dependency on caregivers was important in terms of measuring the cost and effectiveness of the care provided by alternative PC models. The level of care needed was determined by using the Rush-Medicus patient classification system and the calculated scores were used to decide which patients should be included in the sample. | **Dates of data collection:** The data was collected between February 2015 and August 2016.<br>**Length of follow-up:** No follow-up.<br>**Type of economic evaluation/cost analysis:** Cost-effectiveness<br>**Perspective of analysis:** Societal perspective.<br>**Currency and cost year:** USA dollars, 2016 (mid-year).<br>**Discounting:** No.<br>**Sensitivity analysis:** No. | **Outcome:** Costs of palliative care in different settings in Turkey. The patients' quality of life and their levels of satisfaction were used as the indicators of effectiveness, while direct and indirect costs incurred by service providers, patients and relatives were considered in estimating the costs of alternative models.<br>**Types of costs measured:** Costs of three models of palliative care in Turkey.<br>(a) comprehensive palliative care center (CPCC);<br>(b) hospital inpatient services (HIS); and<br>(c) home healthcare (HHC) services. | **Base case results:** From a societal perspective, palliative care services provided the hospital inpatient service model was found to be more cost-effective than the comprehensive palliative care centre (CPCC) model. From a patient perspective, home healthcare services was found to be more cost-effective compared to the other two models. The average indirect cost (\$164.10) for the patients receiving care from HISs was found to be the lowest compared with the indirect costs of HHCs (\$344.62) and CPCCs (\$778.43). Estimated ICERs indicated that HHC was more likely to produce a better quality of life at the cost of an additional \$33.43 per additional 1 QoL score when it was compared with HIS. However, HHC has a capability of producing a |
| | **Intervention description:** Costs of palliative care in different settings in Turkey. | | | better QoL score, and even reduces indirect costs (\$18.30) for patients with an additional 1 QoL score compared with CPCC.<br>**Sensitivity analysis results:** Not reported. |

**Table 17:**
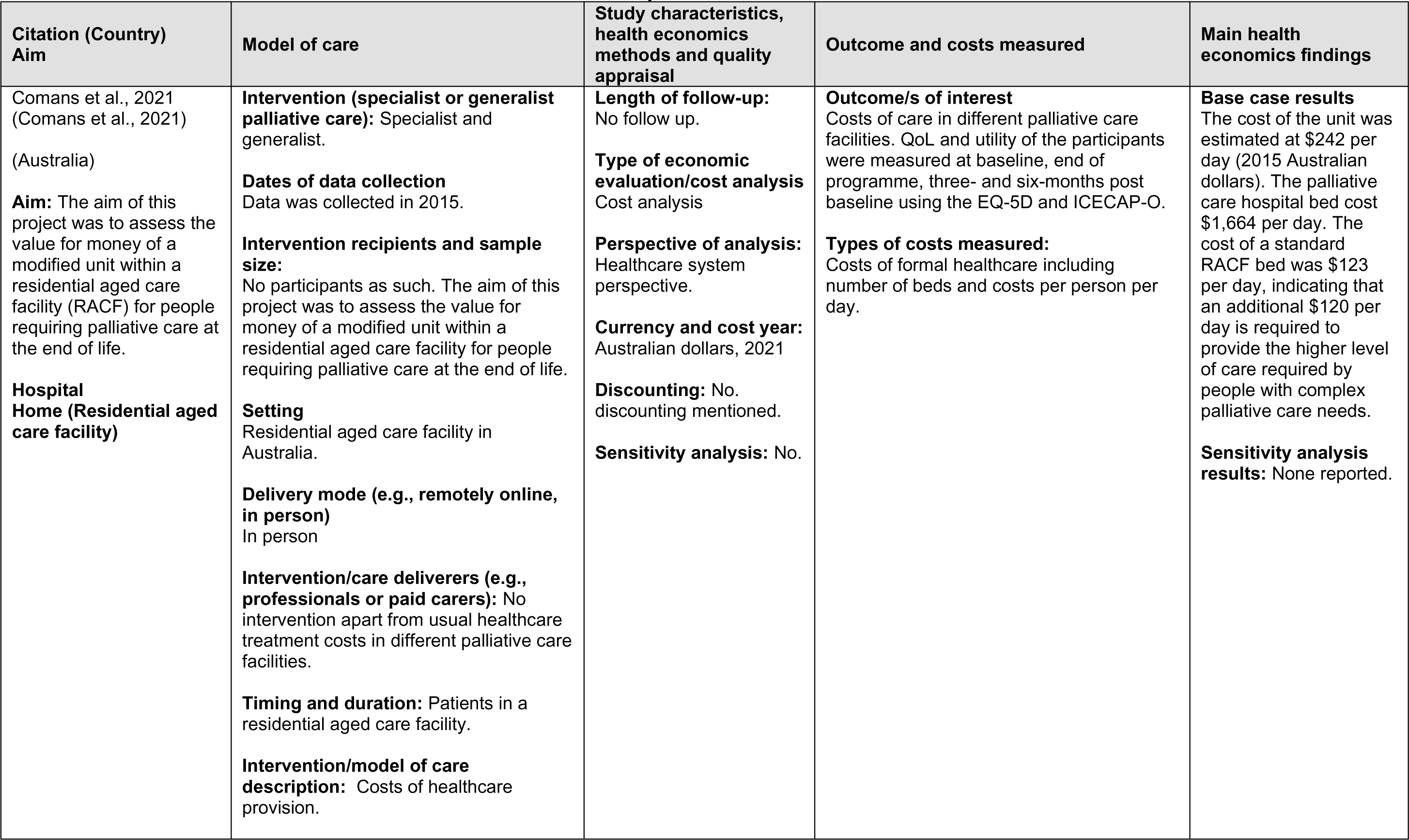
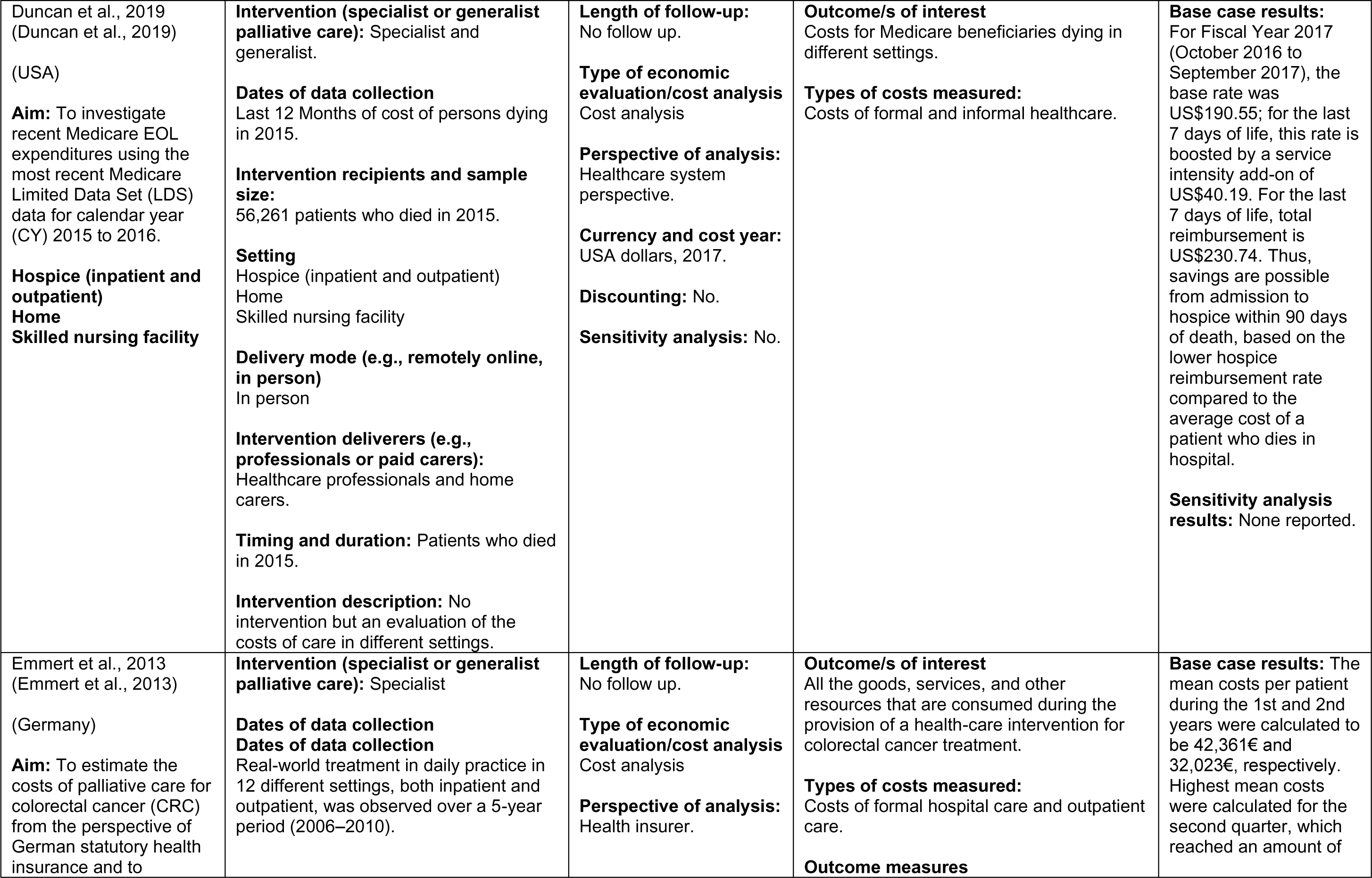
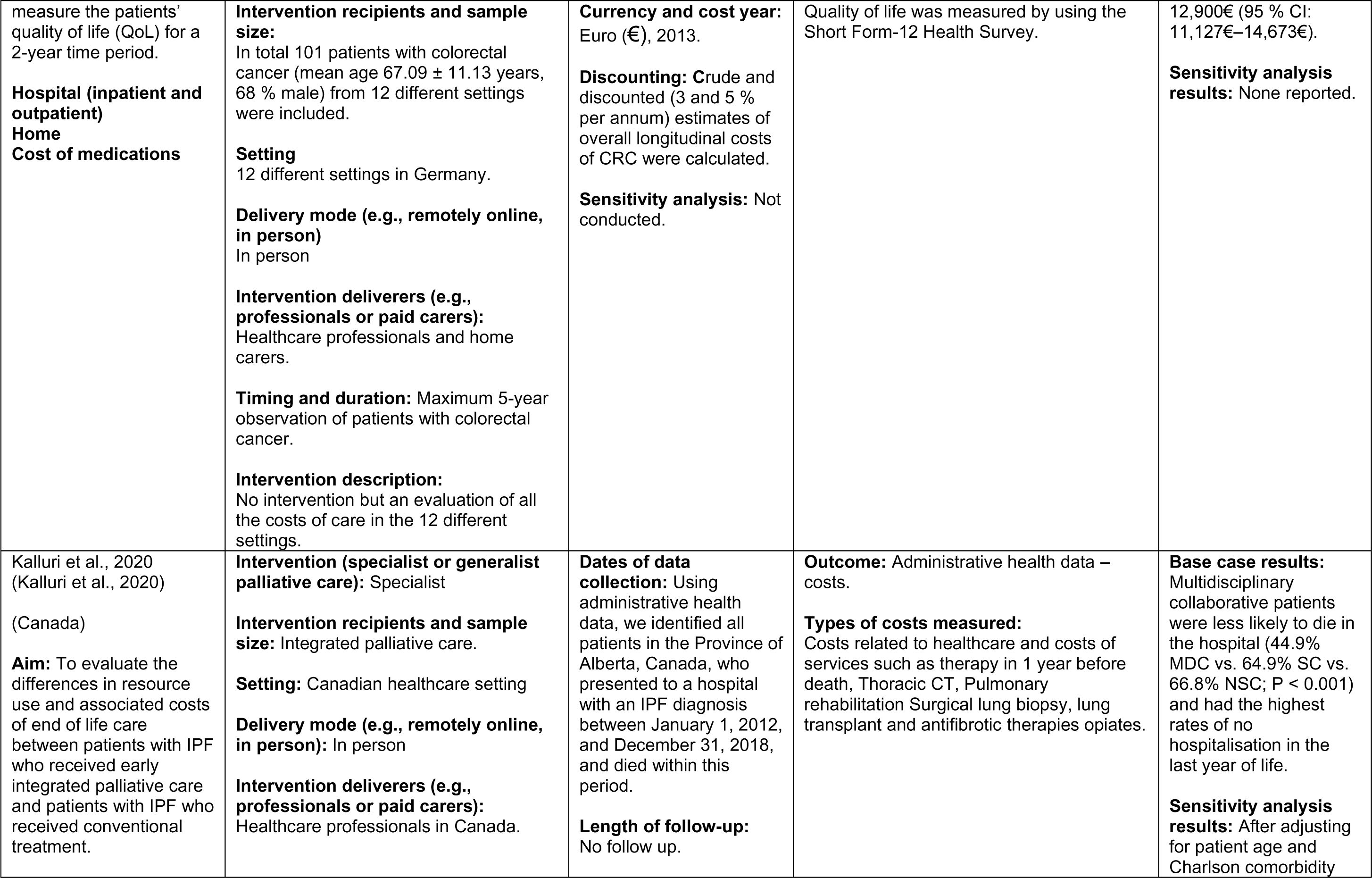
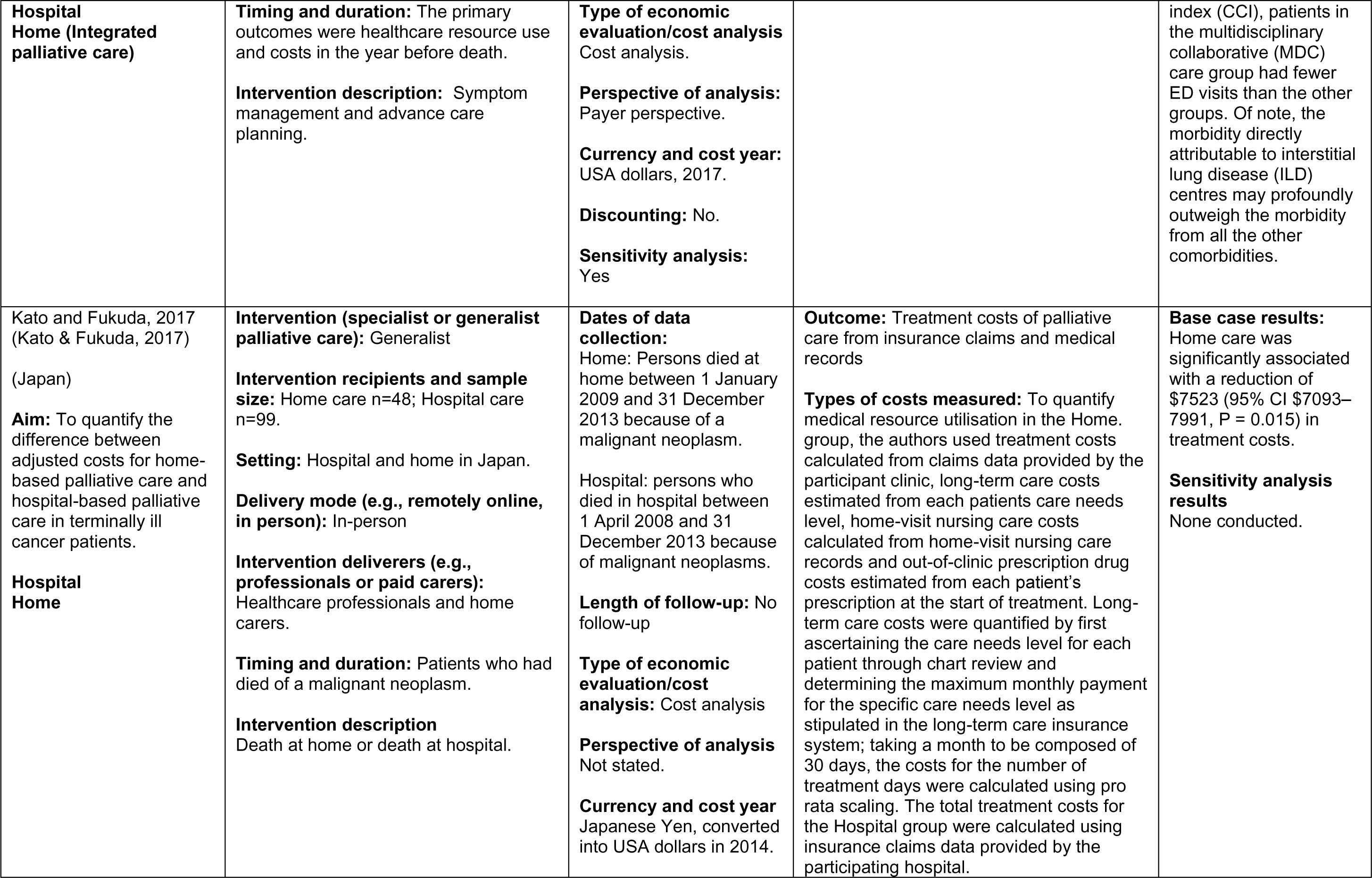
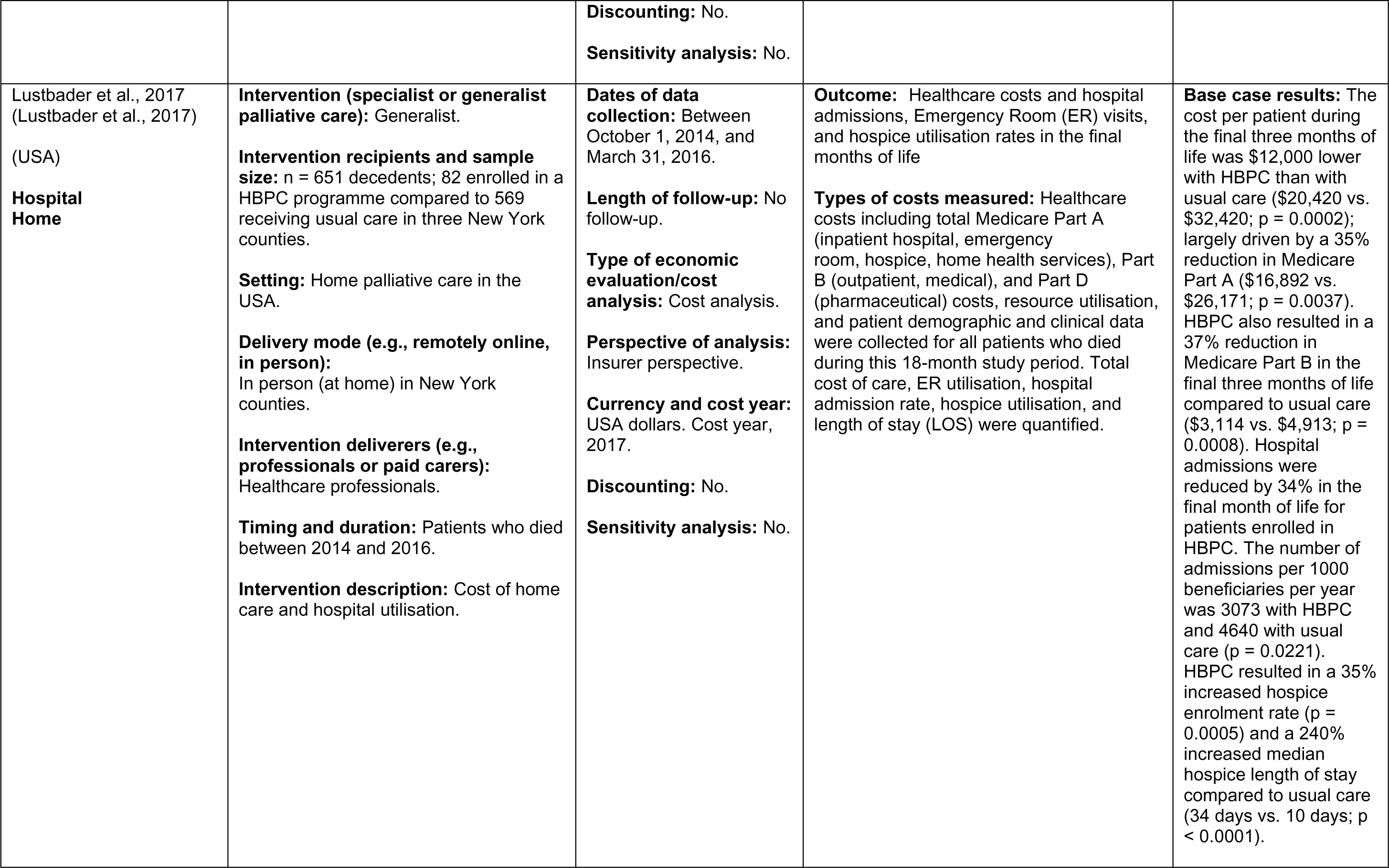
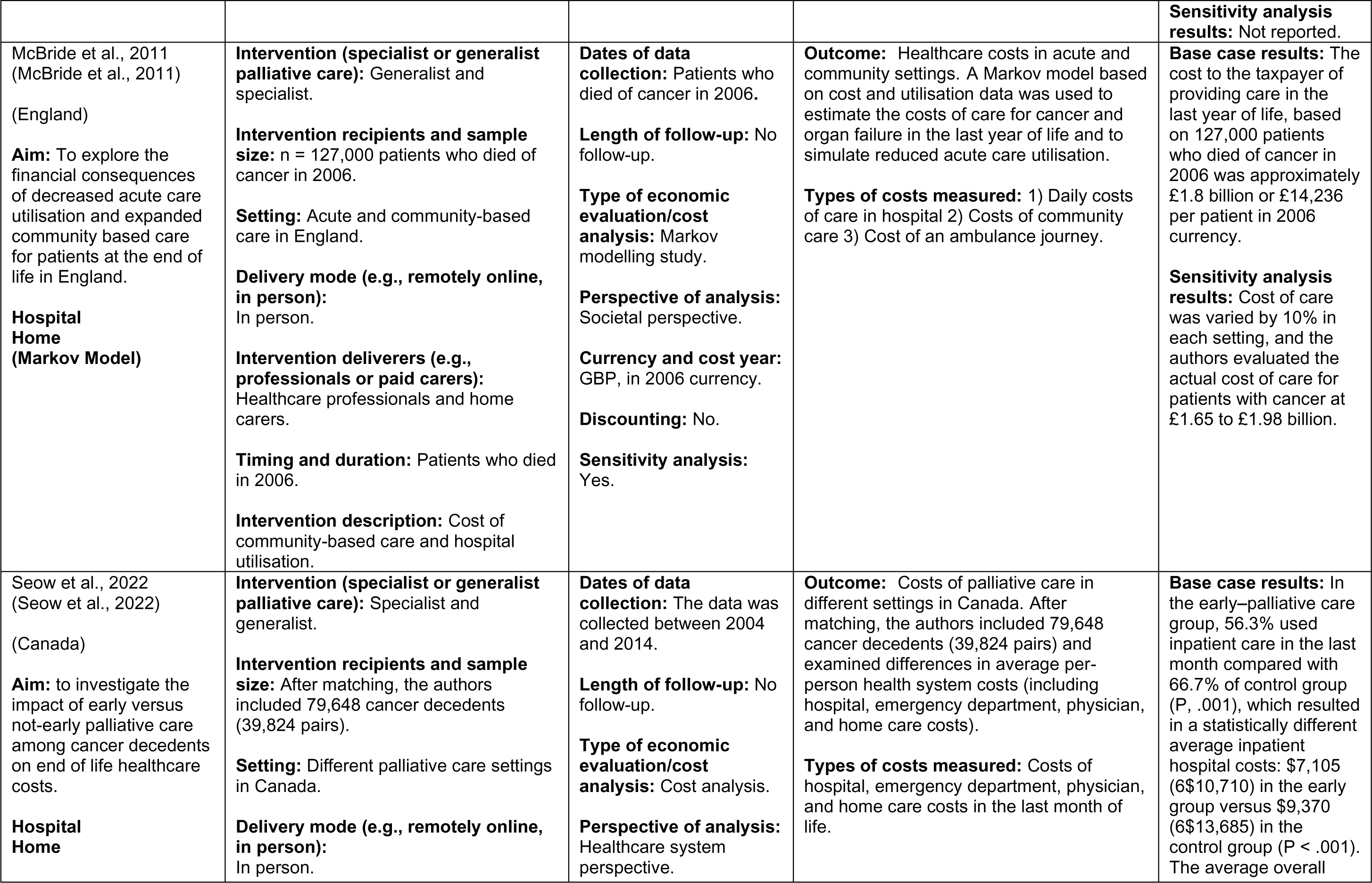
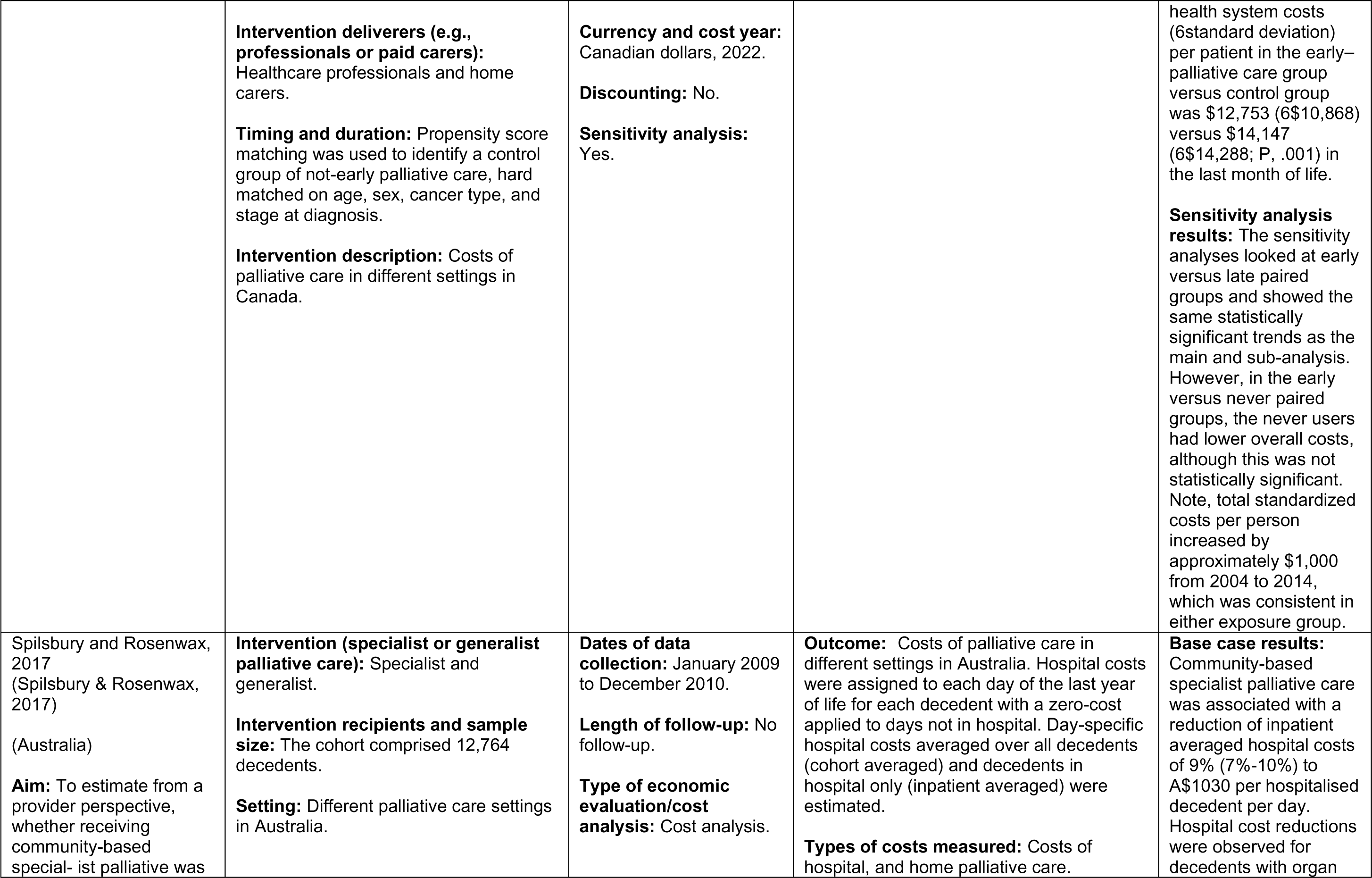
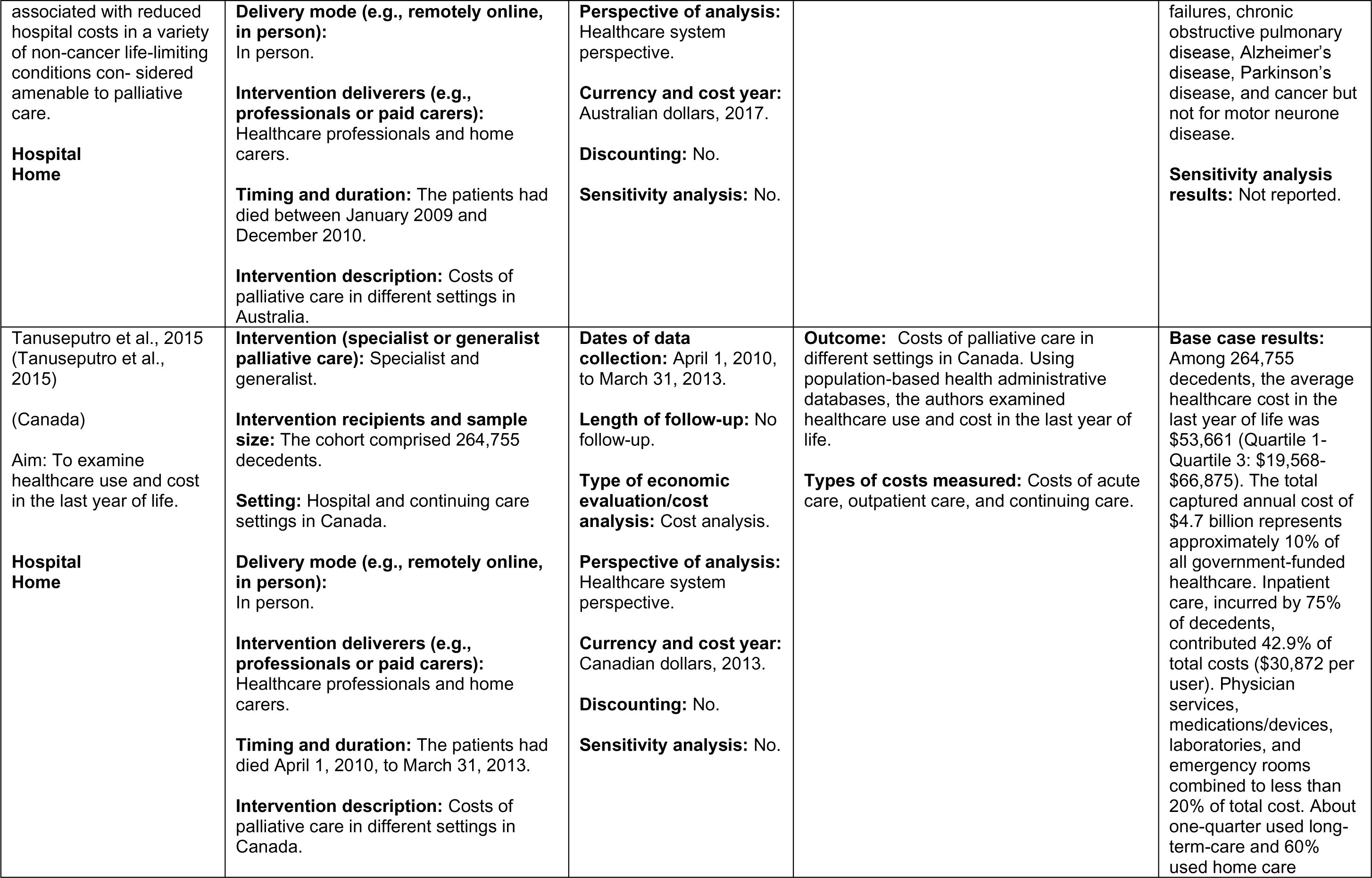
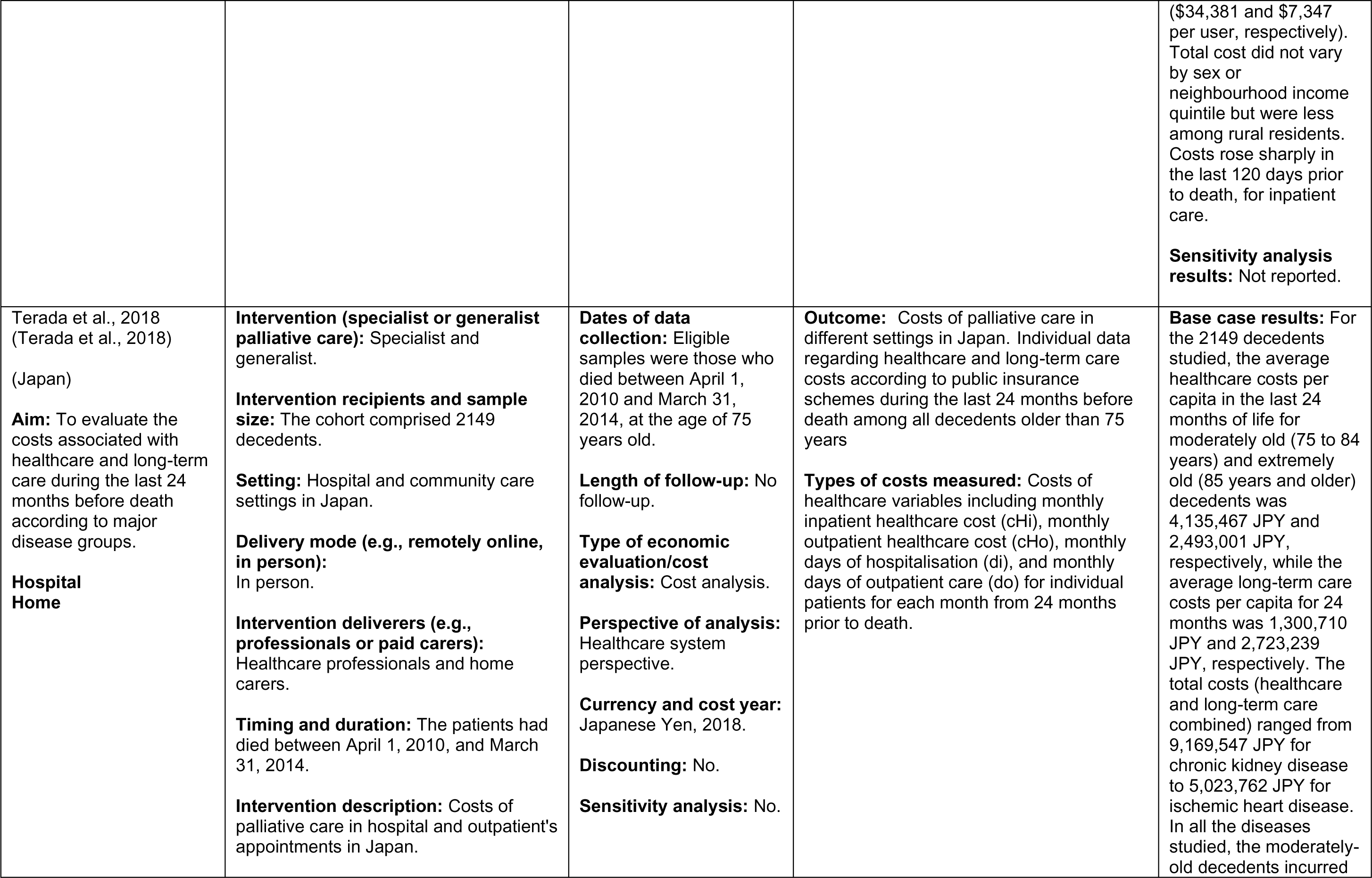
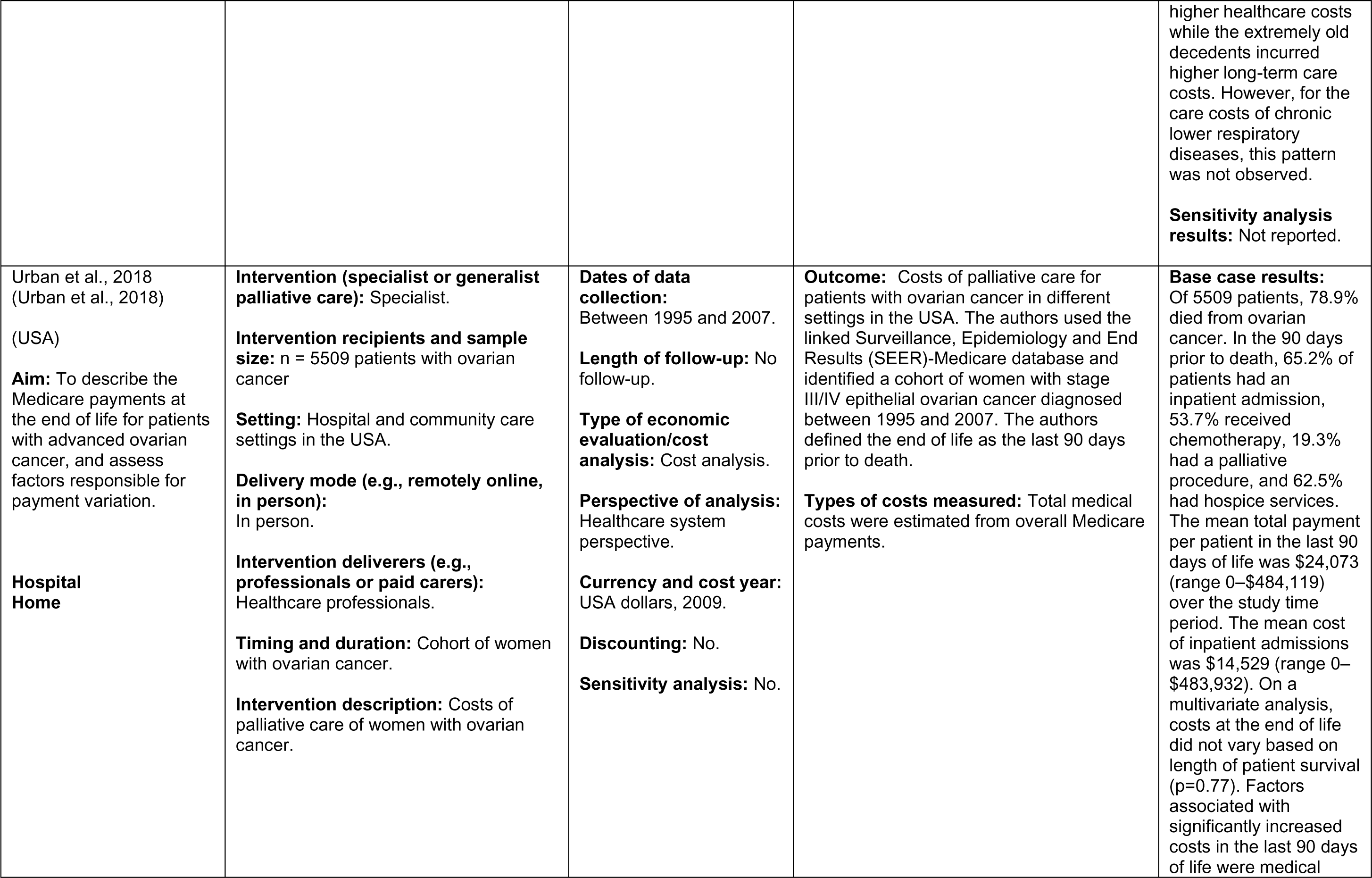
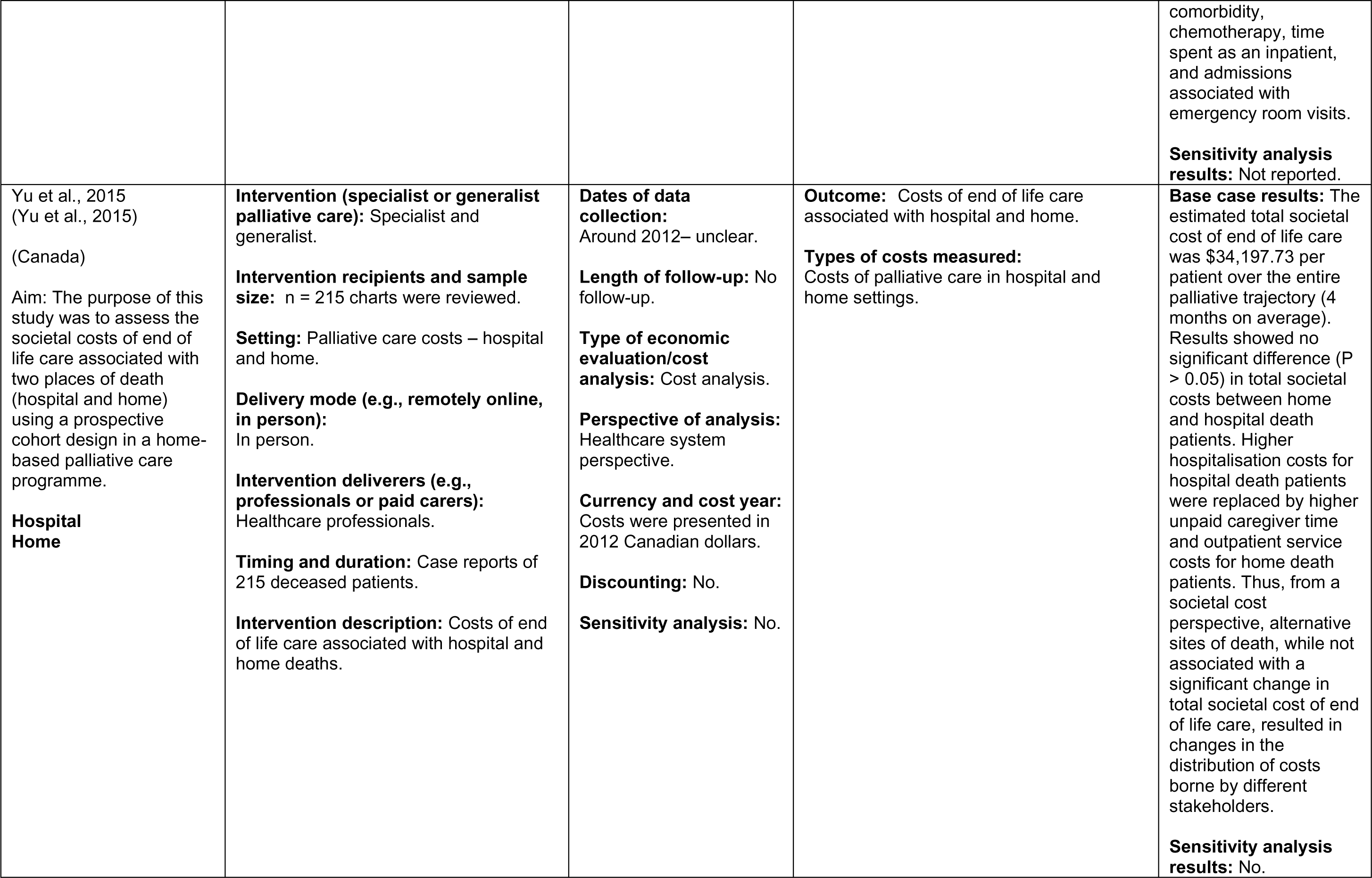
Evidence of costs from combined studies - Hospital and Home.

**Table 18:**
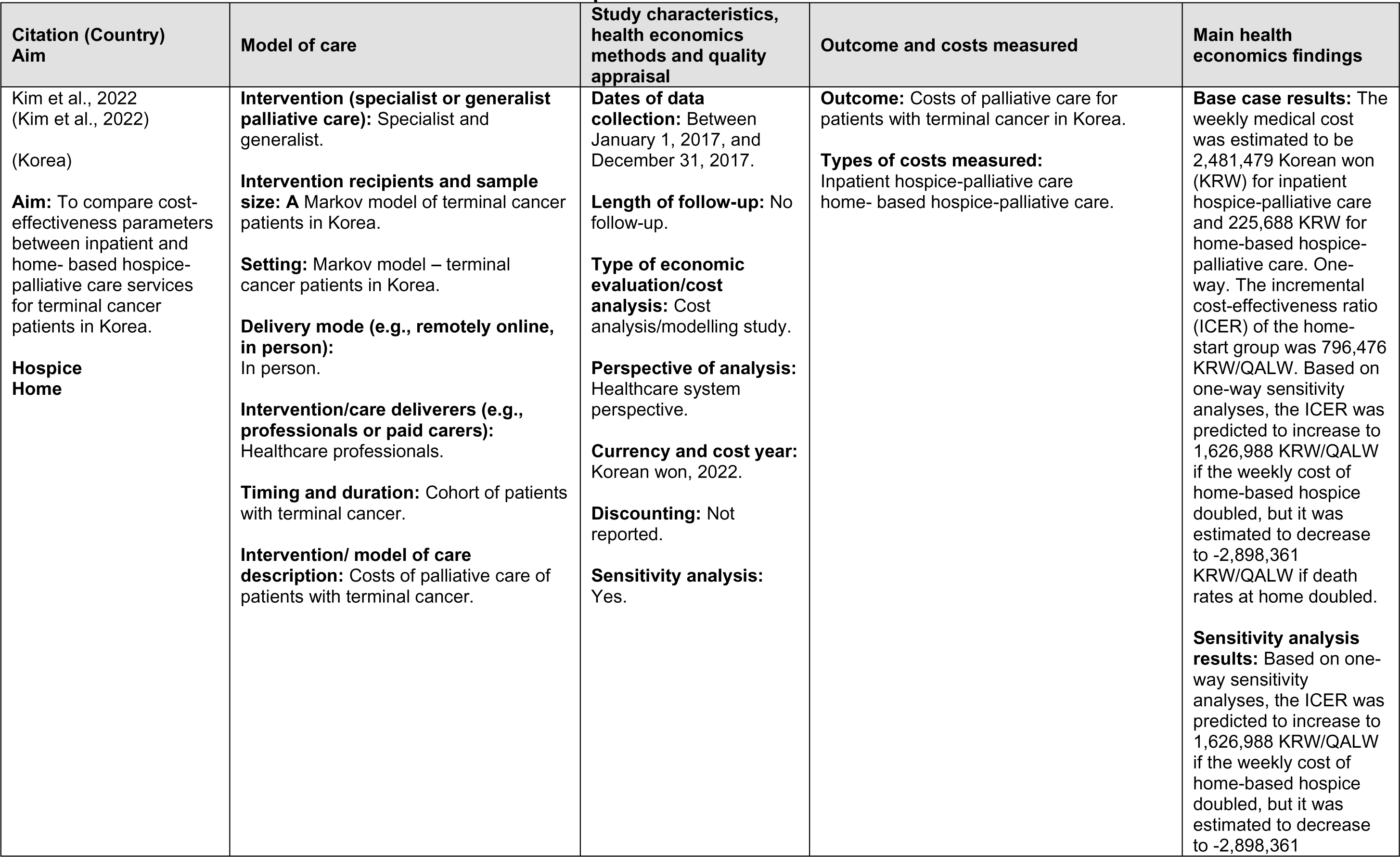
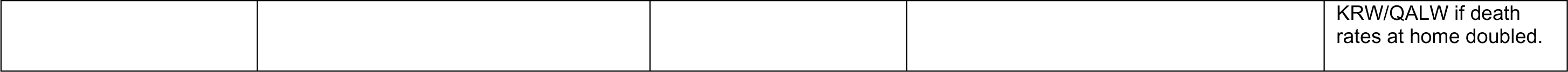
Evidence of costs from combined studies - Hospice and Home.

| Citation (Country)<br>Aim | Model of care | Study characteristics,<br>health economics<br>methods and quality<br>appraisal | Outcome and costs measured | Main health<br>economics findings |
| --- | --- | --- | --- | --- |
| Kim et al., 2022<br>(Kim et al., 2022)<br>(Korea)<br>**Aim:** To compare cost-effectiveness parameters between inpatient and home- based hospice-palliative care services for terminal cancer patients in Korea.<br>**Hospice**<br>**Home** | **Intervention (specialist or generalist palliative care):** Specialist and generalist.<br>**Intervention recipients and sample size:** A Markov model of terminal cancer patients in Korea.<br>**Setting:** Markov model – terminal cancer patients in Korea.<br>**Delivery mode (e.g., remotely online, in person):**<br>In person.<br>**Intervention/care deliverers (e.g., professionals or paid carers):**<br>Healthcare professionals.<br>**Timing and duration:** Cohort of patients with terminal cancer.<br>**Intervention/ model of care description:** Costs of palliative care of patients with terminal cancer. | **Dates of data collection:** Between January 1, 2017, and December 31, 2017.<br>**Length of follow-up:** No follow-up.<br>**Type of economic evaluation/cost analysis:** Cost analysis/modelling study.<br>**Perspective of analysis:** Healthcare system perspective.<br>**Currency and cost year:** Korean won, 2022.<br>**Discounting:** Not reported.<br>**Sensitivity analysis:** Yes. | **Outcome:** Costs of palliative care for patients with terminal cancer in Korea.<br>**Types of costs measured:**<br>Inpatient hospice-palliative care<br>home- based hospice-palliative care. | **Base case results:** The weekly medical cost was estimated to be 2,481,479 Korean won (KRW) for inpatient hospice-palliative care and 225,688 KRW for home-based hospice-palliative care. One-way. The incremental cost-effectiveness ratio (ICER) of the home-start group was 796,476 KRW/QALW. Based on one-way sensitivity analyses, the ICER was predicted to increase to 1,626,988 KRW/QALW if the weekly cost of home-based hospice doubled, but it was estimated to decrease to -2,898,361 KRW/QALW if death rates at home doubled.<br>**Sensitivity analysis results:** Based on one-way sensitivity analyses, the ICER was predicted to increase to 1,626,988 KRW/QALW if the weekly cost of home-based hospice doubled, but it was estimated to decrease to -2,898,361 |
|  |  |  |  | KRW/QALW if death rates at home doubled. |

**Table 19:** Evidence of costs from combined studies - Enhanced supported care services.

| Citation (Country)<br>Aim | Model of care | Study characteristics,<br>health economics<br>methods and quality<br>appraisal | Outcome and costs measured | Main health<br>economics findings |
| --- | --- | --- | --- | --- |
| Nguyen et al., 2017<br>(Nguyen et al., 2017)<br>(Australia)<br>**Aim:** This study aims to evaluate the cost-effectiveness of delivering a nationwide Advance Care Planning programme within the Australian primary care setting.<br>**Primary care setting** | **Intervention (specialist or generalist palliative care):** Specialist.<br>**Intervention recipients and sample size:** Markov model of people aged 65+ years, who are at risk of dementia.<br>**Setting:** Australian primary care system.<br>**Delivery mode (e.g., remotely online, in person):** In person.<br>**Intervention/care deliverers (e.g., professionals or paid carers):** Healthcare professionals.<br>**Timing and duration:** Markov model of patients at risk of dementia.<br>**Intervention/model of care description:** Cost of an advanced care planning programme within the Australian primary care setting, | **Dates of data collection:** Markov model published in 2017.<br>**Length of follow-up:** No.<br>**Type of economic evaluation/cost analysis:** Markov modelling<br>**Perspective of analysis:** Healthcare system perspective.<br>**Currency and cost year:** All costs were calculated in 2015, Australian dollars.<br>**Discounting:** Both costs and outcomes were discounted at 5% in the base case and at 2% and 7% in the sensitivity analyses.<br>**Sensitivity analysis:** Yes. | **Outcome:** Cost-effectiveness of advanced care planning in Australia. Inputs for the model was sourced and estimated from the literature.<br>**Types of costs measured:** The cost of delivering advanced care planning in Australia compared to palliative care in hospital. | **Base case results:** The model indicated that if the cost per individual ACP reaches \$850 (equivalent to seven visits) then the programme is no longer cost-effective. Nonetheless, this scenario is unlikely because individual ACP has been provided successfully in Australia through group information sessions followed by 1–2 visits by trained ACP facilitators, which costs less than AU\$250. At this cost, the programme is more likely to be cost-effective than the base case scenario.<br>**Sensitivity analysis results:** Extensive sensitivity analyses, including threshold analyses, were conducted on the key parameters to assess the likelihood of ACP remaining cost-effective. The result was highly sensitive to several key parameters (see Table 1): ACP completion and compliance rates; and<br>dying choice (hospital versus non-hospital settings). The probability sensitivity analysis of 5000 Monte Carlo replications highlighted that there was a 50–50 chance that a nationwide ACP programme would be cost-effective (see Fig. 2) due to high uncertainty around the key parameters. |
| Monnery et al., 2023 (Monnery et al., 2023)<br>(England)<br>**Aim:** to determine the impact and health economic benefits of ESC for patients living with treatable but not curable cancer.<br>**Enhanced supported care (ESC) services** | **Intervention (specialist or generalist palliative care):** Specialist.<br>**Intervention recipients and sample size:** Enhanced supported care service, n = 4594 cancer patients were observed.<br>**Setting:** Enhanced supported care services for cancer patients in England.<br>**Delivery mode (e.g., remotely online, in person):** In person.<br>**Intervention deliverers (e.g., professionals or paid carers):** Healthcare professionals.<br>**Timing and duration:** Cancer patients followed up from April 2021 to July 2022. (Some died, but some were living with cancer).<br>**Intervention description:** Cost of an enhanced supported care cancer service. | **Study design:** Cost analysis.<br>**Dates of data collection:** From April 2021 to July 2022.<br>**Length of follow-up:** Follow-up 3 months after accessing ESC.<br>**Type of economic evaluation/cost analysis:** Cost analysis<br>**Perspective of analysis:** Taxpayer perspective.<br>**Currency and cost year:** GBP, in 2022.<br>**Discounting:** No.<br>**Sensitivity analysis:** No. | **Outcome:** Costs and outcomes of an enhanced supported care (ESC) cancer service.<br>**Types of costs measured:** The cost of delivery of the service was established by costing professionals' time spent with patients. These data included salaries and local costs (e.g., clinical space) but not medications and capital equipment costs. | **Base case results:** In total, 4594 patients were seen by enhanced supported care services, of whom 1061 died during follow-up. Mean Integrated Palliative Care Scale (IPOS) scores improved across all tumour groups. In total, £1,676,044 was spent delivering Enhanced Supported Care (ESC) across the eight centres. Reductions in secondary care usage for the 1061 patients who died saved a total of £8,490,581.<br>**Sensitivity analysis results:** None reported. |

**Table 20:** Summary of the recent palliative care systematic reviews.

| Citation (Country) | Review details | Included studies | Quality | Key findings | Observations/notes |
| --- | --- | --- | --- | --- | --- |
| **Bajwah et al., 2020**<br>(Bajwah et al., 2020)<br>**(England)** | **Review period:** up to August 2019<br>**Review purpose:** To assess the effectiveness and cost-effectiveness of Hospital based Palliative Care (HSPC) compared to usual care for adults with advanced illness (hereafter patients) and their unpaid caregivers/families.<br>**Included study designs:** Full economic evaluations and costs studies that were conducted alongside an RCT.<br>**Included costs and outcome measures:** Cost of HSPC and Health Related Quality of Life (HRQoL).<br>**Eligibility criteria:** RCTs evaluating the impact of HSPC on outcomes for patients or their unpaid caregivers/ families, or both. | **Number of included studies:** n = 42<br>**Key characteristics:** A total of 31 studies were included in the economic component of this review.<br>4 full economic evaluations compared costs and effects of the intervention and control group between baseline and follow-up.<br>5 partial economic evaluations compared only costs and outcomes without reporting incremental changes or decision criteria.<br>22 studies reported more limited resource use/cost information.<br>**Number of economic evaluations:** n = 4 | High | Of 13 studies reporting costs of HSPC, nine studies found no difference between HSPC, and usual care and two studies favoured HSPC over usual care. The difference in cost was unclear in one study, while another study reported mixed findings with lower cost of hospitalisation in favour of HSPC but no difference in the cost of emergency room visits. Four studies with full economic analysis were inconclusive on the cost-effectiveness of HSPC. |  |
| **Gomes et al., 2013**<br>(Gomes et al., 2013)<br>**(England)** | **Review period:** 1950 to 21<sup>st</sup> November 2012<br>**Review purpose:**<br>1. To quantify the effect of home palliative care services for adult patients with advanced illness and their family caregivers on patients' odds of dying at home.<br>2. to examine the clinical effectiveness of home palliative care services on other outcomes for patients and their caregivers such as symptom control, quality of life, | **Number of included studies:** n = 23<br>**Key characteristics:** 6 economic evaluations reported on total care cost. 5 RCTs and 1 CBA).<br>Canada, Italy, Norway, Sweden, UK, USA<br>**Number of economic evaluations:** n = 6 | High | Brumley, 2007 - intervention n=145; control n=152.<br>Total adjusted mean costs per patient USD7552 lower in intervention group (33% lower; 95% CI - USD12,411 to - USD780; P value = 0.03; R2 0.16). Unadjusted difference: t 3.63; P value <0.001. Time horizon: from enrolment to death, transfer to hospice care or study end (mean survival of 196 days in intervention group and 242 days in control group; 73% patients died). | Only two of the reported studies Brumley, 2007 and Higginson, 2009 are within our review period of 2003 to 2023. |
|  | caregiver distress and satisfaction with care.<br>3. to compare the resource use and costs associated with these services<br>4. to critically appraise and summarise the current evidence on cost-effectiveness.<br>**Included study designs:**<br>randomised controlled trials (RCTs), controlled clinical trials (CCTs), controlled before and after studies (CBAs) and interrupted time series (ITSs).<br>**Included costs and outcome measures:**<br>Resource use costs for home palliative care services and carer costs.<br>**Eligibility criteria:**<br>randomised controlled trials (RCTs), controlled clinical trials (CCTs), controlled before and after studies (CBAs) and interrupted time series (ITSs) evaluating the impact of home palliative care services on outcomes for adults with advanced illness or their family caregivers, or both. |  |  | Currency: 2002 USD<br>Adjusted mean costs per patient<br>Intervention USD2670 ± 12,523.<br>Control USD20,222 ± 30,026<br>Adjusted mean costs per patient per day. Intervention USD95.30<br>Control USD212.80, t - 2.417; P value = 0.02.<br>Higginson, 2009 – Intervention n = 26; control n = 24.<br>Total mean costs per patient<br>GBP1789 lower in intervention group (29% lower; bootstrapped 95% CI - GBP5224 to GBP1902; n.s.); excluding inpatient care and informal care, mean service costs were GBP1195 lower in the intervention group (50% lower; bootstrapped 95% CI - GBP2916 to GBP178; n.s.).<br>Time horizon: 12 weeks from enrolment (only 4 deaths).<br>Currency: 2005 GBP<br>Mean costs per patient.<br>Intervention GBP4294<br>Control GBP6084 |  |
| **Gonzalez-Jaramillo et al., 2021**<br>(Gonzalez-Jaramillo et al., 2021)<br>**(Switzerland)** | **Review period:** between 2013 and 11 February 2019<br>**Review purpose:** To assess the effectiveness of home-based palliative care (HBPC) on reducing hospital visits and whether HBPC lowered healthcare cost.<br>**Included study designs:** retrospective cohort, quasi-experimental, RCT. | **Number of included studies:** n = 21<br>**Key characteristics:**<br>7 of the included studies reported on costs of HBPC.<br>**Number of economic evaluations:**<br>Not stated. | High | Seven studies assessed costs generated by hospitalisations, and all found that inpatient costs were lower in patients who received palliative home care.<br>One RCT conducted among patients with heart failure with 6 months follow-up found that inpatient cost in the group with access to HBPC was at least three times less than the cost in the control arm. |  |
|  | **Included costs and outcome measures:** Costs of HBPC.<br>**Eligibility criteria:** Studies that Included adult palliative population ($\geq 18$ years old), at the end of life, with severe illness or with a disease end-stage and compared hospital visits or healthcare costs of those who received PC at home to those who received usual care. |  |  | Two large retrospective cohorts with a combined total of over 25,000 participants used matched analysis to adjust for confounders and found significantly lower hospitalisation cost among patients with HBPC during the last 3 months, 2 months, 1 month, and two weeks of patients' life.<br>Two studies assessed outpatient cost. Of them, one included the home care cost in the outpatient cost and found higher values for those with access to HBPC. The other one reported no difference in cost at 6 months before death and lower cost in the lasts 3 months, 2 months, and two weeks of life among patient with access to HBPC.<br>One study reported costs derived from visits to the ED and found no difference in none of the time periods |  |
| **Mathew et al., 2020**<br>(Mathew et al., 2020)<br>**(Canada)** | **Review period:** 2006-2017<br>**Review purpose:** To describe and critically appraise economic evaluations of palliative care models and to identify cost-effective models in improving patient-centred outcomes.<br>**Included study designs:** Modelling studies and economic evaluations.<br>**Included costs and outcome measures:** Two studies compared home-based palliative care models to usual care, and one compared home-based palliative care to no<br>care. Effectiveness outcomes included hospital readmission prevented, days at home, and palliative care symptom severity.<br>**Eligibility criteria:** cost-effectiveness of any population group receiving palliative care services were included. | **Number of included studies:** n = 5<br>**Key characteristics:**<br>2 modelling studies from US and England and 3 economic evaluations from England, Australia, and Italy.<br>**Number of economic evaluations:** n = 3 | | All studies suggested that palliative care was cost-effective compared to either usual care or absence of care.<br>Home-based palliative care for individuals affected with multiple sclerosis and their caregivers is cost-effective.<br>home-based palliative care, which led to reduced hospitalisation rates, saved \$23,559 Euros at the programme level | |
| **Simoens et al., 2010)**<br>(Simoens et al., 2010)<br>**(Belgium)** | **Review period:**<br>**Review purpose:**<br>To review the international literature on the costs of treating terminal patients.<br>**Included study designs:**<br>Cohort studies and case studies.<br>**Included costs and outcome measures:**<br>Healthcare resource use costs for hospital care and home palliative care.<br>**Eligibility criteria:**<br>Studies on the costs of treating terminal patients. Inclusion was limited to studies that contrasted costs in different healthcare settings and to studies that compared palliative care with alternative therapeutic approaches for terminal patients. | **Number of included studies:** n = 15<br>**Key characteristics:**<br>Cohort studies and case studies from Spain, Britain, Hong Kong, France, U.S. Italy, and Israel.<br>**Number of economic evaluations:**<br>Not defined |  | **Costs across healthcare settings.**<br>A prospective multicentre centre in Spain found that the total cost per patient amounted to €2,774, and it could be broken down into a hospitalisation cost of €2,390 per patient and other costs (i.e., outpatient clinic and home care) of €384 per patient.<br>The mean costs of palliative care in a UK study amounted to €3,418 for colon cancer, €4,672 for breast cancer, €4,936 for lung cancer, €5,069 for uterus cancer, €6,577 for stomach/oesophagus cancer, €7,086 for prostate cancer, and €9,014 for ovarian cancer.<br>**Costs of treating terminal patients in hospital.**<br>A case series measured and identified the determinants of palliative care costs of hepatocellular carcinoma in Hong Kong. The mean cost for formal health services per patient amounted to €3,546 from first hospitalisation until death.<br>A prospective cohort study calculated costs of palliative care in two hospitals providing general<br>medicine, surgical, and obstetric care, and in two hospitals offering extended care and rehabilitation in France. Total costs per day amounted to €493 for all patients, €547 for patients admitted to hospitals providing general medicine, and €440 for patients admitted to hospitals providing extended care.<br>A U.S. case-control study included 38 patients admitted to a hospital palliative care unit and 38 patients who died outside the palliative care unit and who were cared for by other medical or surgical teams. The palliative care unit generated lower daily charges (59%) and lower daily costs (57%).<br>A cohort study calculated the costs of patients admitted to a hospital palliative care unit as compared with those of the patients admitted to an intensive care unit or any unit other than palliative care in the United States. The cost per day for hospitalised patients during the last 20 days leading to their death was significantly lower in the palliative care unit than in intensive care units and non-palliative care units.<br>A retrospective, observational study of 314 veterans in the U.S. compared costs of palliative care with those of usual care during a terminal hospitalisation. Hospital palliative care was associated with lower inpatient costs per day (-€245) and lower ancillary costs per day (-€100). |  |
|  |  |  |  | A U.S. study adopted a case control design to compare the charges of 164 patients who received an inpatient palliative care consultation with those of 152 inpatients who did not. Mean daily charges amounted to €4,043 for cases and €4,358 for control patients.<br>A U.S. study investigated costs of patients who received an inpatient palliative care consultation with costs of inpatients who received usual care. Palliative care patients discharged alive had net savings of V1,684 in costs per admission and V277 in costs per day. Cost savings originated from reductions in laboratory and intensive care unit costs as compared with usual care patients. Palliative care patients who died had net savings of V4,872 in costs per admission and V371 in costs per day because of reductions in pharmacy, laboratory, and intensive care unit costs as compared with usual care patients.<br>**Costs of treating terminal patients at home.**<br>In an Italian case series, Costs of the home care service amounted to €39.9 per patient per day. This figure covered the costs of the support and coordination team (€8.3), medicines (€14.4), general practice fees (€5.7), medical examinations (€5.0), nursing (€4.2), supplies (€1.6), and specialist consultations (€0.5). One hundred forty-four patients were assigned to one of the following groups: 1) |  |
|  |  |  |  | terminal phase requiring palliative care (89 patients); 2) advanced phase requiring palliative care (31 patients); 3) chronic phase requiring supportive therapy (nine patients); and 4) curable phase requiring supportive therapy (15 patients). The mean monthly costs of healthcare providers, materials and medicines, transfusion support, and laboratory and diagnostic procedures, amounted to €4,533 for the terminal phase, €2,468 for the advanced phase, €1,594 for the chronic phase, and €4,270 for the curable phase.<br>A retrospective, observational study enrolled all patients undergoing palliative care who died of cancer in a Spanish town in 1998. Mean costs per patient were lower for patients receiving home support than for patients receiving standard care management (-€683). |  |
| **Smith et al., 2014**<br>(Smith et al., 2014)<br>**(Ireland)** | **Review period:**<br>2002-2011<br>**Review purpose:**<br>Literature review of available international evidence on the costs and cost-effectiveness of palliative care interventions in any setting (e.g., hospital-based, home-based and hospice care)<br>**Included study designs:**<br>5 randomised controlled trials (RCTs),<br>2 non-RCTs, 34 cohort studies, 2 case studies, 2 before and-after studies and 1 'other' study.<br>**Included costs and outcome measures:**<br>Resource costs | **Number of included studies:** n = 46<br>**Key characteristics:**<br>5 randomised controlled trials (RCTs), 2 non-RCTs, 34 cohort studies, 2 case studies, 2 before and-after studies and 1 'other' study.<br>**Number of economic evaluations:** n = 1 |  | Only one study met the criteria for cost-effectiveness. Higginson et al, 2009 assessed the cost-effectiveness of a short-term palliative care in multiple sclerosis with a randomised phase II trial. Total costs of care £1789 (2005 prices) lower for PC group over 12-week period: bootstrapped 95% CI = -£5224 to £1902. Excluding inpatient acute and informal care, mean service cost £1195 lower for PC group: 95% CI = - £2916 to £178. Control group more likely to be in contact with GPs, receive help from family/friends, admitted or seen in hospital relative to PC group. Cost-effectiveness: point |  |
|  | **Eligibility criteria:**<br>Identify studies that investigate the cost or resource use implications of a 'palliative care intervention' relative to some type of comparator or control.<br>Identify studies that investigate the cost-effectiveness of a 'palliative care intervention' relative to some type of comparator or control. |  |  | estimates indicate that the intervention is cost-saving with equivalent outcomes on the POS-8 scale and improved outcomes on the Zarit Carer Burden Inventory (ZBI). |  |
| **Salamanca-Balen et al 2018**<br>(Salamanca-Balen et al., 2018)<br>(UK) | **Review period:** up to 2015.<br>**Review purpose:**<br>To present results from a systematic review of the international evidence on the costs, resource use and cost-effectiveness of Clinical Nurse Specialist-led (CNS) interventions for patients with palliative care needs, defined as seriously ill patients and those with advanced disease or frailty who are unlikely to be cured, recover, or stabilise.<br>**Included study designs:**<br>37 randomised controlled trials, 22 quasi-experimental studies, 7 service evaluations and other studies, and 13 economic analyses.<br>**Included costs and outcome measures:**<br>Resource use and nurse led interventions.<br>**Eligibility criteria:**<br>Quantitative studies addressing cost-effectiveness of CNS interventions including randomised controlled trials (RCTs), quasi-experimental, before and after, prospective and retrospective cohort studies, case control studies, | **Number of included studies:** n = 79<br>**Key characteristics:**<br>37 randomised controlled trials, 22 quasi-experimental studies, 7 service evaluations and other studies, and 13 economic analyses.<br>**Number of economic evaluations:**<br>n = 13 |  | A cost-utility analysis in the United Kingdom showed that a nurse-led disease management programme for patients with heart disease was cost-effective, generating an additional QALY at an incremental cost of £13,158 per QALY, compared to the control group.<br>Two studies of cancer care by CNSs also demonstrated cost-effectiveness. These included a trial of a UK nurse-led follow-up clinic for patients with colorectal cancer and a 5-year follow-up study in Sweden of patients with breast cancer showing that CNS-led care was cheaper and led to similar outcomes compared to physician-led care (€495 vs € 630 per person, respectively).<br>Two studies classified as economic analyses included a short-term cost-minimisation study examining a telephone follow-up CNS-led intervention for patients with breast cancer in the United Kingdom and a cost analysis of effects of practice nurse-led care for chronic diseases. In both cases, the interventions were associated with higher costs compared to usual care although |  |
|  | analytical cross-sectional studies, service evaluations and economic analyses. |  |  | some patients preferred these care models. |  |
| **Yadav et al., 2020**<br>(Yadav et al., 2020)<br>**(USA)** | **Review period:**<br>January 2008 and July 2018<br>**Review purpose:**<br>To identify and summarise the best available evidence on cost associated with palliative care for patients diagnosed with cancer.<br>**Included study designs:**<br>Retrospective cohort studies, secondary analysis of randomised controlled trial, prospective observational design.<br>**Included costs and outcome measures:**<br>Resource costs for inpatient, outpatient, and ancillary.<br>**Eligibility criteria:**<br>All clinical studies that assessed the cost (outcome) associated with palliative care (intervention) in patients diagnosed with cancer (population) and were conducted in the US (context/setting). | **Number of included studies:**<br>n =16<br>**Key characteristics:**<br>13 retrospective cohort studies.<br>2 secondary analysis of a randomised control trial.<br>1 prospective observational design.<br>**Number of economic evaluations:**<br>Not defined. | | The inpatient cost savings ranged from \$1285 –\$28,270.<br>The outpatient savings were in the range of \$1000 to \$1491.<br>Within the studies that incorporated both inpatient and outpatient services in their palliative care programmes, the total cost savings achieved ranged from \$1000 to \$5198. | |

### 7.3 Quality appraisal

The summary tables for the quality appraisals are presented in Appendix 9.2.

### 7.4 Information available on request

The data that supports the findings of this study are available in the data extraction tables of this report.

## 8.2 Acknowledgements

Ms Mala Mann, Information Specialist, Cardiff University is thanked for allowing us to use part of a search strategy that she has developed for another project for the Palliative Care Evidence Review Service (PaCERS). Dr Sofie Angharad Roberts is thanked for proofreading the rapid review and providing valuable feedback. We wish to thank the Health and Care Research Wales Evidence Centre Public Partnership Group - Praveena Pemmasani and Rashmi Kumar - and stakeholders - Idris Baker of Swansea Bay University Health Board, and Melanie Lewis of Cwm Taf Morgannwg University Health Board - for their time, expertise and input.

## 9. APPENDIX

## List of Appendices

APPENDIX 1: Palliative care Rapid Review search strategy for MEDLINE (via OVID) database

APPENDIX 2: Quality Appraisal Tables

APPENDIX 3: List of inflation calculators used for inflating and converting costs from original currency into GBP

## 9.1 APPENDIX 1: Palliative care Rapid Review search strategy for MEDLINE (via OVID) database

1. Palliative care/
2. Terminal Care/
3. Terminally ill/
4. Hospice care/
5. (“palliative care” or “hospice care” or “end of life care“).tw.
6. ((hospice or terminal*) adj3 (care or caring or ill*)).tw.
7. (“last year of life” or LYOL or “end of life” or “end of their lives“).tw.
8. (end-stage disease* or end stage disease* or end-stage ill* or end stage ill*).tw.
9. 1 or 2 or 3 or 4 or 5 or 6 or 7 or 8
10. “Evidence-Based Practice“/
11. *“Models, Organisational“/
12. “Organisational Innovation“/
13. Diffusion of Innovation/
14. Patient-Centered Care/
15. Health Priorities/
16. “Delivery of Healthcare“/
17. ((integrat* or combined or joined) adj3 (care or service or delivery or strategy* or programme* or management)).tw.
18. “models of healthcare“.tw.
19. (service adj (delivery or innovation or programme* or model*1 or restructur* or chang*)).tw.
20. 10 or 11 or 12 or 13 or 14 or 15 or 16 or 17 or 18 or 19
21. Cost-benefit analysis/
22. Cost-effective*.tw.
23. Cost-benefit.tw.
24. Cost-utility.tw.
25. Cost-consequence.tw.
26. Cost-minimisation.tw.
27. Cost-minimization.tw.
28. Social Return on Investment.tw.
29. SROI.tw.
30. Return on Investment.tw.
31. Economic evaluation.tw.
32. Budget impact.tw.
33. exp “Costs and Cost Analysis“/
34. 21 or 22 or 23 or 24 or 25 or 26 or 27 or 28 or 29 or 30 or 31 or 32 or 33
35. 9 and 20 and 34

## 9.2 APPENDIX 2: Quality Appraisal Tables

### List of quality appraisal tables

Table 21. Quality appraisals – hospital based palliative care. Table 22. Quality appraisals – hospice based palliative care.

**Table 21:**
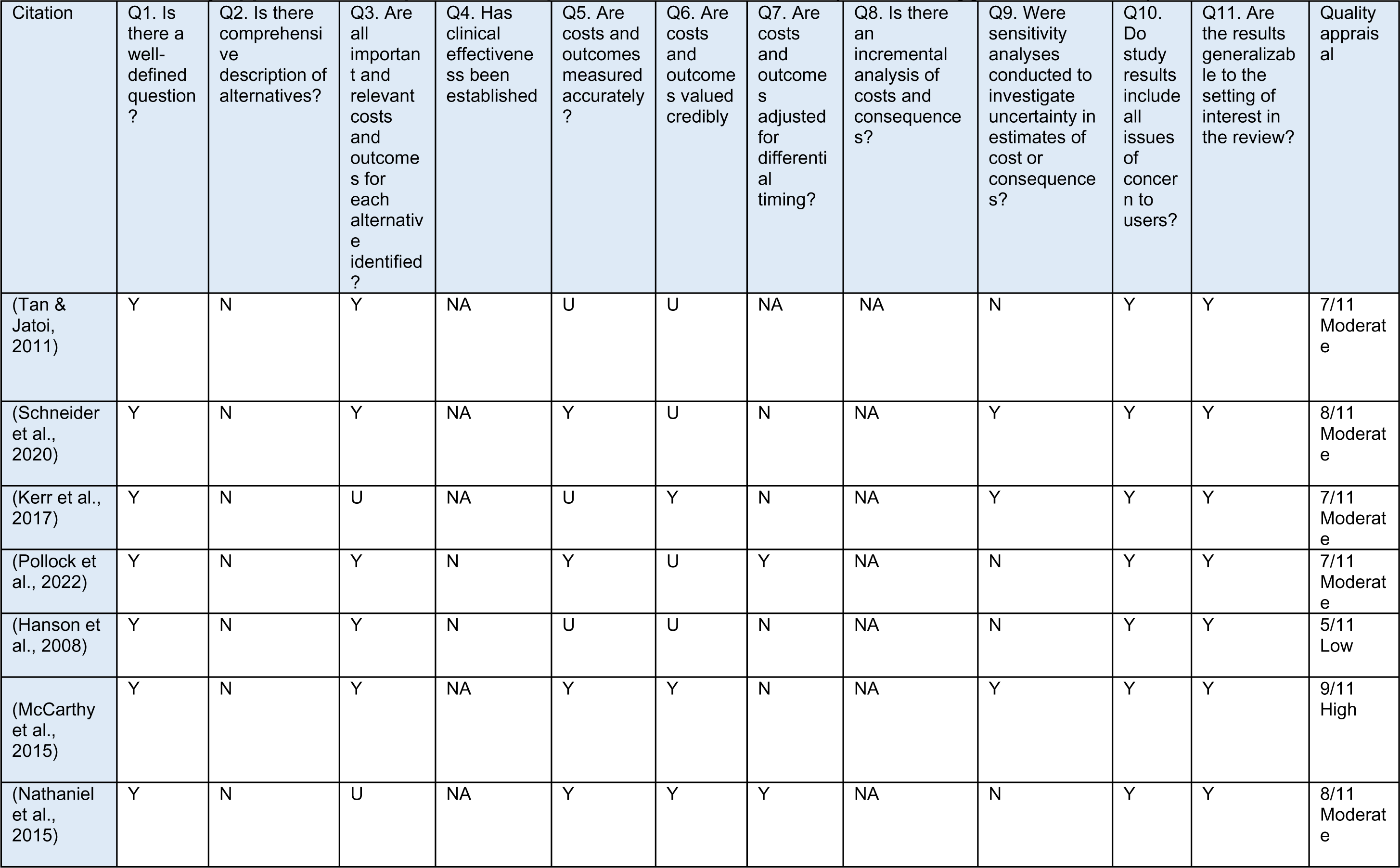
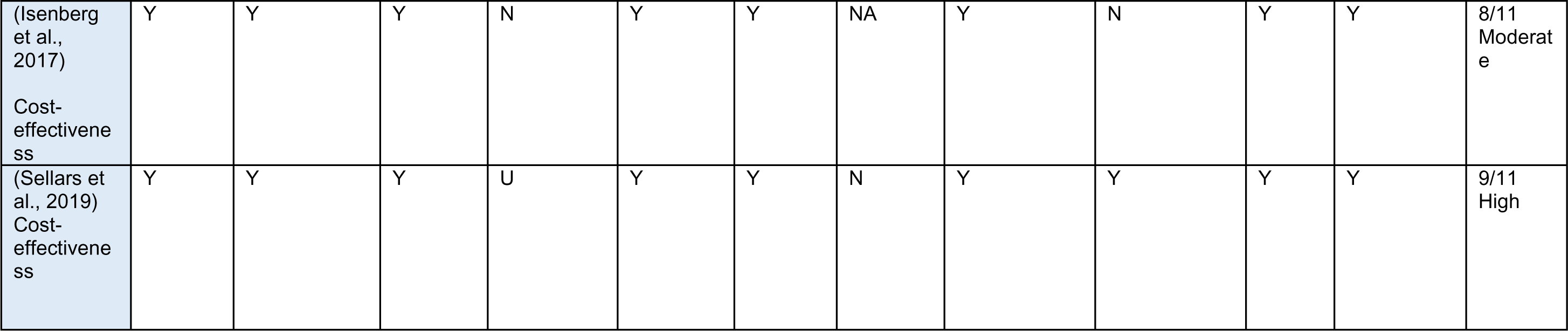
Quality appraisal for hospital focused palliative care cost studies (Joanna Briggs Institute, 2022)

Table 23. Quality appraisals – home/community-based palliative care Table 24. Quality appraisals – combined models of palliative care Table 25. Quality appraisals – systematic reviews The lack of a standardised cost analysis quality appraisal checklist/tool limits the ability to quality appraise such studies (Xu et al., 2021). Due to the nature of such studies, they fail to meet some components of the JBI Critical Appraisal Checklist for Economic Evaluations. Notably, questions surrounding discounting, incremental analyses, and the comprehensive description of alternatives. The authors chose to extend the application of the JBI Critical Appraisal Checklist for Economic Evaluations checklist to cost analyses, through awarding an equal point score to any element marked with an ‘NA’ to not penalise such studies. The scoring algorithm employed by the authors awarded a single point to any element marked Y or NA, while awarding no point to any element marked U or N. These points were totalled out of 11 and quality cut offs created to categorise the evidence into quality levels. For the costing studies, only the ‘cost’ aspect of questions 3, 5 and 6 were considered and the outcome aspect was disregarded due to irrelevancy. Cut off scores are defined in this review as; 11 to 9 out of 11 – high quality, 6 to 8 out of 11 – moderate quality, 0 to 5 out of 11 – low quality.

**Table 22:** Quality appraisal for hospice cost studies (Joanna Briggs Institute, 2022)

| Citation | Q1. Is there a well-defined question? | Q2. Is there a comprehensive description of alternatives? | Q3. Are all important and relevant costs and outcomes for each alternative identified? | Q4. Has clinical effectiveness been established | Q5. Are costs and outcomes measured accurately? | Q6. Are costs and outcomes valued credibly | Q7. Are costs and outcomes adjusted for differential timing? | Q8. Is there an incremental analysis of costs and consequences? | Q9. Were sensitivity analyses conducted to investigate uncertainty in estimates of cost or consequences? | Q10. Do study results include all issues of concern to users? | Q11. Are the results generalizable to the setting of interest in the review? | Quality Appraisal |
| --- | --- | --- | --- | --- | --- | --- | --- | --- | --- | --- | --- | --- |
| Gans et al, 2016<br>(Gans et al., 2016) | Y | N | N | Y | Y | Y | N/A | N/A | N | Y | Y | 8/11<br>Moderate |
| Hughes, 2021<br>(Hughes, 2021) | Y | Y | Y | N | Y | Y | N | N | Y | Y | Y | 8/11<br>Moderate |
| Huskamp et al, 2008<br>(Huskamp et al., 2008) | N | Y | N | Y | Y | Y | N | Y | N | Y | Y | 7/11<br>Moderate |
| Mitchell et al, 2020<br>(Mitchell et al., 2020) | Y | Y | Y | N/A | Y | Y | N/A | N/A | N | Y | Y | 10/11<br>High |

**Table 23:** Quality appraisal for home-based economic evaluation studies (Joanna Briggs Institute, 2022)

| Citation | Q1. Is there a well-defined question? | Q2. Is there a comprehensive description of alternatives? | Q3. Are all important and relevant costs and outcomes for each alternative identified? | Q4. Has clinical effectiveness been established? | Q5. Are costs and outcomes measured accurately? | Q6. Are costs and outcomes valued credibly? | Q7. Are costs and outcomes adjusted for differential timing? | Q8. Is there an incremental analysis of costs and consequences? | Q9. Were sensitivity analyses conducted to investigate uncertainty in estimates of cost or consequences? | Q10. Do study results include all issues of concern to users? | Q11. Are the results generalizable to the setting of interest in the review? | Quality Appraisal |
| --- | --- | --- | --- | --- | --- | --- | --- | --- | --- | --- | --- | --- |
| Amador et al. 2014 | Y | NA | Y | NA | Y | Y | NA | NA | N | Y | Y | 10/11<br>High |
| Bentur et al. 2014 | Y | NA | Y | NA | Y | Y | NA | NA | N | Y | Y | 10/11<br>High |
| Butler et al. 2022 | Y | NA | Y | NA | Y | Y | NA | NA | N | Y | Y | 10/11<br>High |
| Chai et al. 2014 | Y | NA | Y | NA | Y | Y | NA | NA | N | Y | Y | 10/11<br>High |
| Chen et al. 2018 | Y | Y | U | Y | U | U | NA | NA | N | Y | Y | 7/11<br>Moderate |
| Enguidanos et al. 2005 | Y | Y | U | N | U | U | N | NA | N | Y | Y | 5/11<br>Low |
| Gage et al. 2015 | Y | Y | Y | N | Y | Y | NA | NA | N | Y | Y | 9/11<br>High |
| Goldhagen et al. 2016 | Y | NA | U | N | U | U | NA | NA | N | U | Y | 5/11<br>Low |
| Gordon et al. 2022 | Y | Y | U | N | U | U | NA | NA | N | U | N | 4/11<br>Low |
| Johnson et al. 2009 | Y | NA | Y | N | Y | Y | NA | NA | N | N | Y | 8/11<br>Moderate |
| Klinger et al. 2013 | Y | NA | Y | Y | U | U | NA | NA | N | Y | N | 7/11<br>Moderate |
| Maetens et al. 2019 | Y | Y | Y | Y | Y | Y | NA | NA | Y | Y | Y | 11/11<br>High |
| Pham and Krahn, 2014 | Y | Y | Y | Y | Y | Y | NA | Y | Y | Y | Y | 11/11<br>High |
| Rosato et al. 2021 | Y | Y | Y | Y | Y | Y | NA | Y | Y | N | Y | 10/11<br>High |
| Spiro et al. 2020 | Y | NA | Y | Y | Y | Y | NA | NA | N | N | Y | 9/11<br>High |

Answers are: Yes = Y; No = N; Unclear = U; Not Applicable = N/A

**Table 24:** Quality appraisal for combined models of palliative care cost studies (Joanna Briggs Institute, 2022)

| Citation | Q1. Is there a well-defined question? | Q2. Is there a comprehensive description of alternatives? | Q3. Are all important and relevant costs and outcomes for each alternative identified? | Q4. Has clinical effectiveness been established? | Q5. Are costs and outcomes measured accurately? | Q6. Are costs and outcomes valued credibly? | Q7. Are costs and outcomes adjusted for differential timing? | Q8. Is there an incremental analysis of costs and consequences? | Q9. Were sensitivity analyses conducted to investigate uncertainty in estimates of cost or consequences? | Q10. Do study results include all issues of concern to users? | Q11. Are the results generalizable to the setting of interest in the review? | Quality Appraisal |
| --- | --- | --- | --- | --- | --- | --- | --- | --- | --- | --- | --- | --- |
| (Bjørnelv et al., 2020) | Y | Y | Y | N/A | Y | Y | N/A | Y | N | Y | Y | 10/11 High |
| (Brick et al., 2017) | Y | Y | Y | N/A | Y | Y | N/A | Y | N | Y | Y | 10/11 High |
| (Comans et al., 2021) | Y | Y | Y | N/A | Y | Y | N/A | Y | N | Y | Y | 10/11 High |
| (Duncan et al., 2019) | Y | Y | Y | N/A | Y | Y | N/A | Y | N | Y | Y | 10/11 High |
| (Emmert et al., 2013) | Y | Y | Y | Y | Y | NA | Y | N | N | Y | Y | 9/11 High |
| (Hoverman et al., 2020) | Y | Y | Y | N/A | Y | Y | N/A | Y | N | Y | Y | 10/11 High |
| (Kalluri et al., 2020) | Y | Y | Y | N/A | Y | Y | Y | Y | Y | Y | Y | 11/11 High |
| (Kato & Fukuda, 2017) | Y | Y | Y | N/A | Y | Y | N/A | Y | N | Y | Y | 10/11 High |
| (Kim et al., 2022) | Y | Y | Y | N/A | Y | Y | Y | Y | Y | Y | Y | 11/11 High |
| (Lustbader et al., 2017) | Y | Y | Y | N/A | Y | Y | N/A | Y | N | Y | Y | 10/11 High |
| (Nguyen et al., 2017) | Y | Y | Y | Y | Y | Y | Y | Y | Y | Y | Y | 11/11 High |
| (McBride et al., 2011) | Y | Y | Y | N/A | Y | Y | Y | Y | Y | Y | Y | 11/11 High |
| (Monnery et al., 2023) | Y | Y | Y | N/A | Y | Y | N/A | Y | N | Y | Y | 10/11 High |
| (Rolden et al., 2014) | Y | Y | Y | N/A | Y | Y | N/A | Y | N | Y | Y | 10/11 High |
| (Saygili & Çelik, 2019) | Y | Y | Y | Y | Y | Y | N/A | Y | N | Y | Y | 10/11<br>High |
| (Seow et al., 2022) | Y | Y | Y | N/A | Y | Y | Y | Y | Y | Y | Y | 11/11<br>High |
| (Spilsbury & Rosenwax, 2017) | Y | Y | Y | N/A | Y | Y | N/A | Y | N | Y | Y | 10/11<br>High |
| (Tanuseputro et al., 2015) | Y | Y | Y | N/A | Y | Y | N/A | Y | N | Y | Y | 10/11<br>High |
| (Terada et al., 2018) | Y | Y | Y | N/A | Y | Y | N/A | Y | N | Y | Y | 10/11<br>High |
| (Urban et al., 2018) | Y | Y | Y | N/A | Y | Y | N/A | Y | N | Y | Y | 10/11<br>High |
| (Yi et al., 2020) | Y | Y | Y | N/A | Y | Y | Y | Y | Y | Y | Y | 11/11<br>High |
| (Yu et al., 2015) | Y | Y | Y | N/A | Y | Y | N/A | Y | N | Y | Y | 10/11<br>High |

**Table 25:** Quality appraisal table for models of palliative care systematic reviews (Joanna Briggs Institute, 2017)

| Citation | Q1. Is the review question clearly and explicitly stated? | Q2. Were the inclusion criteria appropriate for the review question? | Q3. Was the search strategy appropriate? | Q4. Were the sources and resources used to search for studies adequate? | Q5. Were the criteria for appraising studies appropriate? | Q6. Was critical appraisal conducted by two or more reviewers independently? | Q7. Were there methods to minimize errors in data extraction? | Q8. Were the methods used to combine studies appropriate? | Q9. Was the likelihood of publication bias assessed? | Q10. Were recommendations for policy and/or practice supported by the reported data? | Q11. Were the specific directives for new research appropriate? | Quality Appraisal |
| --- | --- | --- | --- | --- | --- | --- | --- | --- | --- | --- | --- | --- |
| Bajwah et al., 2020 | Y | Y | Y | Y | Y | Y | Y | Y | Y | Y | Y | 11/11<br>High |
| Gomes et al., 2013 | Y | Y | Y | Y | Y | Y | Y | Y | Y | Y | Y | 11/11<br>High |
| Gonzalez-Jaramillo et al., 2021 | Y | Y | Y | Y | Y | Y | Y | Y | Y | Y | Y | 11/11<br>High |
| Mathew et al., 2020 | Y | Y | Y | Y | Y | U | Y | Y | N | N | Y | 9/11<br>High |
| Salamanca-Balen et al 2018 | Y | Y | Y | Y | Y | Y | N | Y | Y | Y | Y | 10/11<br>High |
| Simoens et al., 2010) | Y | Y | Y | Y | Y | N | U | Y | N | Y | Y | 9/11<br>High |
| Smith et al., 2014 | Y | Y | Y | Y | N | N | Y | Y | Y | N | Y | 8/11<br>Moderate |
| Yadav et al., 2020 | Y | Y | Y | Y | Y | U | Y | Y | N | N | Y | 9/11<br>High |

## 9.3 APPENDIX 3: List of inflation calculators used for inflating and converting costs from original currency into GBP

Currency, calculator, and web link:

**AUS$** - Reserve Bank of Australia Inflation Calculator - https://www.rba.gov.au/calculator/

**CAN$** - Bank of Canada Inflation Calculator - https://www.bankofcanada.ca/rates/related/inflation-calculator/

**Euro** – CPI Inflation Calculator: https://www.in2013dollars.com/Euro-inflation

**GPB** – Bank of England Inflation Calculator: https://www.bankofengland.co.uk/monetary-policy/inflation/inflation-calculator

**Japanese Yen** - Inflation Tool - https://www.inflationtool.com/japanese-yen

**Korean Won** – CPI Inflation Calculator: https://www.in2013dollars.com/south-korea/inflation

**Norwegian Krone** – Norges Bank Price Calculator: https://www.norges-bank.no/en/topics/Statistics/Price-calculator-/

**US$** - US Inflation Calculator: https://www.usinflationcalculator.com/

**Currency converter** – XE Currency Converter: https://www.xe.com/currencyconverter/convert/?Amount=1&From=USD&To=GBP

## REFERENCES

Amador, S., Goodman, C., King, D., Ng, Y. T., Elmore, N., Mathie, E., Machen, I., & Knapp, M. (2014). Exploring resource use and associated costs in end-of-life care for older people with dementia in residential care homes. International Journal of Geriatric Psychiatry, 29(7), 758–766. 10.1002/gps.4061

Bajwah, S., Oluyase, A. O., Yi, D., Gao, W., Evans, C. J., Grande, G., Todd, C., Costantini, M., Murtagh, F. E., & Higginson, I. J. (2020). The effectiveness and cost-effectiveness of hospital-based specialist palliative care for adults with advanced illness and their caregivers. Cochrane Database of Systematic Reviews, 2020(9). 10.1002/14651858.CD012780.pub2

Baker, J. I. (2020). Ethics at the end of life. Medicine (United Kingdom), 48(10), 647–650. 10.1016/j.mpmed.2020.07.015

Bedendo, A., Hinde, S., Beresford, B., Papworth, A., Phillips, B., Vasudevan, C., McLorie, E., Walker, G., Peat, G., Weatherly, H., Feltbower, R., Hewitt, C., Haynes, A., Murtagh, F., Noyes, J., Hackett, J., Hain, R., Oddie, S., Subramanian, G., & Fraser, L. (2023). Consultant-led UK paediatric palliative care services: Professional configuration, services, funding. BMJ Supportive and Palliative Care, 1–10. 10.1136/spcare-2023-004172

Bentur, N., Resnizky, S., Balicer, R., & Eilat-Tsanani, T. (2014). Utilization and Cost of Services in the Last 6 Months of Life of Patients With Cancer - With and Without Home Hospice Care. American Journal of Hospice and Palliative Medicine, 31(7), 723–725. 10.1177/1049909113499604

Bjørnelv, G. M. W., Edwin, B., Fretland, Å. A., Deb, P., & Aas, E. (2020). Till death do us part: The effect of marital status on health care utilization and costs at end-of-life. A register study on all colorectal cancer decedents in Norway between 2009 and 2013. BMC Health Services Research, 20(1), 1–13. 10.1186/s12913-019-4794-6

Brereton, L., Clark, J., & Ingleton, C. (2017). What do we know about different models of providing palliative care? Findings from a systematic review of reviews. Palliative, 31(9), 781–797.

Brick, A., Smith, S., Normand, C., O’Hara, S., Droog, E., Tyrrell, E., Cunningham, N., & Johnston, B. (2017). Costs of formal and informal care in the last year of life for patients in receipt of specialist palliative care. Palliative Medicine, 31(4), 356–368. 10.1177/0269216316686277

Butler, C., Wilson, P., Abrahamson, V., Mikelyte, R., Gage, H., Williams, P., Brigden, C., Swash, B., Rees-Roberts, M., Silsbury, G., Goodwin, M., Greene, K., Wee, B., & Barclay, S. (2022). Optimum models of hospice at home services for end-of-life care in England: a realist-informed mixed-methods evaluation. Health and Social Care Delivery Research, 10(24), 1–304. 10.3310/MSAY4464

Chai, H., Guerriere, D. N., Zagorski, B., & Coyte, P. C. (2014). The magnitude, share and determinants of unpaid care costs for home-based palliative care service provision in Toronto, Canada. Health and Social Care in the Community, 22(1), 30–39. 10.1111/hsc.12058

Chen, C. Y., Naessens, J. M., Takahashi, P. Y., McCoy, R. G., Borah, B. J., Borkenhagen, L. S., Kimeu, A. K., Rojas, R. L., Johnson, M. G., Visscher, S. L., Cha, S. S., Thorsteinsdottir, B., & Hanson, G. J. (2018). Improving Value of Care for Older Adults With Advanced Medical Illness and Functional Decline: Cost Analyses of a Home-Based Palliative Care Program. Journal of Pain and Symptom Management, 56(6), 928–935. 10.1016/j.jpainsymman.2018.08.015

Comans, T., Nguyen, K. H., Stafford-Bell, F., & Agar, M. (2021). Cost comparison of different models of palliative care delivery. Australasian Journal on Ageing, 40(1), 90–93. 10.1111/ajag.12843

Davidson, P., Halcomb, E., Hickman, L., Phillips, J., & Graham, B. (2006). Beyond the rhetoric: what do we mean by a “model of care”? Australian Journal of Advanced Nursing, 23(3), 47–55.

Dreamscape and Hospice UK. (2023). Hospice Health Report 2023. https://www.hospiceuk.org/latest-from-hospice-uk/hospice-health-report-2023

Dumont, K., Marcoux, I., Warren, É., Alem, F., Alvar, B., Ballu, G., Bostock, A., Cohen, S. R., Daneault, S., Dubé, V., Houle, J., Minyaoui, A., Rouly, G., Weil, D., Kellehear, A., & Boivin, A. (2022). How compassionate communities are implemented and evaluated in practice: a scoping review. BMC Palliative Care, 21(1), 1–13. 10.1186/s12904-022-01021-3

Duncan, I., Ahmed, T., Dove, H., & Maxwell, T. L. (2019). Medicare Cost at End of Life. American Journal of Hospice and Palliative Medicine, 36(8), 705–710. 10.1177/1049909119836204

Emmert, M., Pohl-Dernick, K., Wein, A., Dörje, F., Merkel, S., Boxberger, F., Männlein, G., Joost, R., Harich, H. D., Thiemann, R., Lamberti, C., Neurath, M. F., Hohenberger, W., & Schöffski, O. (2013). Palliative treatment of colorectal cancer in Germany: Cost of care and quality of life. European Journal of Health Economics, 14(4), 629–638. 10.1007/s10198-012-0408-5

Enguidanos, S., Cherin, D., & Brumley, R. (2005). Home-based palliative care study: site of death, and costs of medical care for patients with congestive heart failure, chronic obstructive pulmonary disease, and cancer. Journal of Social Work in End-of-Life & Palliative Care, 1(3), 37–56 20p. 10.1300/J457v01n03

Gage, H., Holdsworth, L. M., Flannery, C., Williams, P., & Butler, C. (2015). Impact of a hospice rapid response service on preferred place of death, and costs Palliative care in other conditions. BMC Palliative Care, 14(1). 10.1186/s12904-015-0065-4

Gans, D., Hadler, M. W., Chen, X., Wu, S. H., Dimand, R., Abramson, J. M., Ferrell, B., Diamant, A. L., & Kominski, G. F. (2016). Cost Analysis and Policy Implications of a Pediatric Palliative Care Program. Journal of Pain and Symptom Management, 52(3), 329–335. 10.1016/j.jpainsymman.2016.02.020

Goldhagen, J., Fafard, M., Komatz, K., Eason, T., & Livingood, W. C. (2016). Community-based pediatric palliative care for health related quality of life, hospital utilization and costs lessons learned from a pilot study. BMC Palliative Care, 15(1), 1–12. 10.1186/s12904-016-0138-z

Gomes, B., Calanzani, N., Curiale, V., Mccrone, P., & Higginson, I. J. (2013). Effectiveness and cost-effectiveness of home palliative care services for adults with advanced illness and their caregivers. Cochrane Database of Systematic Reviews, 2016(3). 10.1002/14651858.CD007760.pub2

Gonzalez-Jaramillo, V., Fuhrer, V., Gonzalez-Jaramillo, N., Kopp-Heim, D., Eychmüller, S., & Maessen, M. (2021). Impact of home-based palliative care on health care costs and hospital use: A systematic review. Palliative and Supportive Care, 19(4),474–487. 10.1017/S1478951520001315

Gordon, M. J., Le, T., Lee, E. W., & Gao, A. (2022). Home Palliative Care Savings. Journal of Palliative Medicine, 25(4), 591–595. 10.1089/jpm.2021.0142

Hanson, L. C., Usher, B., Spragens, L., & Bernard, S. (2008). Clinical and Economic Impact of Palliative Care Consultation. Journal of Pain and Symptom Management, 35(4), 340–346. 10.1016/j.jpainsymman.2007.06.008

Hatziandreu, E., Archontakis, F., & Daly, A. (2008). The potential cost savings of greater use of home-and hospice-based end of life care in England. The Potential Cost Savings of Greater Use of Home-and Hospice-Based End of Life Care in England. 10.7249/tr642

Hoverman, J. R., Mann, B. B., Phu, S., Nelson, P., Hayes, J. E., Taniguchi, C. B., & Neubauer, M. A. (2020). Hospice or Hospital: The Costs of Dying of Cancer in the Oncology Care Model. Palliative Medicine Reports, 1(1), 92–96. 10.1089/pmr.2020.0023

Hughes, N. (2021). Social Return on Investment: A Mixed Methods Approach to Assessing the Value of Adult Hospice Services in North Wales. January, 1–400.

Huskamp, H. A., Newhouse, J. P., Norcini, J. C., & Keating, N. L. (2008). Variation in patients’ hospice costs. Inquiry, 45(2), 232–244. 10.5034/inquiryjrnl_45.02.232

Isenberg, S. R., Lu, C., McQuade, J., Razzak, R., Weir, B. W., Gill, N., Smith, T. J., & Holtgrave, D. R. (2017). Economic evaluation of a hospital-based palliative care program. Journal of Oncology Practice, 13(5), e408–e415. 10.1200/JOP.2016.018036

Joanna Briggs Institute. (2017). Checklist for Systematic Reviews and Research Syntheses. The Joanna Briggs Institute. http://joannabriggs.org/research/critical-appraisal-tools.htmlwww.joannabriggs.org www.joannabriggs.org

Joanna Briggs Institute. (2022). Checklist for Economic Evaluations. *Jbi*. https://jbi.global/sites/default/files/2020-07/Checklist_for_Analytical_Cross_Sectional_Studies.pdf

Johnson, A. P., Abernathy, T., Howell, D., Brazil, K., & Scott, S. (2009). Resource utilisation and costs of palliative cancer care in an interdisciplinary health care model. Palliative Medicine, 23(5), 448–459. 10.1177/0269216309103193

Kain, V. J., & Chin, S. D. (2020). Conceptually Redefining Neonatal Palliative Care. Advances in Neonatal Care, 20(3), 187–195. 10.1097/ANC.0000000000000731

Kalluri, M., Lu-Song, J., Younus, S., Nabipoor, M., Richman-Eisenstat, J., Ohinmaa, A., & Bakal, J. A. (2020). Health Care Costs at the End of Life for Patients with Idiopathic Pulmonary Fibrosis Evaluation of a Pilot Multidisciplinary Collaborative Interstitial Lung Disease Clinic. Annals of the American Thoracic Society, 17(6), 706–713. 10.1513/AnnalsATS.201909-707OC

Kato, K., & Fukuda, H. (2017). Comparative economic evaluation of home-based and hospital-based palliative care for terminal cancer patients. Geriatrics and Gerontology International, 17(11), 2247–2254. 10.1111/ggi.12977

Kerr, M., Matthews, B., Medcalf, J. F., & O’Donoghue, D. (2017). End-of-life care for people with chronic kidney disease: Cause of death, place of death and hospital costs. Nephrology Dialysis Transplantation, 32(9), 1504–1509. 10.1093/ndt/gfw098

Kim, Y., Han, E., Lee, J., & Kang, H.-T. (2022). Cost-Effectiveness Analysis of Home-Based Hospice-Palliative Care for Terminal Cancer Patients. The Korean Journal of Hospice and Palliative Care, 25(2), 76–84. 10.14475/jhpc.2022.25.2.76

Klinger, C. A., Howell, D., Marshall, D., Zakus, D., Brazil, K., & Deber, R. B. (2013). Resource utilization and cost analyses of home-based palliative care service provision: The Niagara West End-of-Life Shared-Care Project. Palliative Medicine, 27(2), 115–122. 10.1177/0269216311433475

Lustbader, D., Mudra, M., Romano, C., Lukoski, E., Chang, A., Mittelberger, J., Scherr, T., & Cooper, D. (2017). The Impact of a Home-Based Palliative Care Program in an Accountable Care Organization. Journal of Palliative Medicine, 20(1), 23–28. 10.1089/jpm.2016.0265

Maetens, A., Beernaert, K., De Schreye, R., Faes, K., Annemans, L., Pardon, K., Deliens, L., & Cohen, J. (2019). Impact of palliative home care support on the quality and costs of care at the end of life: A population-level matched cohort study. BMJ Open, 9(1), 1–9. 10.1136/bmjopen-2018-025180

Mathew, C., Hsu, A. T., Prentice, M., Lawlor, P., Kyeremanteng, K., Tanuseputro, P., & Welch, V. (2020). Economic evaluations of palliative care models: A systematic review. Palliative Medicine, 34(1), 69–82. 10.1177/0269216319875906

McBride, T., Morton, A., Nichols, A., & Van Stolk, C. (2011). Comparing the costs of alternative models of end-of-life care. Journal of Palliative Care, 27(2), 126–133. 10.1177/082585971102700208

McCarthy, I. M., Robinson, C., Huq, S., Philastre, M., & Fine, R. L. (2015). Cost savings from palliative care teams and guidance for a financially viable palliative care program. Health Services Research, 50(1), 217–236. 10.1111/1475-6773.12203

Mitchell, P. M., Coast, J., Myring, G., Ricciardi, F., Vickerstaff, V., Jones, L., Zafar, S., Cudmore, S., Jordan, J., McKibben, L., Graham-Wisener, L., Finucane, A. M., Hewison, A., Haraldsdottir, E., Brazil, K., & Kernohan, W. G. (2020). Exploring the costs, consequences and efficiency of three types of palliative care day services in the UK: A pragmatic before-and-after descriptive cohort study. BMC Palliative Care, 19(1), 1–9. 10.1186/s12904-020-00624-y

Monnery, D., Tredgett, K., Hooper, D., Barringer, G., Munton, A., Thomas, M., Vijeratnam, N., Godfrey, N., Summerfield, L., Hawkes, K., Staley, P., Holyhead, K., Liu, Y., Lockhart, J., Bass, S., Tavabie, S., White, N., Stewart, E., Droney, J., & Minton, O. (2023). Delivery Models and Health Economics of Supportive Care Services in England: A Multicentre Analysis. Clinical Oncology, 35(6), e395–e403. 10.1016/j.clon.2023.03.002

Nathaniel, J. D., Garrido, M. M., Chai, E. J., Goldberg, G., & Goldstein, N. E. (2015). Cost savings associated with an inpatient palliative care unit: Results from the first two years. Journal of Pain and Symptom Management, 50(2), 147–154. 10.1016/j.jpainsymman.2015.02.023

National Health Service (NHS). (2022). Hospice Care: End of life care. https://www.nhs.uk/conditions/end-of-life-care/hospice-care/

National Institute for Health and Care Excellence (NICE). (2019). End of life care for adults: service delivery. https://www.nice.org.uk/guidance/ng142/resources/end-of-life-care-for-adults-service-delivery-pdf-66141776457925

National Institute for Health and Care Excellence (NICE). (2023). What is palliative care? https://cks.nice.org.uk/topics/palliative-care-general-issues/background-information/definition/#general-palliative-care.

Nguyen, K. H., Sellars, M., Agar, M., Kurrle, S., Kelly, A., & Comans, T. (2017). An economic model of advance care planning in Australia: A cost-effective way to respect patient choice. BMC Health Services Research, 17(1), 1–8. 10.1186/s12913-017-2748-4

NHS England. (2023a). Palliative and End of Life Care. https://www.england.nhs.uk/eolc/#:~:text,=It prevents and relieves suffering,the last year of life.

NHS England. (2023b). Specialist palliative and end of life care services. https://www.england.nhs.uk/wp-content/uploads/2023/01/B1674-specialist-palliative-and-end-of-life-care-services-adult-service-specification.pdf

Noyes, J., Edwards, R. T., Hastings, R. P., Hain, R., Totsika, V., Bennett, V., Hobson, L., Davies, G. R., Humphreys, C., Devins, M., Spencer, L. H., & Lewis, M. (2013). Evidence-based planning and costing palliative care services for children: novel multi-method epidemiological and economic exemplar.

Noyes, J., Hastings, R. P., Lewis, M., Hain, R., Bennett, V., Hobson, L., & Spencer, L. H. (2013). Planning ahead with children with life-limiting conditions and their families: development, implementation and evaluation of “My Choices”. BMC Palliative Care, 12(5), 5. 10.1186/1472-684X-12-5

Page, M., McKenzie, J., Bossuyt, P., Boutron, I., Hoffmann, T., & Mulrow, C. (2021). The PRISMA 2020 statement: an updated guideline for reporting systematic reviews. BMJ, 10(89). doi10.1186/s13643-021-01626-4

Papworth, A., Hackett, J., Beresford, B., Murtagh, F., Weatherly, H., Hinde, S., Bedendo, A., Walker, G., Noyes, J., Oddie, S., Vasudevan, C., Feltbower, R. G., Phillips, B., Hain, R., Subramanian, G., Haynes, A., & Fraser, L. K. (2023). Regional perspectives on the coordination and delivery of paediatric end-of-life care in the UK: a qualitative study. BMC Palliative Care, 22(1), 1–11. 10.1186/s12904-023-01238-w

Perea-Bello, A. H., Trapero-Bertran, M., & Dürsteler, C. (2023). Palliative Care Costs in Different Ambulatory-Based Settings: A Systematic Review. PharmacoEconomics, 0123456789. 10.1007/s40273-023-01336-w

Pham, B., & Krahn, M. (2014). End-of-life care interventions: An economic analysis. Ontario Health Technology Assessment Series, 14(18), 1–70. https://pubmed.ncbi.nlm.nih.gov/26339303/

Pollock, C., James, G., Garcia Sanchez, J. J., Arnold, M., Carrero, J. J., Lam, C. S. P., Chen, H., Nolan, S., & Pecoits-Filho, R. (2022). Cost of End-of-Life Inpatient Encounters in Patients with Chronic Kidney Disease in the United States: A Report from the DISCOVER CKD Retrospective Cohort. Advances in Therapy, 39(3), 1432–1445. 10.1007/s12325-021-02010-3

Rolden, H. J. A., van Bodegom, D., & Westendorp, R. G. J. (2014). Variation in the costs of dying and the role of different health services, socio-demographic characteristics, and preceding health care expenses. Social Science and Medicine, 120, 110–117. 10.1016/j.socscimed.2014.09.020

Rosato, R., Pagano, E., Giordano, A., Farinotti, M., Ponzio, M., Veronese, S., Confalonieri, P., Grasso, M. G., Patti, F., & Solari, A. (2021). Living with severe multiple sclerosis: Cost-effectiveness of a palliative care intervention and cost of illness study. Multiple Sclerosis and Related Disorders, 49(January). 10.1016/j.msard.2021.102756

Salamanca-Balen, N., Seymour, J., Caswell, G., Whynes, D., & Tod, A. (2018). The costs, resource use and cost-effectiveness of Clinical Nurse Specialist–led interventions for patients with palliative care needs: A systematic review of international evidence. Palliative Medicine, 32(2), 447–465. 10.1177/0269216317711570

Saygili, M., & Çelik, Y. (2019). An evaluation of the cost-effectiveness of the different palliative care models available to cancer patients in Turkey. European Journal of Cancer Care, 28(5), 1–10. 10.1111/ecc.13110

Schneider, P. P., Pouwels, X. G. L. V., Passos, V. L., Ramaekers, B. L. T., Geurts, S. M. E., Ibragimova, K. I. E., De Boer, M., Erdkamp, F., Vriens, B. E. P. J., Van De Wouw, A. J., Den Boer, M. O., Pepels, M. J., Tjan-Heijnen, V. C. G., & Joore, M. A. (2020). Variability of cost trajectories over the last year of life in patients with advanced breast cancer in the Netherlands. PLoS ONE, 15(4), 1–14. 10.1371/journal.pone.0230909

Sellars, M., Clayton, J. M., Detering, K. M., Tong, A., Power, D., & Morton, R. L. (2019). Costs and outcomes of advance care planning and end-of-life care for older adults with endstage kidney disease: A person-centred decision analysis. PLoS ONE, 14(5), 1–11. 10.1371/journal.pone.0217787

Seow, H., Barbera, L. C., McGrail, K., Burge, F., Guthrie, D. M., Lawson, B., Chan, K. K. W., Peacock, S. J., & Sutradhar, R. (2022). Effect of Early Palliative Care on End-of-Life Health Care Costs: A Population-Based, Propensity Score–Matched Cohort Study. JCO Oncology Practice, 18(1), e183–e192. 10.1200/op.21.00299

Simoens, S., Kutten, B., Keirse, E., Berghe, P. Vanden, Beguin, C., Desmedt, M., Deveugele, M., Léonard, C., Paulus, D., & Menten, J. (2010). The costs of treating terminal patients. Journal of Pain and Symptom Management, 40(3), 436–448. 10.1016/j.jpainsymman.2009.12.022

Smith, S., Brick, A., O’Hara, S., & Normand, C. (2014). Evidence on the cost and cost-effectiveness of palliative care: A literature review. Palliative Medicine, 28(2), 130–150. 10.1177/0269216313493466

Spilsbury, K., & Rosenwax, L. (2017). Community-based specialist palliative care is associated with reduced hospital costs for people with non-cancer conditions during the last year of life. BMC Palliative Care, 16(1), 68. 10.1186/s12904-017-0256-2

Spiro, S., Ward, A., Sixsmith, J., Graham, A., & Varvel, S. (2020). The Cost of Visit-based Home Care for up to Two Weeks in the Last Three Months of Life: APilot Study of Community Care Based at a Hospice-at-home Service in South East of England. Journal of Community Health Nursing, 37(4), 203–213. 10.1080/07370016.2020.1809856

Tan, T. S., & Jatoi, A. (2011). End-of-life hospital costs in cancer patients: Do advance directives or routes of hospital admission make a difference? Oncology, 80(1–2), 118–122. 10.1159/000328279

Tanuseputro, P., Wodchis, W. P., Fowler, R., Walker, P., Bai, Y. Q., Bronskill, S. E., & Manuel, D. (2015). The health care cost of dying: A population-based retrospective cohort study of the last year of life in Ontario, Canada. PLoS ONE, 10(3), 1–11. 10.1371/journal.pone.0121759

Terada, T., Nakamura, K., Seino, K., Kizuki, M., & Inase, N. (2018). Cost of shifting from healthcare to long-term care in later life across major diseases: analysis of end-of-life care during the last 24 months of life. Journal of Rural Medicine, 13(1), 40–47. 10.2185/jrm.2955

The Kings Fund. (2018). End-of-life care. https://www.kingsfund.org.uk/sites/default/files/field/field_document/end-of-life-care-gp-inquiry-research-paper-mar11.pdf

The National Gold Standards Framework (GSF) Centre in End of Life Care. (2022). The Gold Standards Framework. https://www.goldstandardsframework.org.uk/

Urban, R. R., He, H., Alfonso, R., Hardesty, M. M., & Goff, B. A. (2018). The end of life costs for Medicare patients with advanced ovarian cancer. Gynecologic Oncology, 148(2), 336–341. 10.1016/j.ygyno.2017.11.022

Wheatley, V. J., & Baker, J. I. (2007). “Please, I want to go home”: Ethical issues raised when considering choice of place of care in palliative care. Postgraduate Medical Journal, 83(984), 643–648. 10.1136/pgmj.2007.058487

Wichmann, A. B., Goltstein, L. C. M. J., Obihara, N. J., Berendsen, M. R., Van Houdenhoven, M., Morrison, R. S., Johnston, B. M., Engels, Y., Berendsen, M., Goltstein, L., Knol, E., Kool, M., Nienhuis, W., Nies, L., Obihara, N., Pieksma, J., & Rovers, J. (2020). QALY-time: experts’ view on the use of the quality-adjusted life year in cost-effectiveness analysis in palliative care. BMC Health Services Research, 20(1), 1–7. 10.1186/s12913-020-05521-x

World Health Organisation (WHO). (2023). Palliative Care. https://www.who.int/news-room/fact-sheets/detail/palliative-care#:∼:text=Palliative care uses a team,the human right to health.

Yadav, S., Heller, I. W., Schaefer, N., Salloum, R. G., Kittelson, S. M., Wilkie, D. J., & Huo, J. (2020). The health care cost of palliative care for cancer patients: a systematic review. Supportive Care in Cancer, 28(10), 4561–4573. 10.1007/s00520-020-05512-y

Yi, D., Johnston, B. M., Ryan, K., Daveson, B. A., Meier, D. E., Smith, M., McQuillan, R., Selman, L., Pantilat, S. Z., Normand, C., Morrison, R. S., & Higginson, I. J. (2020). Drivers of care costs and quality in the last 3 months of life among older people receiving palliative care: A multinational mortality follow-back survey across England, Ireland and the United States. Palliative Medicine, 34(4), 513–523. 10.1177/0269216319896745

Yu, M., Guerriere, D. N., & Coyte, P. C. (2015). Societal costs of home and hospital end-of-life care for palliative care patients in Ontario, Canada. Health and Social Care in the Community, 23(6), 605–618. 10.1111/hsc.12170

Ziwary, S. R., Samad, D., Johnson, C. D., & Edwards, R. T. (2017). Impact of place of residence on place of death in Wales: An observational study. BMC Palliative Care, 16(1), 1–6. 10.1186/s12904-017-0261-5

